# Proteomic Specificity of Soft Contact Lenses for Tear Protein Sampling

**DOI:** 10.1101/2023.05.17.23290135

**Authors:** Robert K. Roden, Fangfang Jiang, Nathan Zuniga, Alyssa Nitz, Rebecca S. Burlett, Joshua C. Wright, Caleb Shelton, Alex Reed, Samantha Latham, Connor O. Roper, Leena M. Patil, P. Christine Ackroyd, Samuel H. Payne, John C. Price, Kenneth A. Christensen

**Affiliations:** Department of Chemistry & Biochemistry, Brigham Young University, Provo, UT, 84602, USA; College of Optometry, Rocky Mountain University of Health Professions, Provo, UT, 84606, USA; Department of Biology, Brigham Young University, Provo, UT, 84602, USA

**Keywords:** Tear Sampling, Mass Spectrometry, Proteomics, Soft Contact Lenses, Etafilcon A, Verofilcon A

## Abstract

Soft contact lenses (SCLs) have recently been introduced as an alternative method for human tear protein sampling. However, SCLs are available in a variety of chemical compositions which affect protein binding specificity. Here we analyzed 8 different SCL materials to identify an optimal lens for tear protein sampling. Polymer contamination, mass spectrometry (MS) sample preparation method, total protein capture, individual protein specificity, and SCL cost were all assessed. Using a filter-aided sample prep (FASP) method with 4M guanidine for protein removal, only etafilcon A and verofilcon A did not have significant polymer contamination. Polymer was successfully removed using phosphate buffered saline (PBS) with S-Trap columns for all SCL materials, though yielding a slightly lower number of protein identifications per sample. Minor quantitative differences were observed between SCL materials. However, we also saw significant intersubject variation in protein abundance. Of all the assessed SCL materials, verofilcon A lenses yielded the most total protein while comfilcon A and senofilcon A had the least protein variability. As a newly released daily disposable modality (Precision 1, Alcon), verofilcon A has one of the longest predictable production schedules and one of the lowest costs per lens, making it beneficial for large-scale experiments and diagnostics. Furthermore, we demonstrate how protein binding bias with SCL tear sampling is useful for intra-experiment normalization. Overall, these experiments have led us to optimize our previous protocol for SCL tear protein sampling, highlighting important differences between SCL materials and identifying etafilcon A and verofilcon A as optimal materials for tear protein sampling.

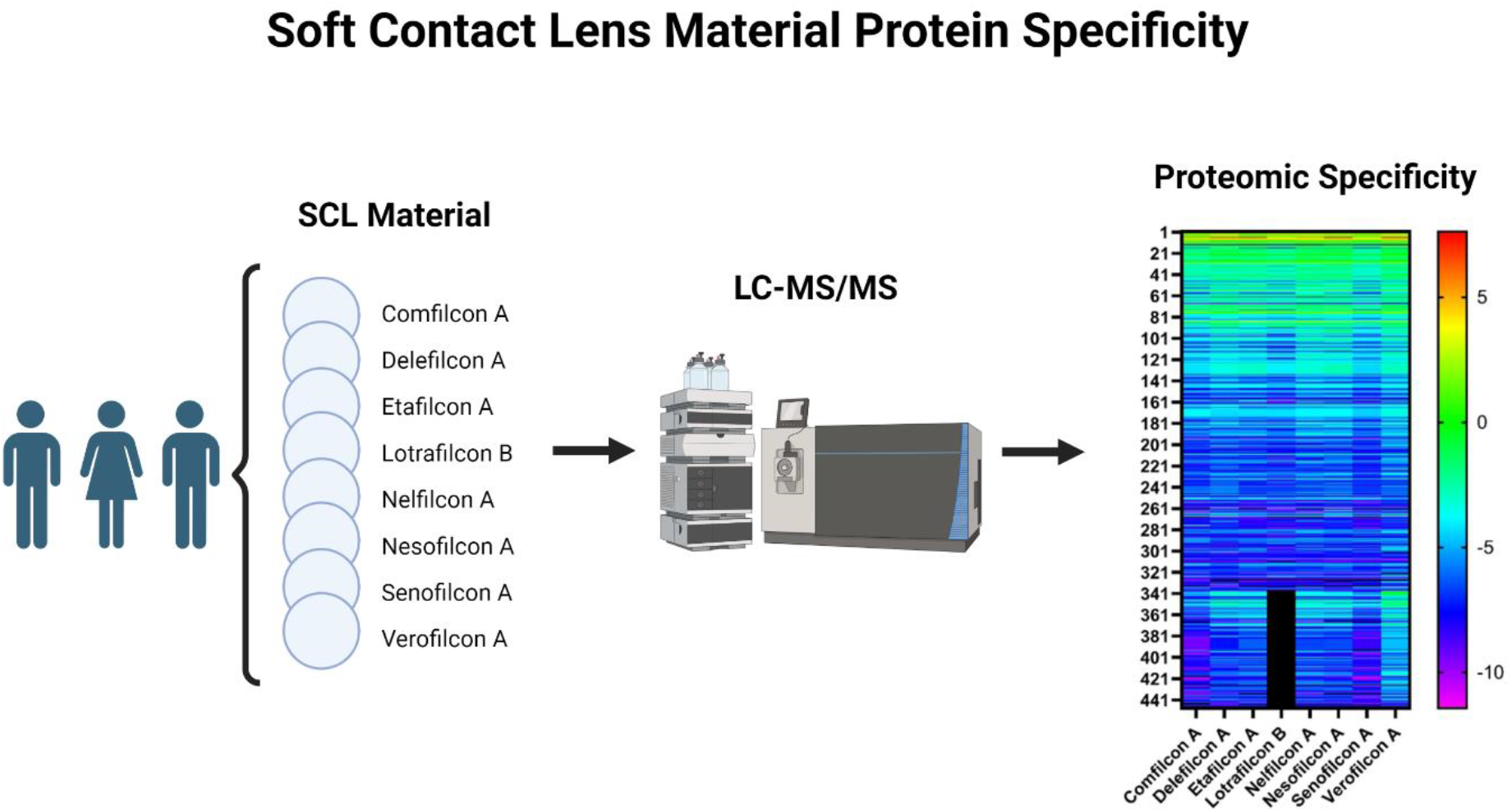

## Introduction

Tear sampling methodology significantly affects both subjects and researchers. For example, total protein for mass spectrometry (MS), subject comfort, ease of sampling, and tear type are all influenced by the sampling method [1, 2]. In addition, significant proteomic differences exist between sampling approaches [1, 2]. Thus, it is important for researchers to determine which type of tear sampling best accomplishes the purposes of their research.

Our recent work demonstrated that soft contact lenses (SCLs) can be used to sample basal tears for proteomics [2]. When comparing SCLs to currently recommended tear sampling methods (microcapillary tubes [MCT] and Schirmer strips [SS]) [3], SCLs were shown to be comparable to MCT in both protein quality and quantity. In our hands, SCLs and MCTs did not show visible signs of eye irritation and had >86 % similarity in protein identifications. SCLs were rated above SSs in both comfort and overall experience and sampled basal tears rather than the reflex tears elicited with SS. Our original study investigated a limited number of SCL materials. However, an important step for continued method development is to understand how different SCL materials affect tear protein sampling.

SCLs vary significantly in chemical composition and have been classified by silicone content, water content, and ionicity [4]. Importantly, protein deposition profiles have been reported to vary based on contact lens material [5]. We therefore hypothesize that SCL chemistry affects tear protein sampling specificity and that certain SCL materials are optimal for tear sampling.

Using MS-based proteomics, we characterized SCL protein sampling specificity by analyzing tears sampled by 8 different SCL materials in 3 human subjects. Tear proteins were compared by protein quantity and quality to determine if there was an optimal lens material for tear sampling. Our data shows that while most SCLs identify the same protein species, individual quantitative protein biases are evident, and some materials introduce polymer contamination. Furthermore, protein sampling specificity can be advantageous and allow for inter-experiment normalization. These optimizations improve SCL tear sampling as an attractive method for a variety of applications, such as tear protein biomarker discovery and clinical diagnostics.

## Materials and Methods

### Human Subject Enrollment & Study Design

Human subjects research was performed in accordance with the Declaration of Helsinki; approval was granted by the Internal Review Board at Brigham Young University (IRB2022-295). Samples were collected at Alpine Vision Center (AVC) (Saratoga Springs, UT). Subjects were educated on the purposes, risks, and benefits of the study. Informed consent was obtained before subject enrollment and privacy rights of human subjects were observed. Subjects who were 18 years or younger and pregnant women were excluded from the study. One female and two male subjects between the ages of 21 and 34 were recruited. All subjects reported having previously used SCLs for at least 1 year.

### Soft Contact Lens Tear Sampling

At the beginning of the study, all subjects were instructed on proper SCL insertion and removal techniques. Each subject was then given 9 randomized sets of SCLs (balafilcon A [Purevision2, Bausch + Lomb], comfilcon A, [Biofinity, CooperVision], delefilcon A [Dailies Total 1, Alcon], etafilcon A [Acuvue 1-day Moist, Johnson & Johnson Vision], lotrafilcon B [Air Optix Plus Hydraglyde, Alcon], nelfilcon A [DAILIES Aqua Comfort Plus, Alcon], nesofilcon A [Biotrue 1 day, Bausch + Lomb], senofilcon A [Acuvue Oasys, Johnson & Johnson Vision], and verofilcon A [Precision1, Alcon]). SCLs were randomly ordered and given to subjects who used a clean set of nitrile powder-free exam gloves (NIGHT ANGEL, Adenna) to insert contacts on their own eyes. After 5 minutes, subjects donned a fresh set of gloves, removed the SCLs, and placed them directly into microcentrifuge tubes. This process was repeated until tears were sampled with all SCL sets. SCLs were then cold chain transported to the sample preparation lab and stored at -80 °C.

### MS Sample Preparation

Samples were first prepped using 4M guanidine and the Filter Aided Sample Prep (FASP) method, as described previously [2]. Upon discovery of polymer contamination in our test samples, we used phosphate buffered saline (PBS, Genesse Scientific) and S-Trap (Protifi) micro spin columns to remove contaminants. Specifically, SCLs were thawed and proteins were removed from SCLs by sonicating for 10 min, then boiling for 5 min in 400 μL PBS. Total protein was then measured using the Pierce BCA kit with standards in PBS. All tear samples were normalized to 22 μg per sample. Proteins were then digested and collected using the S-Trap micro spin column digestion protocol with 5% SDS in each sample. The filtrate was then transferred to MS vials and labeled using the TMT10-plex kit Isobaric label reagent set (ThermoFisher Scientific). Three TMT 10-plex samples were created, each consisting of 9 tear samples using different SCLs from the same human subject and 1 pooled sample. A balafilcon A sample from subject 1 and a lotrafilcon B from subject 3 were failed samples.

### Mass Spectrometry

Tear peptides were analyzed by liquid chromatography-tandem mass spectrometry (LC-MS/MS) using instruments and methods as reported previously [2].

### Data Analysis

MS data was analyzed using Peaks software (Bioinformatics Solutions Inc.). Normalization of each TMT 10-plex was performed at the peptide level by normalizing to the pooled sample TMT tag using PEAKSQ. Spectrum filter settings were set to a false discovery rate (FDR) of 1% (−10logP ≥ 26.4) and quality ≥ 9.5. Proteins filter settings were significance ≥ 0, fold change ≥ 1, and at least 1 unique peptide. All spectra with intensity ≤1E2 were excluded. For quantitative analysis, the data was log_2_ transformed, zero’s replaced blanks, then data was normalized to the average intensity of lysozyme C, lactoferrin, lipocalin-1, polymeric immunoglobulin receptor, clusterin, and all proteins with a coefficient of variance (CV) <0.05.

The Shapiro-Wilk test was used to test data normality (Supplementary Table 1). FDR was accounted for by adjusting p-values using the R-Stats p.adjust function with the method specified as Benjamini-Hochberg. Adjusted p-values <0.05 were considered significant. The overall quantitative total protein abundance between SCL materials was calculated by averaging each protein’s MS intensity from all subjects for each SCL material, then averaging all of the individual protein intensity averages. Statistical analyses of quantitative comparisons for overall protein binding specificity as well as for individual proteins between SCL materials were made in R using Skillings-Mack and in GraphPad Prism using Kruskal-Wallis tests. Wilcoxon Signed-Rank tests were performed in R to compare quantitative protein differences between etafilcon A and verofilcon A.

Next, protein abundance was compared to molecular weight, aromaticity, instability index, gravy, isoelectric point, charge at pH 7.0, and molar extinction coefficient as determined by Biopython’s ProtParam module. As we had a small sample size and most of our data was not normally distributed, Spearman correlations were then made for each SCL material. Normalization methods were compared using unnormalized data, the average of the pooled sample, and the average of lysozyme C, lactoferrin, lipocalin-1, polymeric immunoglobulin receptor, clusterin, and all proteins with a coefficient of variance (CV) <0.05.

Purevision2 (balafilcon A) lenses were discontinued in 2023. As such, we did not include them in our analyses. The raw data is available online at the Mendeley Data repository [6].

## Results

In determining an optimal SCL material for MS proteomics and diagnostics, we propose that the ideal SCL for proteomics and diagnostics would have the features listed in Table 1. We began by selecting 9 different SCL lenses for testing. The US Food and Drug Administration (FDA) has grouped SCLs by material group (Table 2) and SCL choices were designed to sample as many material groups as possible from currently available SCLs [7, 8]. Our study design involved placing each lens in 3 different subjects for 5 minutes, then eluting proteins for analysis as previously described [2]. Unfortunately, many of the SCL materials had significant polymer contamination detected by MS when using the filter assisted sample preparation (FASP) method (Figure 1, Supplementary Figure 1). Only etafilcon A and verofilcon A did not show signs of polymer contamination.

**Table 1.**
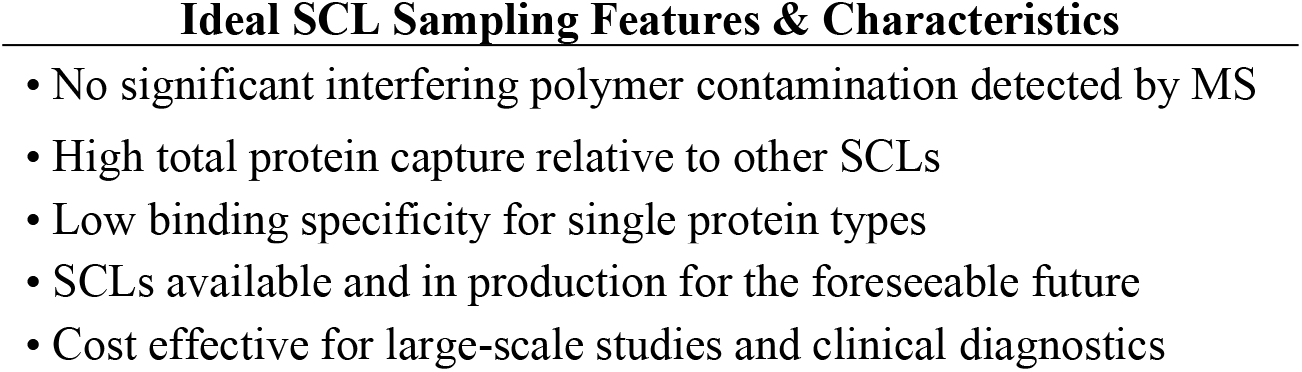
Features and Characteristics of an Ideal SCL Material for Tear Film Protein Sampling

**Table 2.** SCL Materials Representing Each of the US FDA SCL Material Groups

| **Material** | **<sup>1</sup> Group** | **<sup>2</sup> Water Content** | **Ionicity** |
| --- | --- | --- | --- |
| Comfilcon A | 5 | Low | Non-Ionic |
| Delefilcon A | 5 | Low | Non-Ionic |
| Etafilcon A | 4 | High | Ionic |
| Lotrafilcon B | 5 | Low | Non-Ionic |
| Nelfilcon A | 2 | High | Non-Ionic |
| Nesofilcon A | 2 | High | Non-Ionic |
| Senofilcon A | 5 | Low | Non-Ionic |
| Verofilcon A | 5B | High | Non-Ionic |

<sup>1</sup> US FDA SCL group based on material
<sup>2</sup> $\geq 50\%$ water content = High, $< 50\%$ water content = Low

**Figure 1.**
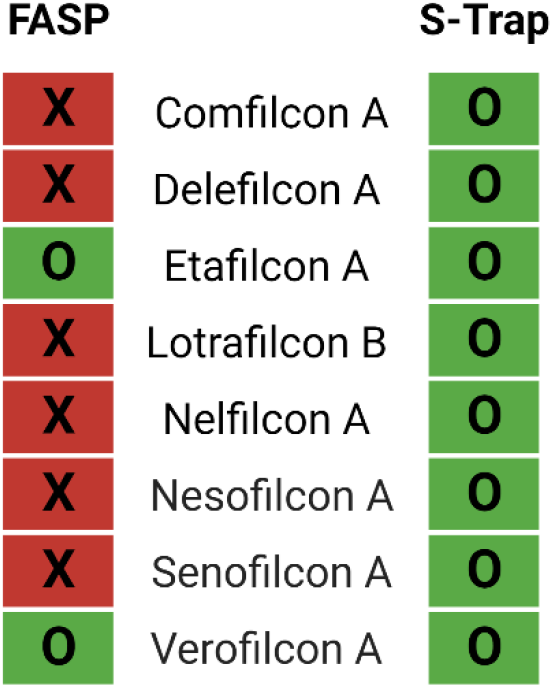
Polymer Contamination as Detected by MS for SCL Materials Tested. Red “X” boxes indicate contamination while green “O” boxes indicate no contamination.

Polymer contamination can sometimes be eliminated by alteration of sample prep methods. Instead of using guanidine to elute proteins from SCLs, we eluted protein with PBS followed by S-Trap sample preparation. The altered elution and sample preparation strategy yielded an average of 42 ± 14 μg total protein across all SCL materials (Figure 2), comparable to our previous method using 4M guanidine for protein removal [2]. Importantly, under these conditions we observed no polymer contamination for any of the SCL materials (Figure 1).

**Figure 2.**
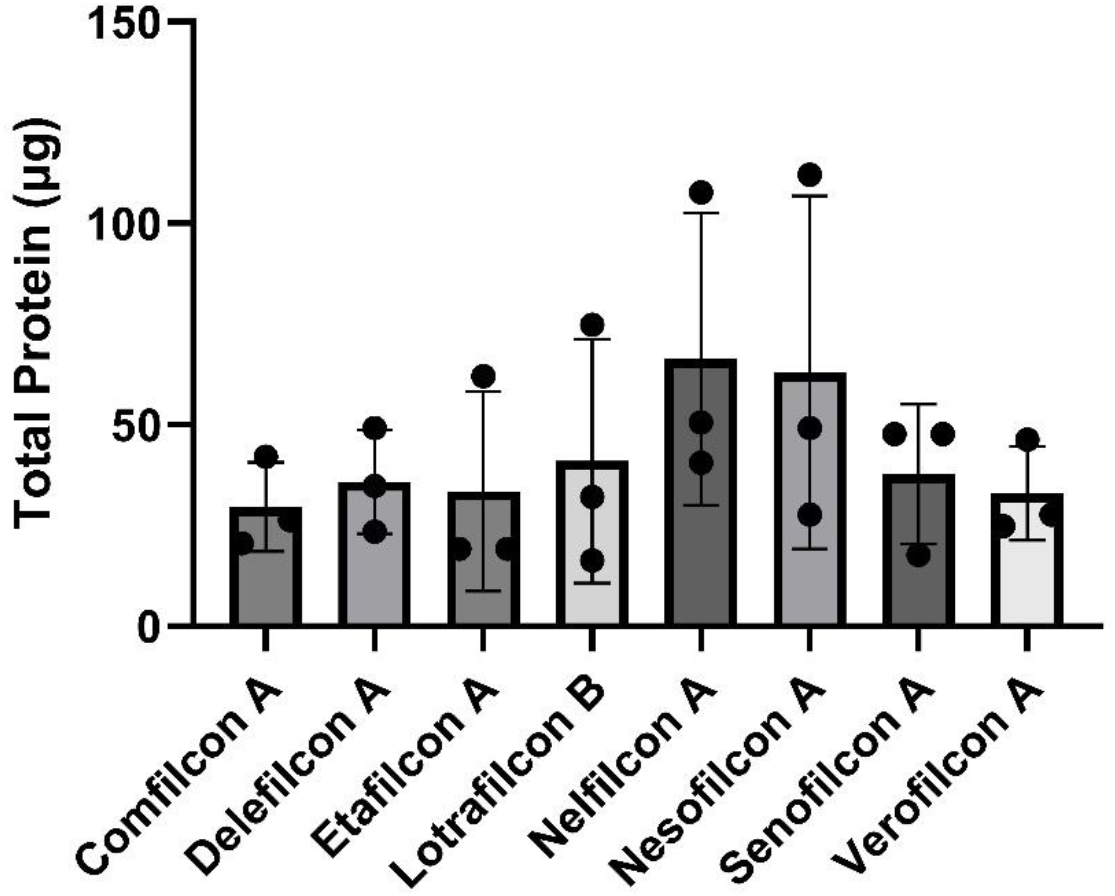
*In vivo* total protein capture by SCL material as measured by BCA protein assay

As SCL chemical compositions vary, we hypothesized that SCLs had unique protein binding specificity that depended on their material. The SCL materials tested in this study were chosen to represent a diverse chemical spectrum. However, some FDA groups were not represented due to older SCL materials being discontinued. The materials tested in this study can be divided into two main groups: non-ionic, low-water, silicone hydrogel (SiHy, group V) and non-ionic, high-water, non-silicone hydrogels (group II). We also tested Etafilcon A, an ionic, high-water, non-silicone hydrogel (group IV).

We analyzed the qualitative and quantitative protein profiles for each SCL material tested. When we assessed the number of total protein species identified, most materials performed similarly; only minor differences were found between materials (Figure 3A, Supplementary Table 2). However, significant differences were observed between total protein species identified between individual subjects (Figure 3B, Supplementary Table 3). We also observed that the abundance of individual proteins varied between SCL materials, as shown in Figure 4. Given our small sample size and the large inter-subject protein differences previously noted, the loss of one lotrafilcon B sample led to a significant loss in overall protein identifications, making comparisons of lotrafilcon B to other lenses challenging. Nevertheless, it is clear that individual protein specificity varies by SCL material.

**Figure 3.**
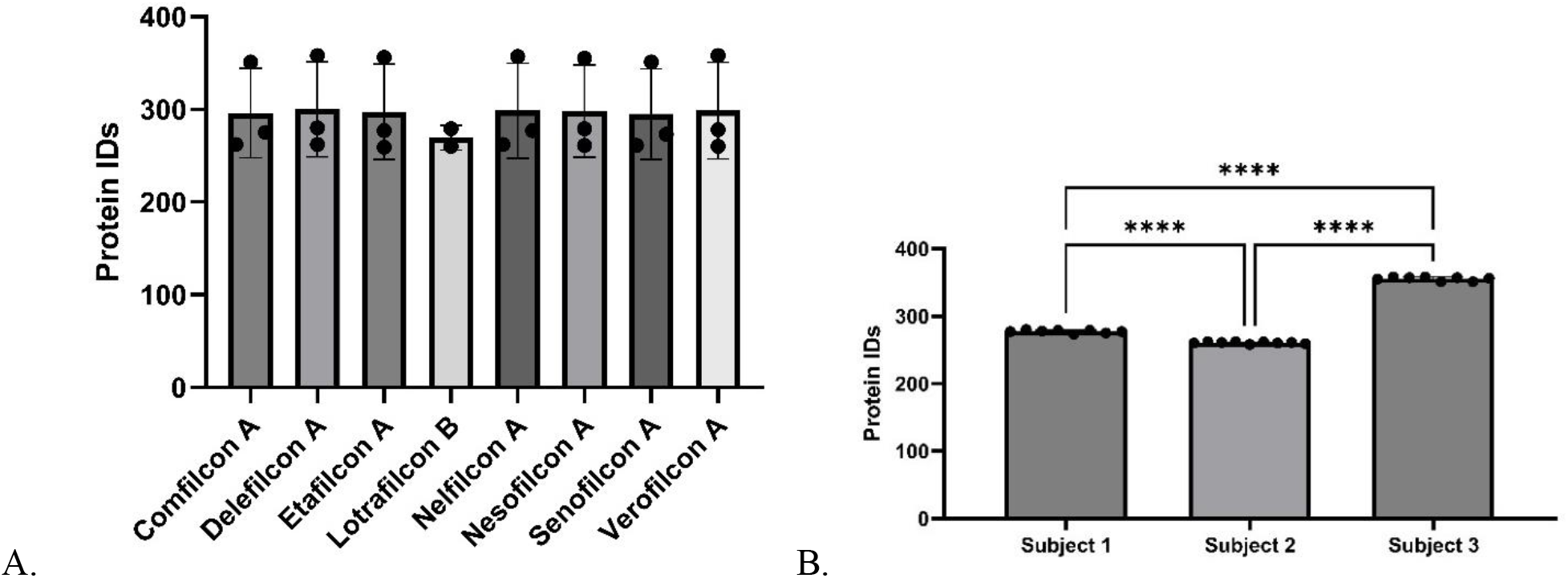
SCL tear sampling protein identifications A) by material, B) by subject. **** = p value ≤ 0.0001.

**Figure 4.**
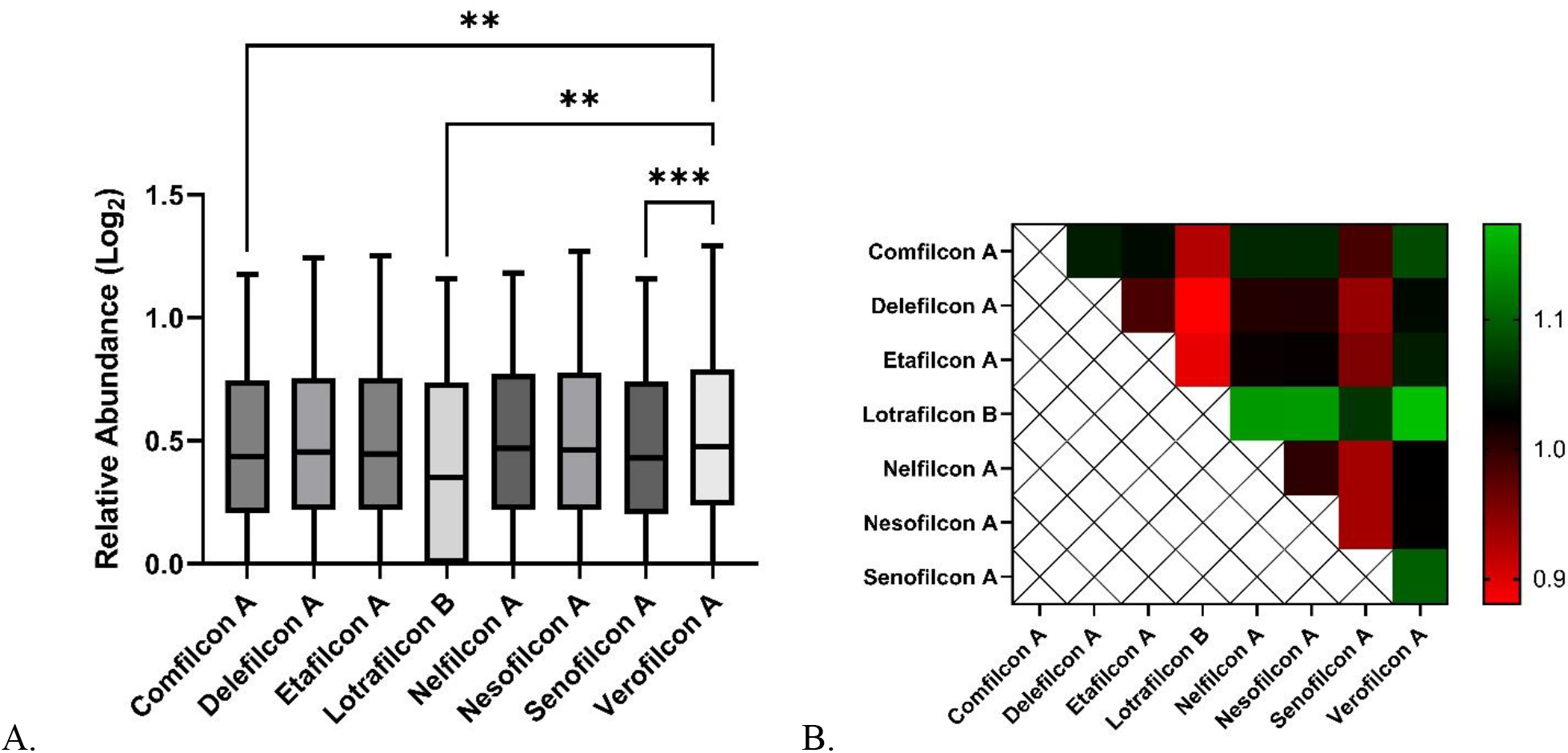
Quantitative protein specificity by SCL material. A) The average protein MS signal intensity, and B) A heat map comparison of average protein abundance between SCL materials. p-values: ** = ≤ 0.01 and *** = ≤ 0.001

In an effort to correlate possible SCL protein biases with protein characteristics, we analyzed various protein characteristics including molecular weight, aromaticity, instability index, gravy, isoelectric point, charge at pH 7.0, and molar extinction coefficient for each lens material. We identified no clear correlations between individual SCL materials and protein collection profile (Supplementary Table 4). We did observe, however, that verofilcon A averaged significantly more protein than many of the other SCL materials (Figure 4). These data support significant differences between total protein yield obtained from different materials, regardless of intersubject variance in tear proteomes.

An ideal sample collection method for MS analysis would collect all proteins equally. This is because large quantitative differences in protein abundance of individual proteins can cause ion suppression in MS and make lower abundant species harder to identify [9, 10]. To identify the SCL material with the most uniform protein collection, we compared the CV of induvial protein abundances between lenses. Our data revealed that comfilcon A and senofilcon A had the lowest quantitative protein variation relative to the other tested SCL materials (Figure 5). We also performed statistical analyses to identify proteins captured at different rates between all lenses and found the 11 proteins listed in Table 3.

**Table 3.** Statistically Significant Quantitative Differences in Individual Proteins for All SCL Materials

| **Uniprot** | **Protein** | **<sup>1</sup> S-M** | **<sup>2</sup> p-value** |
| --- | --- | --- | --- |
| P03973 | Antileukoproteinase (SLPI) | 20.84 | 0.008 |
| P61626 | Lysozyme C (LYSC) | 20.16 | 0.010 |
| P12830 | Cadherin-1 (CADH1) | 17.82 | 0.023 |
| P34096 | Ribonuclease 4 (RNAS4) | 17.64 | 0.024 |
| Q7Z5P9 | Mucin-19 (MUC19) | 17.52 | 0.025 |
| Q6UXB2 | C-X-C motif chemokine 17 (CXL17) | 16.81 | 0.032 |
| P30838 | Aldehyde dehydrogenase, dimeric NADP-preferring (AL3A1) | 16.61 | 0.034 |
| P83731 | 60S ribosomal protein L24 (RL24) | 16.39 | 0.037 |
| P02788 | Lactotransferrin (TRFL) | 16.30 | 0.038 |
| P01034 | Cystatin-C (CYTC) | 16.04 | 0.042 |
| P06744 | Glucose-6-phosphate isomerase (G6PI) | 16.04 | 0.042 |

<sup>1</sup> Skillings-Mack statistic
<sup>2</sup> Un-adjusted p-values

**Figure 5.**
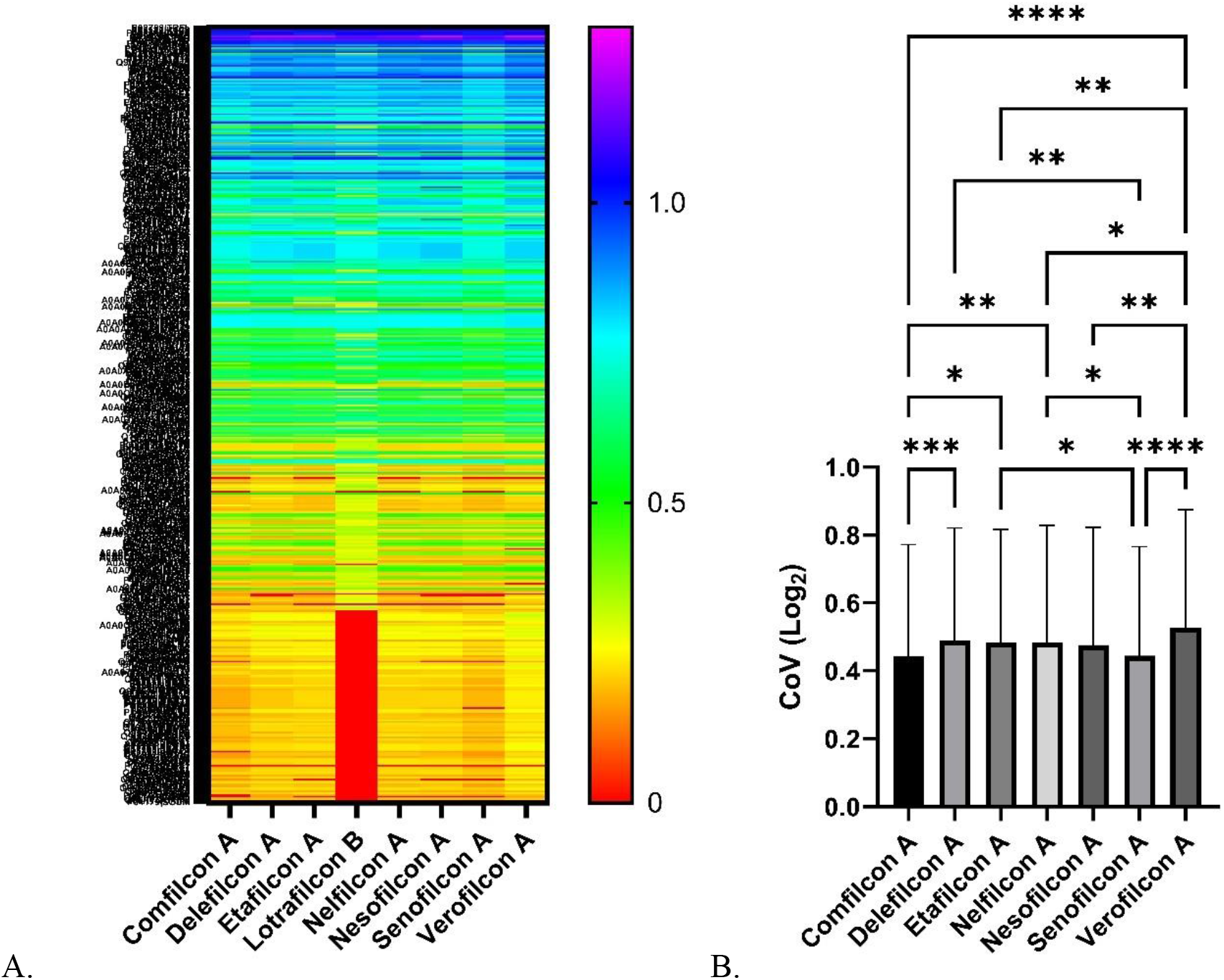
Quantitative protein variation between lenses as measured by the coefficient of variation (CV). A) Heat map of variation by lens material, B) Average CV for all identified proteins by lens material. p-values: * = ≤ 0.05, ** = ≤ 0.01, *** = ≤ 0.001, and **** = ≤ 0.0001.

As etafilcon A and verofilcon A were both unique for not having polymer contamination with FASP, we also compared quantitative changes in individual protein capture between these two lens materials, though no significant differences were observed (Supplementary Table 5). Ironically, verofilcon A lenses use proprietary technology (SMARTSURFACE™) to coat the surface of its lenses to decrease tear protein adsorption [11, 12] but still proved highly effective for SCL sampling based on our pre-defined criteria.

Upon visualizing preferential protein binding for various proteins, we hypothesized that SCL tear sampling could also be advantageous for intra-experiment tear sample normalization. To normalize our experiment using SCL sampled proteins, we determined consistently sampled proteins with low variability and used them to normalize our data set (Table 4). To reduce the effects of normalization compression, we chose to include 5 common and highly abundant tear film protein species: lysozyme C, lactoferrin, lipocalin-1, polymeric immunoglobulin receptor, and clusterin. Importantly, 4 of these 5 proteins were already included in the normalization protein set as they had a CV <0.05. Using this group of proteins to normalize our data, we observed very similar trends to pool normalization within each TMT 10-plex (Figure 6).

**Table 4.**
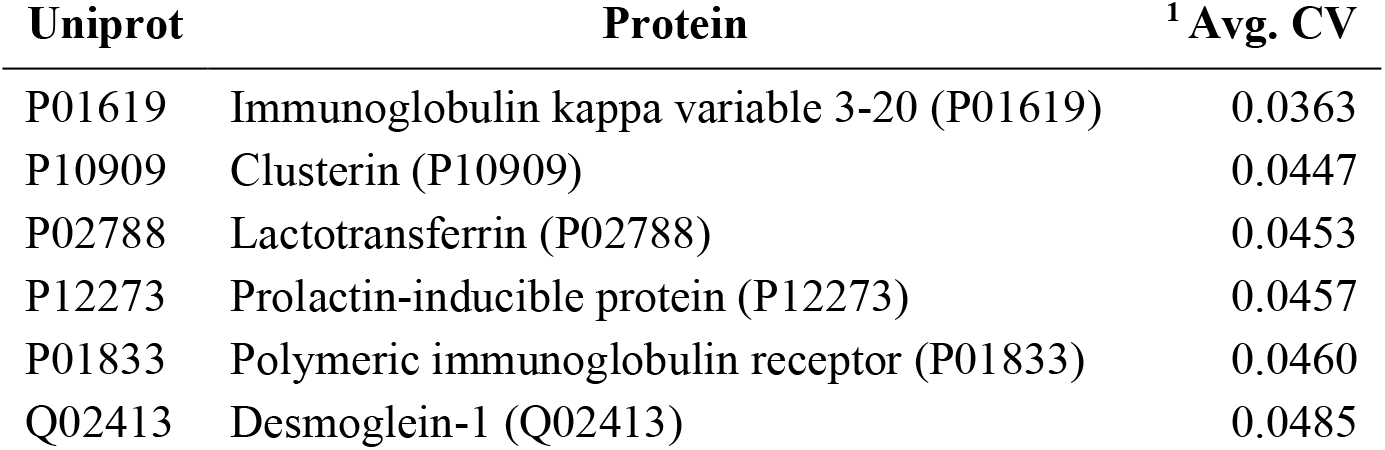
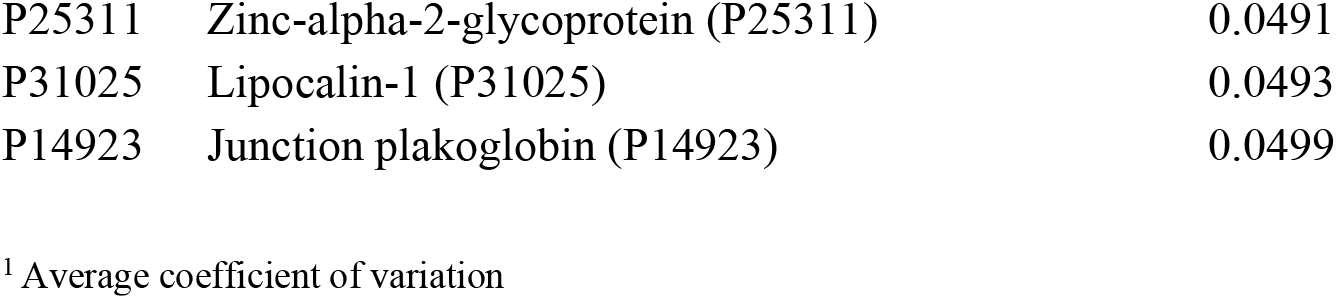
Proteins Sampled by All Lenses with a Coefficient of Variation <0.05

**Figure 6.**
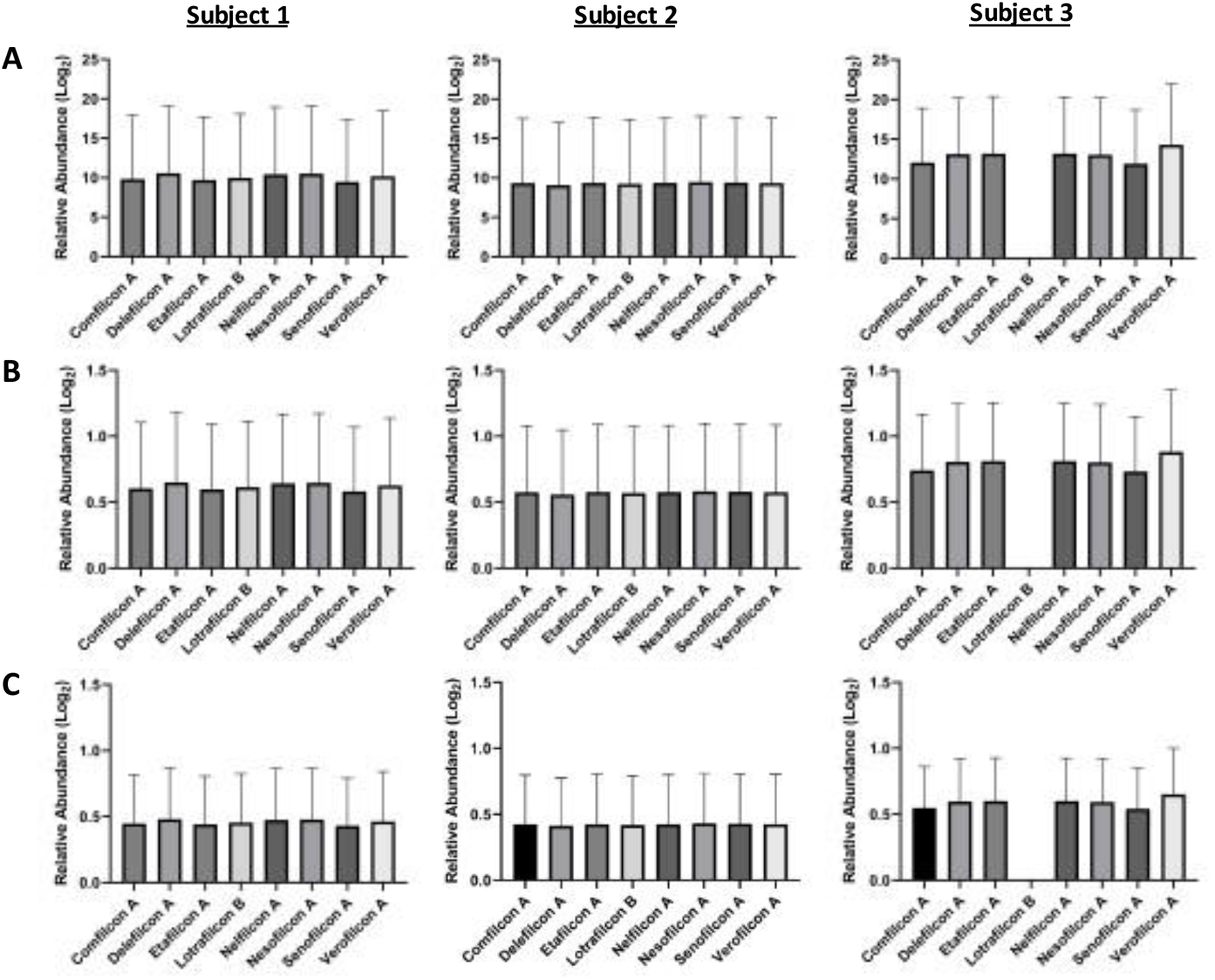
SCL intra-experiment normalization. A) No normalization, B) Normalized to average pool intensity, and C) Normalized to the average intensity of lysozyme C, lactoferrin, lipocalin-1, polymeric immunoglobulin receptor, clusterin, and proteins with a coefficient of variance (CV) <0.05.

Daily SCLs have a significantly lower cost per lens than the alternative monthly modalities (Figure 7). Thus, daily SCLs are a more attractive option for large scale studies and potentially for tear diagnostics. Still, another important consideration is the SCL product lifecycle as SCLs have a limited market lifetime as manufactures replace their products with constantly improving technology. Because Precision 1 (verofilcon A) is at the beginning of its production cycle (released February 2021), it is more likely to remain commercially available for a longer period of time than other SCL materials.

**Figure 7.**
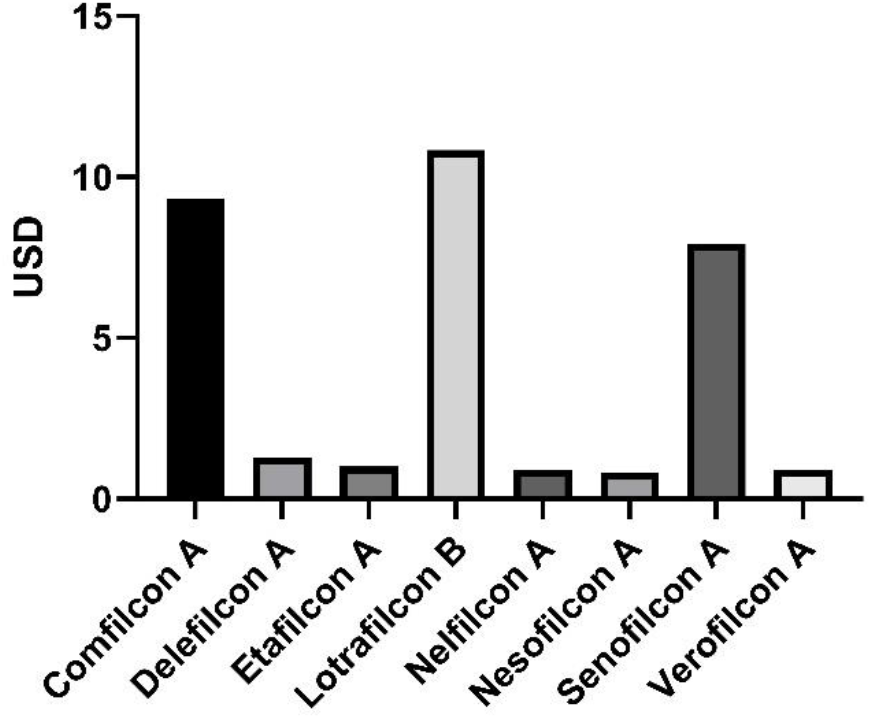
Cost per lens in US dollars. Balafilcon A (Purevision 2, Bausch + Lomb), comfilcon A, (Biofinity, CooperVision), delefilcon A (Dailies Total 1, Alcon), etafilcon A (Acuvue 1-day Moist, Johnson & Johnson Vision), lotrafilcon B (Air Optix Plus Hydraglyde, Alcon), nelfilcon A (DAILIES Aqua Comfort Plus, Alcon), nesofilcon A (Biotrue 1 day, Bausch + Lomb), senofilcon A (Acuvue Oasys, Johnson & Johnson Vision), and verofilcon A (Precision1, Alcon).

After performing all of these tests, we compiled the data and sought to identify the ideal SCL material for tear protein sampling. Given that etafilcon A and verofilcon A materials do not introduce contamination with either FASP or S-trap MS prep protocols, they provide researchers greater flexibility in how they wish to prep the samples. Verofilcon A had significantly higher protein levels relative to several SCL materials (including etafilcon A) as measured by MS. However, comfilcon A and senofilcon A had significantly lower protein variation relative to other lenses. Etafilcon A also had a significant reduction in overall CV compared to verofilcon A. It is also important to consider that verofilcon A (Precision 1, Alcon) are new to the SCL market, and thus have a longer foreseeable production cycle than etafilcon A lenses, which have been available for over 15 years. Daily lenses, such as etafilcon A and verofilcon A, are considerably less expensive than monthly brands, making them a more affordable option. In the end, these data lead us to conclude that both etafilcon A and verofilcon A lenses are optimal for SCL tear protein sampling.

## Discussion

In this study we evaluated 8 SCL materials for use in tear protein sampling studies. Our experiments confirm our hypothesis that the quantity of different proteins captured varies between SCL materials. The choice of SCL material also influences which MS sample prep method can be used for each material to avoid polymer contamination in MS. Understanding these differences will be important in experimental study design and potential diagnostic use moving forward.

A goal of these experiments was to identify a universal sample preparation method for downstream MS analysis of SCL sampled tear film. We compared two well-established MS proteomic sample preparation methods (FASP and S-Trap) [13]. While S-trap has a significantly faster preparation time than FASP, both methods have shown higher protein identifications depending on the sample used [13-15]. In our hands, the highest protein IDs with SCL sampling have been with FASP MS sample prep using 4M guanidine, yielding 328 ± 38 proteins for 6 human subjects [2], though PBS protein removal and S-trap yielded similar results (300 ± 51, delefilcon A). Importantly, significant polymer contamination was observed by MS from samples processed using FASP, but was not observed in same subject samples processed using S-trap.

This study highlights the large variability that exists between individual tear proteomes. Both the total protein capture and protein profile varied substantially between subjects. Importantly, low abundance biomarkers could potentially be masked in patient samples with low total protein capture. Sample normalization is a commonly used to compensate for differences in variable total protein. Our method used a two-pronged approach to normalize between subjects. First, we normalized MS sample loading for total protein capture between subjects by using concentration measurements to ensure that 22 ug total protein was loaded for each sample. However, total protein concentration levels can be skewed by a high or low concentration of only a few highly abundant proteins. As a result, we also performed an internal normalization of the MS data that took advantage of the proteins that were consistently captured by SCLs for all individual subjects tested. This approach avoids the need to use spike proteins by selecting proteins that are consistently identified between subjects with low quantitative variability. Furthermore, it helps account for inter-subject variation in tear volume and protein concentrations, variables that can be significant. Pooling samples for normalization may also be challenging given the low sample volumes collected with tear sampling (often 2 uL or less [16]).

This study has several important limitations. Because it was a pilot study designed to determine if there were large differences between SCL materials for MS proteomics, it had a small number of subjects. To better understand the details of qualitative and quantitative changes between SCL types going forward, more subjects are needed to increase statistical confidence. Second, this is not a comprehensive study as not every SCL material was tested. There may be important pros and cons to other lenses not assessed in this study. Third, consistent with our previous work, this study demonstrates that different combinations of SCL and protein removal chemistry choices have specific advantages and disadvantages. Hence, given the number of variables, further studies will be required to optimize our protocol for SCL tear sampling.

These experiments underscore the fact that SCL materials are significantly different, and that not every SCL can or should be used for tear sampling. Of all the materials tested, etafilcon A and verofilcon A lenses are compatible with FASP, while all tested lenses may be used with the S-Trap protocol. However, we note that SCL design and availability is constantly changing as older lenses are discontinued and newer materials take their place [5, 17, 18]. While etafilcon A and verofilcon A lenses are functionally similar choices, verofilcon A is a much newer lens while etafilcon A has been on the market for over 15 years. However, etafilcon A showed less proteomic variability relative to verofilcon A lenses. Thus, we identified etafilcon A and verofilcon A lenses as optimal choices for tear protein sampling. Continued efforts to understand proteomic specificity will be important as SCL chemistry evolves.

## Data Availability

All data produced are available online at Mendeley Data.

https://data.mendeley.com/datasets/dm3mj729t6/draft?a=05fff7b4-4a6f-4f2b-bcc1-72ffbcb6f16e

## Statements and Declarations

## Acknowledgements

We would like to thank Dr. Steven Weaver and Dr. Carlan Reese at AVC.

