## Supplementary Materials for "Proteomic Specificity of Soft Contact Lenses for Tear Protein Sampling"


### Supplementary Material

Comfilcon A

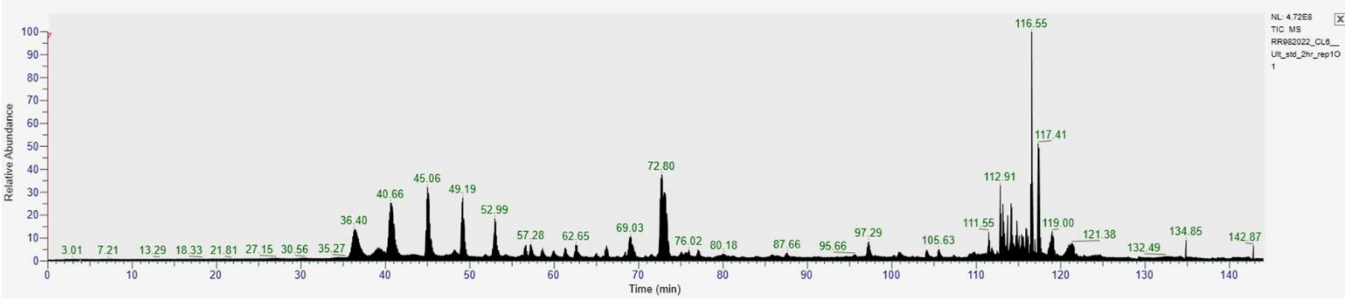


Delefilcon A

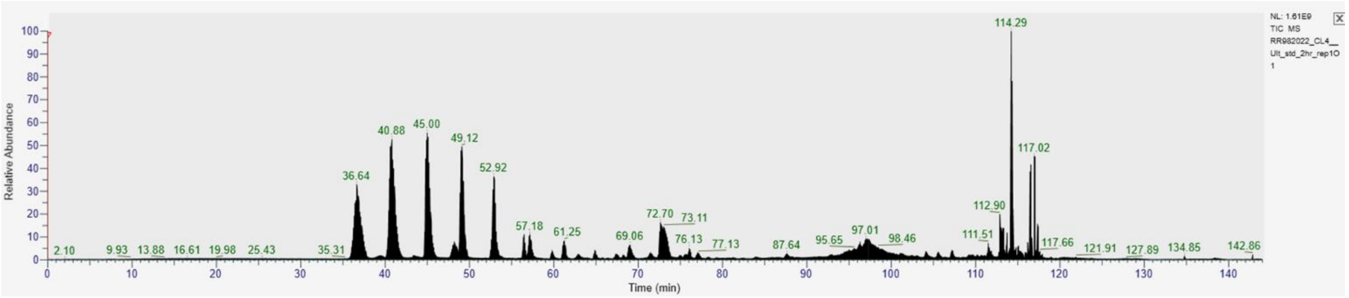


Etafilcon A

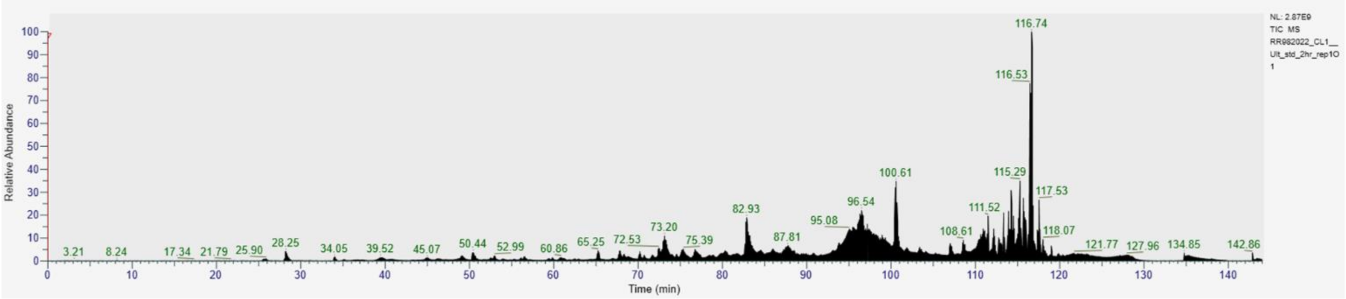


Lotrafilcon B

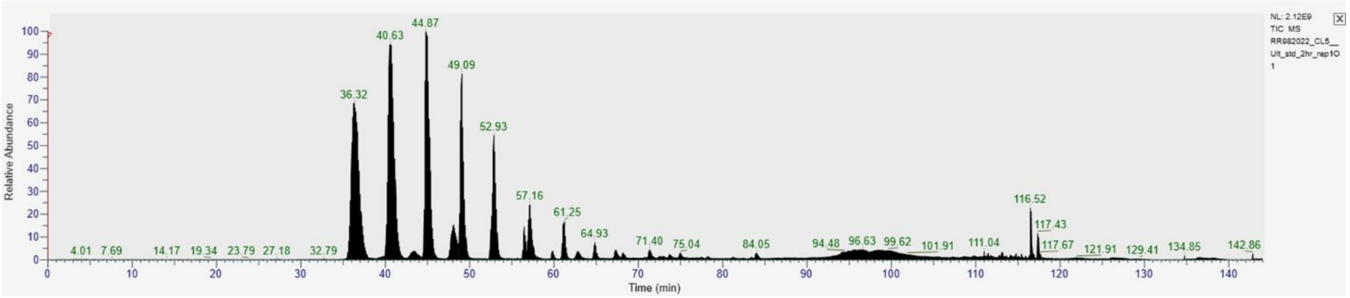


Nelfilcon A

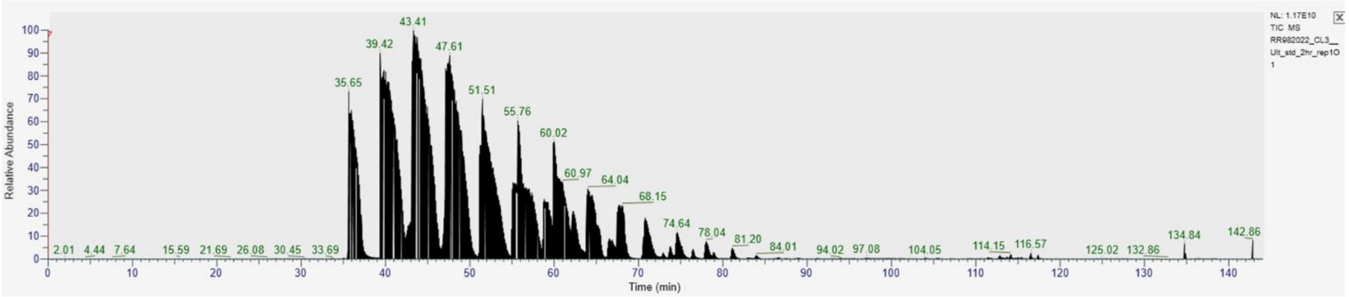


Nesofilcon A

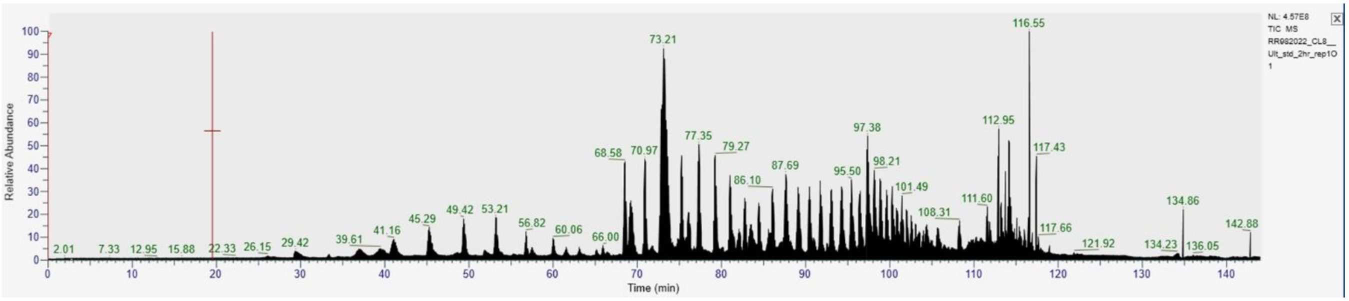


Senofilcon A

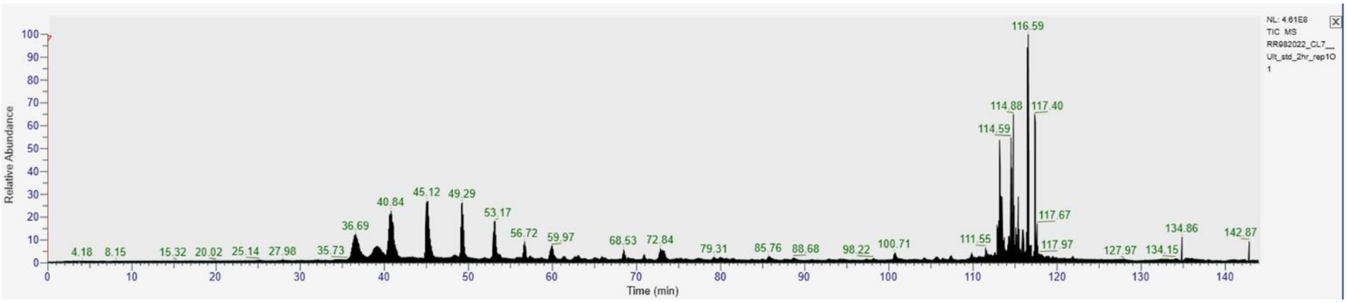


Verofilcon A

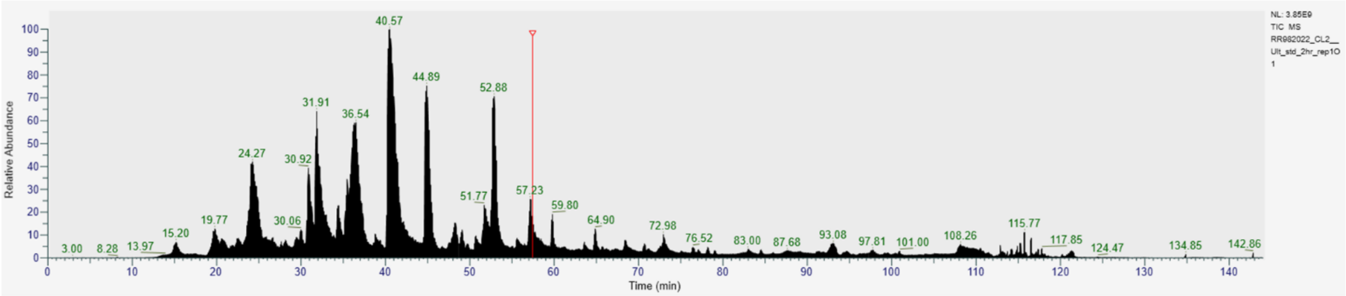


**Supplementary Figure 1.** Polymer contamination in MS by SCL material using the FASP MS sample preparation method. Of the materials tested, only Etafilcon A and Verofilcon A did not show signs of contamination.

#### Supplementary Table 1

*Shapiro-Wilk Data Normality Tests for Proteins Sampled by SCL and Protein Characteristics Datasets*

| Dataset | [1] S-W p-value | Normal Distribution? |
|---|---|---|
| Balafilcon A | 1.72706E-12 | No |
| Comfilcon A | 2.62635E-16 | No |
| Delefilcon A | 7.52941E-17 | No |
| Etafilcon A | 1.89763E-16 | No |
| Lotrafilcon B | 2.84636E-16 | No |
| Nelfilcon A | 1.82684E-16 | No |
| Nesofilcon A | 2.20486E-16 | No |
| Senofilcon A | 5.115E-16 | No |

| | | |
|---|---|---|
| Verofilcon A | 2.72331E-16 | No |
| Aromaticity | 0.226054954 | Yes |
| Charge at pH | 7.62771E-25 | No |
| Gravy | 1.87072E-09 | No |
| Instability index | 1.03526E-06 | No |
| Isoelectric point | 4.01664E-14 | No |
| Molar extinction coefficient | 9.62623E-32 | No |
| Molar extinction coefficient reduced | 1.24443E-31 | No |
| Molecular weight | 2.12377E-34 | No |
| Percent content A | 3.89388E-19 | No |
| Percent content C | 1.42923E-23 | No |
| Percent content D | 1.95603E-12 | No |
| Percent content E | 4.79255E-15 | No |
| Percent content F | 3.44806E-08 | No |
| Percent content G | 5.08233E-21 | No |
| Percent content H | 1.60184E-20 | No |
| Percent content I | 0.01387529 | No |
| Percent content K | 1.71234E-18 | No |
| Percent content L | 6.83285E-05 | No |
| Percent content M | 1.092E-12 | No |
| Percent content N | 6.09775E-06 | No |
| Percent content P | 4.43008E-19 | No |
| Percent content Q | 3.87858E-07 | No |
| Percent content R | 4.03475E-08 | No |
| Percent content S | 1.11733E-16 | No |
| Percent content T | 1.55291E-18 | No |
| Percent content V | 0.065682689 | Yes |
| Percent content W | 3.41721E-16 | No |
| Percent content Y | 0.000454209 | No |
| Sequence length | 7.79273E-35 | No |

[1] Shapiro-Wilk p-value

**Supplementary Table 2**

*Protein Identifications by SCL Material*

| Comfilcon A | Delefilcon A | Etafilcon A | Lotrafilcon B | Nelfilcon A | Nesofilcon A | Senofilcon A | Verofilcon A |
|---|---|---|---|---|---|---|---|
| P02788 | P02788 | P02788 | P02788 | P02788 | P02788 | P02788 | P02788 |
| P35527 | P35527 | P35527 | P35527 | P35527 | P35527 | P35527 | P35527 |
| P04264 | P04264 | P04264 | P04264 | P04264 | P04264 | P04264 | P04264 |
| P13645 | P13645 | P13645 | P13645 | P13645 | P13645 | P13645 | P13645 |
| P35908 | P35908 | P35908 | P35908 | P35908 | P35908 | P35908 | P35908 |
| P61626 | P61626 | P61626 | P61626 | P61626 | P61626 | P61626 | P61626 |
| P02768 | P02768 | P02768 | P02768 | P02768 | P02768 | P02768 | P02768 |
| P31025 | P31025 | P31025 | P31025 | P31025 | P31025 | P31025 | P31025 |
| P01833 | P01833 | P01833 | P01833 | P01833 | P01833 | P01833 | P01833 |
| P01876 | P01876 | P01876 | P01876 | P01876 | P01876 | P01876 | P01876 |
| P01024 | P01024 | P01024 | P01024 | P01024 | P01024 | P01024 | P01024 |
| P25311 | P25311 | P25311 | P25311 | P25311 | P25311 | P25311 | P25311 |
| P04259 | P04259 | P04259 | P04259 | P04259 | P04259 | P04259 | P04259 |
| P19013 | P19013 | P19013 | P19013 | P19013 | P19013 | P19013 | P19013 |
| P13647 | P13647 | P13647 | P13647 | P13647 | P13647 | P13647 | P13647 |
| P02538 | P02538 | P02538 | P02538 | P02538 | P02538 | P02538 | P02538 |
| P08727 | P08727 | P08727 | P08727 | P08727 | P08727 | P08727 | P08727 |
| P13646 | P13646 | P13646 | P13646 | P13646 | P13646 | P13646 | P13646 |
| P08779 | P08779 | P08779 | P08779 | P08779 | P08779 | P08779 | P08779 |
| P02533 | P02533 | P02533 | P02533 | P02533 | P02533 | P02533 | P02533 |
| Q9UGM3 | Q9UGM3 | Q9UGM3 | Q9UGM3 | Q9UGM3 | Q9UGM3 | Q9UGM3 | Q9UGM3 |
| P02787 | P02787 | P02787 | P02787 | P02787 | P02787 | P02787 | P02787 |
| P0DOX7 | P0DOX7 | P0DOX7 | P0DOX7 | P0DOX7 | P0DOX7 | P0DOX7 | P0DOX7 |
| P98160 | P98160 | P98160 | P98160 | P98160 | P98160 | P98160 | P98160 |
| P60709 | P60709 | P60709 | P60709 | P60709 | P60709 | P60709 | P60709 |
| P63261 | P63261 | P63261 | P63261 | P63261 | P63261 | P63261 | P63261 |

| | | | | | | | |
|---|---|---|---|---|---|---|---|
| P15924 | P15924 | P15924 | P15924 | P15924 | P15924 | P15924 | P15924 |
| P06733 | P06733 | P06733 | P06733 | P06733 | P06733 | P06733 | P06733 |
| P01036 | P01036 | P01036 | P01036 | P01036 | P01036 | P01036 | P01036 |
| P12273 | P12273 | P12273 | P12273 | P12273 | P12273 | P12273 | P12273 |
| Q13421 | Q13421 | Q13421 | Q13421 | Q13421 | Q13421 | Q13421 | Q13421 |
| P01037 | P01037 | P01037 | P01037 | P01037 | P01037 | P01037 | P01037 |
| P0DOX2 | P0DOX2 | P0DOX2 | P0DOX2 | P0DOX2 | P0DOX2 | P0DOX2 | P0DOX2 |
| P07355 | P07355 | P07355 | P07355 | P07355 | P07355 | P07355 | P07355 |
| P00450 | P00450 | P00450 | P00450 | P00450 | P00450 | P00450 | P00450 |
| P06396 | P06396 | P06396 | P06396 | P06396 | P06396 | P06396 | P06396 |
| P00738 | P00738 | P00738 | P00738 | P00738 | P00738 | P00738 | P00738 |
| P0DOY2 | P0DOY2 | P0DOY2 | P0DOY2 | P0DOY2 | P0DOY2 | P0DOY2 | P0DOY2 |
| B9A064 | B9A064 | B9A064 | B9A064 | B9A064 | B9A064 | B9A064 | B9A064 |
| P0DOX8 | P0DOX8 | P0DOX8 | P0DOX8 | P0DOX8 | P0DOX8 | P0DOX8 | P0DOX8 |
| P0DOX5 | P0DOX5 | P0DOX5 | P0DOX5 | P0DOX5 | P0DOX5 | P0DOX5 | P0DOX5 |
| P10909 | P10909 | P10909 | P10909 | P10909 | P10909 | P10909 | P10909 |
| P08729 | P08729 | P08729 | P08729 | P08729 | P08729 | P08729 | P08729 |
| P04083 | P04083 | P04083 | P04083 | P04083 | P04083 | P04083 | P04083 |
| P60174 | P60174 | P60174 | P60174 | P60174 | P60174 | P60174 | P60174 |
| P14923 | P14923 | P14923 | P14923 | P14923 | P14923 | P14923 | P14923 |
| P00352 | P00352 | P00352 | P00352 | P00352 | P00352 | P00352 | P00352 |
| P01871 | P01871 | P01871 | P01871 | P01871 | P01871 | P01871 | P01871 |
| P14618 | P14618 | P14618 | P14618 | P14618 | P14618 | P14618 | P14618 |
| P05787 | P05787 | P05787 | P05787 | P05787 | P05787 | P05787 | P05787 |
| O75556 | O75556 | O75556 | O75556 | O75556 | O75556 | O75556 | O75556 |
| P30838 | P30838 | P30838 | P30838 | P30838 | P30838 | P30838 | P30838 |
| P0DMV8 | P0DMV8 | P0DMV8 | P0DMV8 | P0DMV8 | P0DMV8 | P0DMV8 | P0DMV8 |
| P0DMV9 | P0DMV9 | P0DMV9 | P0DMV9 | P0DMV9 | P0DMV9 | P0DMV9 | P0DMV9 |
| Q08380 | Q08380 | Q08380 | Q08380 | Q08380 | Q08380 | Q08380 | Q08380 |
| Q16378 | Q16378 | Q16378 | Q16378 | Q16378 | Q16378 | Q16378 | Q16378 |
| Q04695 | Q04695 | Q04695 | Q04695 | Q04695 | Q04695 | Q04695 | Q04695 |
| P68133 | P68133 | P68133 | P68133 | P68133 | P68133 | P68133 | P68133 |
| P68032 | P68032 | P68032 | P68032 | P68032 | P68032 | P68032 | P68032 |

| | | | | | | | |
|---|---|---|---|---|---|---|---|
| P04792 | P04792 | P04792 | P04792 | P04792 | P04792 | P04792 | P04792 |
| P01009 | P01009 | P01009 | P01009 | P01009 | P01009 | P01009 | P01009 |
| P12035 | P12035 | P12035 | P12035 | P12035 | P12035 | P12035 | P12035 |
| P06702 | P06702 | P06702 | P06702 | P06702 | P06702 | P06702 | P06702 |
| P22079 | P22079 | P22079 | P22079 | P22079 | P22079 | P22079 | P22079 |
| P01859 | P01859 | P01859 | P01859 | P01859 | P01859 | P01859 | P01859 |
| P20061 | P20061 | P20061 | P20061 | P20061 | P20061 | P20061 | P20061 |
| Q02413 | Q02413 | Q02413 | Q02413 | Q02413 | Q02413 | Q02413 | Q02413 |
| P19012 | P19012 | P19012 | P19012 | P19012 | P19012 | P19012 | P19012 |
| P30740 | P30740 | P30740 | P30740 | P30740 | P30740 | P30740 | P30740 |
| P80188 | P80188 | P80188 | P80188 | P80188 | P80188 | P80188 | P80188 |
| Q99935 | Q99935 | Q99935 | Q99935 | Q99935 | Q99935 | Q99935 | Q99935 |
| P04406 | P04406 | P04406 | P04406 | P04406 | P04406 | P04406 | P04406 |
| P03973 | P03973 | P03973 | P03973 | P03973 | P03973 | P03973 | P03973 |
| P98088 | P98088 | P98088 | P98088 | P98088 | P98088 | P98088 | P98088 |
| P14555 | P14555 | P14555 | P14555 | P14555 | P14555 | P14555 | P14555 |
| P01591 | P01591 | P01591 | P01591 | P01591 | P01591 | P01591 | P01591 |
| O95968 | O95968 | O95968 | O95968 | O95968 | O95968 | O95968 | O95968 |
| Q7Z794 | Q7Z794 | Q7Z794 | Q7Z794 | Q7Z794 | Q7Z794 | Q7Z794 | Q7Z794 |
| Q8N1N4 | Q8N1N4 | Q8N1N4 | Q8N1N4 | Q8N1N4 | Q8N1N4 | Q8N1N4 | Q8N1N4 |
| P63104 | P63104 | P63104 | P63104 | P63104 | P63104 | P63104 | P63104 |
| P31944 | P31944 | P31944 | P31944 | P31944 | P31944 | P31944 | P31944 |
| Q96P63 | Q96P63 | Q96P63 | Q96P63 | Q96P63 | Q96P63 | Q96P63 | Q96P63 |
| P09228 | P09228 | P09228 | P09228 | P09228 | P09228 | P09228 | P09228 |
| P09211 | P09211 | P09211 | P09211 | P09211 | P09211 | P09211 | P09211 |
| Q9GZZ8 | Q9GZZ8 | Q9GZZ8 | Q9GZZ8 | Q9GZZ8 | Q9GZZ8 | Q9GZZ8 | Q9GZZ8 |
| P02647 | P02647 | P02647 | P02647 | P02647 | P02647 | P02647 | P02647 |
| Q96DA0 | Q96DA0 | Q96DA0 | Q96DA0 | Q96DA0 | Q96DA0 | Q96DA0 | Q96DA0 |
| P05109 | P05109 | P05109 | P05109 | P05109 | P05109 | P05109 | P05109 |
| P07602 | P07602 | P07602 | P07602 | P07602 | P07602 | P07602 | P07602 |
| P05090 | P05090 | P05090 | P05090 | P05090 | P05090 | P05090 | P05090 |
| P11142 | P11142 | P11142 | P11142 | P11142 | P11142 | P11142 | P11142 |
| Q5D862 | Q5D862 | Q5D862 | Q5D862 | Q5D862 | Q5D862 | Q5D862 | Q5D862 |

| | | | | | | | |
|---|---|---|---|---|---|---|---|
| P61769 | P61769 | P61769 | P61769 | P61769 | P61769 | P61769 | P61769 |
| Q01469 | Q01469 | Q01469 | Q01469 | Q01469 | Q01469 | Q01469 | Q01469 |
| P34096 | P34096 | P34096 | P34096 | P34096 | P34096 | P34096 | P34096 |
| P07339 | P07339 | P07339 | P07339 | P07339 | P07339 | P07339 | P07339 |
| P30044 | P30044 | P30044 | P30044 | P30044 | P30044 | P30044 | P30044 |
| P04075 | P04075 | P04075 | P04075 | P04075 | P04075 | P04075 | P04075 |
| P31947 | P31947 | P31947 | P31947 | P31947 | P31947 | P31947 | P31947 |
| Q5VTE0 | Q5VTE0 | Q5VTE0 | Q5VTE0 | Q5VTE0 | Q5VTE0 | Q5VTE0 | Q5VTE0 |
| P68104 | P68104 | P68104 | P68104 | P68104 | P68104 | P68104 | P68104 |
| P62937 | P62937 | P62937 | P62937 | P62937 | P62937 | P62937 | P62937 |
| P62805 | P62805 | P62805 | P62805 | P62805 | P62805 | P62805 | P62805 |
| P07737 | P07737 | P07737 | P07737 | P07737 | P07737 | P07737 | P07737 |
| A0M8Q6 | A0M8Q6 | A0M8Q6 | A0M8Q6 | A0M8Q6 | A0M8Q6 | A0M8Q6 | A0M8Q6 |
| Q99456 | Q99456 | Q99456 | Q99456 | Q99456 | Q99456 | Q99456 | Q99456 |
| Q06830 | Q06830 | Q06830 | Q06830 | Q06830 | Q06830 | Q06830 | Q06830 |
| P30086 | P30086 | P30086 | P30086 | P30086 | P30086 | P30086 | P30086 |
| | Q3SY84 | Q3SY84 | Q3SY84 | Q3SY84 | Q3SY84 | Q3SY84 | Q3SY84 |
| P02545 | P02545 | P02545 | P02545 | P02545 | P02545 | P02545 | P02545 |
| P00338 | P00338 | P00338 | P00338 | P00338 | P00338 | P00338 | P00338 |
| P01034 | P01034 | P01034 | P01034 | P01034 | P01034 | P01034 | P01034 |
| P68363 | P68363 | P68363 | P68363 | P68363 | P68363 | P68363 | P68363 |
| P52209 | P52209 | P52209 | P52209 | P52209 | P52209 | P52209 | P52209 |
| Q8N474 | Q8N474 | Q8N474 | Q8N474 | Q8N474 | Q8N474 | Q8N474 | Q8N474 |
| P01011 | P01011 | P01011 | P01011 | P01011 | P01011 | P01011 | P01011 |
| P17931 | P17931 | P17931 | P17931 | P17931 | P17931 | P17931 | P17931 |
| P01619 | P01619 | P01619 | P01619 | P01619 | P01619 | P01619 | P01619 |
| P11021 | P11021 | P11021 | P11021 | P11021 | P11021 | P11021 | P11021 |
| P00558 | P00558 | P00558 | P00558 | P00558 | P00558 | P00558 | P00558 |
| P07858 | P07858 | P07858 | P07858 | P07858 | P07858 | P07858 | P07858 |
| P29401 | P29401 | P29401 | P29401 | P29401 | P29401 | P29401 | P29401 |
| P80303 | P80303 | P80303 | P80303 | P80303 | P80303 | P80303 | P80303 |
| P15311 | P15311 | P15311 | P15311 | P15311 | P15311 | P15311 | P15311 |
| Q08554 | Q08554 | Q08554 | Q08554 | Q08554 | Q08554 | Q08554 | Q08554 |

| | | | | | | | |
|---|---|---|---|---|---|---|---|
| Q93079 | Q93079 | Q93079 | Q93079 | Q93079 | Q93079 | Q93079 | Q93079 |
| Q5QNW6 | Q5QNW6 | Q5QNW6 | Q5QNW6 | Q5QNW6 | Q5QNW6 | Q5QNW6 | Q5QNW6 |
| Q99877 | Q99877 | Q99877 | Q99877 | Q99877 | Q99877 | Q99877 | Q99877 |
| O60814 | O60814 | O60814 | O60814 | O60814 | O60814 | O60814 | O60814 |
| P62807 | P62807 | P62807 | P62807 | P62807 | P62807 | P62807 | P62807 |
| P58876 | P58876 | P58876 | P58876 | P58876 | P58876 | P58876 | P58876 |
| Q99880 | Q99880 | Q99880 | Q99880 | Q99880 | Q99880 | Q99880 | Q99880 |
| P57053 | P57053 | P57053 | P57053 | P57053 | P57053 | P57053 | P57053 |
| Q99879 | Q99879 | Q99879 | Q99879 | Q99879 | Q99879 | Q99879 | Q99879 |
| P27797 | P27797 | P27797 | P27797 | P27797 | P27797 | P27797 | P27797 |
| Q6UXB2 | Q6UXB2 | Q6UXB2 | Q6UXB2 | Q6UXB2 | Q6UXB2 | Q6UXB2 | Q6UXB2 |
| A0A0B4J1X5 | A0A0B4J1X5 | A0A0B4J1X5 | A0A0B4J1X5 | A0A0B4J1X5 | A0A0B4J1X5 | A0A0B4J1X5 | A0A0B4J1X5 |
| P06744 | P06744 | P06744 | P06744 | P06744 | P06744 | P06744 | P06744 |
| P05783 | P05783 | P05783 | P05783 | P05783 | P05783 | P05783 | P05783 |
| P01040 | P01040 | P01040 | P01040 | P01040 | P01040 | P01040 | P01040 |
| P40394 | P40394 | P40394 | P40394 | P40394 | P40394 | P40394 | P40394 |
| A0A0B4J1V0 | A0A0B4J1V0 | A0A0B4J1V0 | A0A0B4J1V0 | A0A0B4J1V0 | A0A0B4J1V0 | A0A0B4J1V0 | A0A0B4J1V0 |
| P80748 | P80748 | P80748 | P80748 | P80748 | P80748 | P80748 | P80748 |
| P62979 | P62979 | P62979 | P62979 | P62979 | P62979 | P62979 | P62979 |
| P62987 | P62987 | P62987 | P62987 | P62987 | P62987 | P62987 | P62987 |
| P0CG47 | P0CG47 | P0CG47 | P0CG47 | P0CG47 | P0CG47 | P0CG47 | P0CG47 |
| P0CG48 | P0CG48 | P0CG48 | P0CG48 | P0CG48 | P0CG48 | P0CG48 | P0CG48 |
| P16403 | P16403 | P16403 | P16403 | P16403 | P16403 | P16403 | P16403 |
| P27348 | P27348 | P27348 | P27348 | P27348 | P27348 | P27348 | P27348 |
| P07237 | P07237 | P07237 | P07237 | P07237 | P07237 | P07237 | P07237 |
| P00751 | P00751 | P00751 | P00751 | P00751 | P00751 | P00751 | P00751 |
| P06312 | P06312 | P06312 | P06312 | P06312 | P06312 | P06312 | P06312 |
| P02765 | P02765 | P02765 | P02765 | P02765 | P02765 | P02765 | P02765 |
| P10412 | P10412 | P10412 | P10412 | P10412 | P10412 | P10412 | P10412 |
| P31941 | P31941 | P31941 | P31941 | P31941 | P31941 | P31941 | P31941 |
| P55058 | P55058 | P55058 | P55058 | P55058 | P55058 | P55058 | P55058 |
| P01615 | P01615 | P01615 | P01615 | P01615 | P01615 | P01615 | P01615 |
| A0A075B6P5 | A0A075B6P5 | A0A075B6P5 | A0A075B6P5 | A0A075B6P5 | A0A075B6P5 | A0A075B6P5 | A0A075B6P5 |

| | | | | | | | |
|---|---|---|---|---|---|---|---|
| P81605 | P81605 | P81605 | P81605 | P81605 | P81605 | P81605 | P81605 |
| P02774 | P02774 | P02774 | P02774 | P02774 | P02774 | P02774 | P02774 |
| P14550 | P14550 | P14550 | P14550 | P14550 | P14550 | P14550 | P14550 |
| | A0A0B4J1Y9 | A0A0B4J1Y9 | A0A0B4J1Y9 | A0A0B4J1Y9 | A0A0B4J1Y9 | A0A0B4J1Y9 | A0A0B4J1Y9 |
| P32119 | P32119 | P32119 | P32119 | P32119 | P32119 | P32119 | P32119 |
| P26447 | P26447 | P26447 | P26447 | P26447 | P26447 | P26447 | P26447 |
| P31151 | P31151 | P31151 | P31151 | P31151 | P31151 | P31151 | P31151 |
| Q15517 | Q15517 | Q15517 | Q15517 | Q15517 | Q15517 | Q15517 | Q15517 |
| P20930 | P20930 | P20930 | P20930 | P20930 | P20930 | P20930 | P20930 |
| P0DP08 | P0DP08 | P0DP08 | P0DP08 | P0DP08 | P0DP08 | P0DP08 | P0DP08 |
| P01825 | P01825 | P01825 | P01825 | P01825 | P01825 | P01825 | P01825 |
| P0DP07 | P0DP07 | P0DP07 | P0DP07 | P0DP07 | P0DP07 | P0DP07 | P0DP07 |
| A0A0C4DH41 | A0A0C4DH41 | A0A0C4DH41 | A0A0C4DH41 | A0A0C4DH41 | A0A0C4DH41 | A0A0C4DH41 | A0A0C4DH41 |
| P0DP06 | P0DP06 | P0DP06 | P0DP06 | P0DP06 | P0DP06 | P0DP06 | P0DP06 |
| P06331 | P06331 | P06331 | P06331 | P06331 | P06331 | P06331 | P06331 |
| P01824 | P01824 | P01824 | P01824 | P01824 | P01824 | P01824 | P01824 |
| A0A0A0MRZ8 | A0A0A0MRZ8 | A0A0A0MRZ8 | A0A0A0MRZ8 | A0A0A0MRZ8 | A0A0A0MRZ8 | A0A0A0MRZ8 | A0A0A0MRZ8 |
| P04433 | P04433 | P04433 | P04433 | P04433 | P04433 | P04433 | P04433 |
| P31949 | P31949 | P31949 | P31949 | P31949 | P31949 | P31949 | P31949 |
| P60953 | P60953 | P60953 | P60953 | P60953 | P60953 | P60953 | P60953 |
| Q6KB66 | Q6KB66 | Q6KB66 | Q6KB66 | Q6KB66 | Q6KB66 | Q6KB66 | Q6KB66 |
| P10599 | P10599 | P10599 | P10599 | P10599 | P10599 | P10599 | P10599 |
| P40925 | P40925 | P40925 | P40925 | P40925 | P40925 | P40925 | P40925 |
| Q14515 | Q14515 | Q14515 | Q14515 | Q14515 | Q14515 | Q14515 | Q14515 |
| A0A075B6S5 | A0A075B6S5 | A0A075B6S5 | A0A075B6S5 | A0A075B6S5 | A0A075B6S5 | A0A075B6S5 | A0A075B6S5 |
| P01742 | P01742 | P01742 | P01742 | P01742 | P01742 | P01742 | P01742 |
| A0A0C4DH31 | A0A0C4DH31 | A0A0C4DH31 | A0A0C4DH31 | A0A0C4DH31 | A0A0C4DH31 | A0A0C4DH31 | A0A0C4DH31 |
| P23083 | P23083 | P23083 | P23083 | P23083 | P23083 | P23083 | P23083 |
| P07900 | P07900 | P07900 | P07900 | P07900 | P07900 | P07900 | P07900 |
| P84243 | P84243 | P84243 | P84243 | P84243 | P84243 | P84243 | P84243 |
| P01700 | P01700 | P01700 | P01700 | P01700 | P01700 | P01700 | P01700 |
| P23528 | P23528 | P23528 | P23528 | P23528 | P23528 | P23528 | P23528 |
| P21980 | P21980 | P21980 | P21980 | P21980 | P21980 | P21980 | P21980 |

| | | | | | | | |
|---|---|---|---|---|---|---|---|
| Q09666 | Q09666 | Q09666 | Q09666 | Q09666 | Q09666 | Q09666 | Q09666 |
| P01624 | P01624 | P01624 | P01624 | P01624 | P01624 | P01624 | P01624 |
| P07384 | P07384 | P07384 | P07384 | P07384 | P07384 | P07384 | P07384 |
| Q6UWP8 | Q6UWP8 | Q6UWP8 | Q6UWP8 | Q6UWP8 | Q6UWP8 | Q6UWP8 | Q6UWP8 |
| Q8NBJ4 | Q8NBJ4 | Q8NBJ4 | Q8NBJ4 | Q8NBJ4 | Q8NBJ4 | Q8NBJ4 | Q8NBJ4 |
| P40926 | P40926 | P40926 | P40926 | P40926 | P40926 | P40926 | P40926 |
| P68371 | P68371 | P68371 | P68371 | P68371 | P68371 | P68371 | P68371 |
| A0A0A0MS15 | A0A0A0MS15 | A0A0A0MS15 | A0A0A0MS15 | A0A0A0MS15 | A0A0A0MS15 | A0A0A0MS15 | A0A0A0MS15 |
| P01593 | P01593 | P01593 | P01593 | P01593 | P01593 | P01593 | P01593 |
| P01594 | P01594 | P01594 | P01594 | P01594 | P01594 | P01594 | P01594 |
| P02766 | P02766 | P02766 | P02766 | P02766 | P02766 | P02766 | P02766 |
| Q08188 | Q08188 | Q08188 | Q08188 | Q08188 | Q08188 | Q08188 | Q08188 |
| P05089 | P05089 | P05089 | P05089 | P05089 | P05089 | P05089 | P05089 |
| Q9UBT3 | Q9UBT3 | Q9UBT3 | Q9UBT3 | Q9UBT3 | Q9UBT3 | Q9UBT3 | Q9UBT3 |
| O00584 | O00584 | O00584 | O00584 | O00584 | O00584 | O00584 | O00584 |
| | A0A0B4J2D9 | A0A0B4J2D9 | A0A0B4J2D9 | A0A0B4J2D9 | A0A0B4J2D9 | A0A0B4J2D9 | A0A0B4J2D9 |
| | P0DP09 | P0DP09 | P0DP09 | P0DP09 | P0DP09 | P0DP09 | P0DP09 |
| O75874 | O75874 | O75874 | O75874 | O75874 | O75874 | O75874 | O75874 |
| P15814 | P15814 | P15814 | P15814 | P15814 | P15814 | P15814 | P15814 |
| P30085 | P30085 | P30085 | P30085 | P30085 | P30085 | P30085 | P30085 |
| A0A0C4DH34 | A0A0C4DH34 | A0A0C4DH34 | A0A0C4DH34 | A0A0C4DH34 | A0A0C4DH34 | A0A0C4DH34 | A0A0C4DH34 |
| P30101 | P30101 | P30101 | P30101 | P30101 | P30101 | P30101 | P30101 |
| Q14697 | Q14697 | Q14697 | Q14697 | Q14697 | Q14697 | Q14697 | Q14697 |
| P12830 | P12830 | P12830 | P12830 | P12830 | P12830 | P12830 | P12830 |
| Q16651 | Q16651 | Q16651 | Q16651 | Q16651 | Q16651 | Q16651 | Q16651 |
| P13987 | P13987 | P13987 | P13987 | P13987 | P13987 | P13987 | P13987 |
| O43653 | O43653 | O43653 | O43653 | O43653 | O43653 | O43653 | O43653 |
| P02763 | P02763 | P02763 | P02763 | P02763 | P02763 | P02763 | P02763 |
| A0A0B4J1V6 | A0A0B4J1V6 | A0A0B4J1V6 | A0A0B4J1V6 | A0A0B4J1V6 | A0A0B4J1V6 | A0A0B4J1V6 | A0A0B4J1V6 |
| O43852 | O43852 | O43852 | O43852 | O43852 | O43852 | O43852 | O43852 |
| P08758 | P08758 | P08758 | P08758 | P08758 | P08758 | P08758 | P08758 |
| Q5T749 | Q5T749 | Q5T749 | Q5T749 | Q5T749 | Q5T749 | Q5T749 | Q5T749 |
| Q7Z5P9 | Q7Z5P9 | Q7Z5P9 | Q7Z5P9 | Q7Z5P9 | Q7Z5P9 | Q7Z5P9 | Q7Z5P9 |

| | | | | | | | |
|---|---|---|---|---|---|---|---|
| P01714 | P01714 | P01714 | P01714 | P01714 | P01714 | P01714 | P01714 |
| P01717 | P01717 | P01717 | P01717 | P01717 | P01717 | P01717 | P01717 |
| A0A075B6K4 | A0A075B6K4 | A0A075B6K4 | A0A075B6K4 | A0A075B6K4 | A0A075B6K4 | A0A075B6K4 | A0A075B6K4 |
| P23284 | P23284 | P23284 | P23284 | P23284 | P23284 | P23284 | P23284 |
| P12814 | P12814 | P12814 | P12814 | P12814 | P12814 | P12814 | P12814 |
| | P40199 | P40199 | P40199 | P40199 | P40199 | P40199 | P40199 |
| Q02818 | Q02818 | Q02818 | Q02818 | Q02818 | Q02818 | Q02818 | Q02818 |
| P49788 | P49788 | P49788 | P49788 | P49788 | P49788 | P49788 | P49788 |
| P06703 | P06703 | P06703 | P06703 | P06703 | P06703 | P06703 | P06703 |
| P16401 | P16401 | P16401 | P16401 | P16401 | P16401 | P16401 | P16401 |
| O60437 | O60437 | O60437 | O60437 | O60437 | O60437 | O60437 | O60437 |
| P47929 | P47929 | P47929 | P47929 | P47929 | P47929 | P47929 | P47929 |
| Q14508 | Q14508 | Q14508 | Q14508 | Q14508 | Q14508 | Q14508 | Q14508 |
| | Q13835 | Q13835 | Q13835 | Q13835 | Q13835 | Q13835 | Q13835 |
| P02750 | P02750 | P02750 | P02750 | P02750 | P02750 | P02750 | P02750 |
| P21926 | P21926 | P21926 | P21926 | P21926 | P21926 | P21926 | P21926 |
| | P19961 | P19961 | P19961 | P19961 | P19961 | P19961 | P19961 |
| | P0DTE7 | P0DTE7 | P0DTE7 | P0DTE7 | P0DTE7 | P0DTE7 | P0DTE7 |
| | P04746 | P04746 | P04746 | P04746 | P04746 | P04746 | P04746 |
| | P04745 | P04745 | P04745 | P04745 | P04745 | P04745 | P04745 |
| | P0DTE8 | P0DTE8 | P0DTE8 | P0DTE8 | P0DTE8 | P0DTE8 | P0DTE8 |
| Q9UBC9 | Q9UBC9 | Q9UBC9 | Q9UBC9 | Q9UBC9 | Q9UBC9 | Q9UBC9 | Q9UBC9 |
| Q96QA5 | Q96QA5 | Q96QA5 | Q96QA5 | Q96QA5 | Q96QA5 | Q96QA5 | Q96QA5 |
| | Q13296 | Q13296 | Q13296 | Q13296 | Q13296 | Q13296 | Q13296 |
| | Q14002 | Q14002 | Q14002 | Q14002 | Q14002 | Q14002 | Q14002 |
| P62753 | P62753 | P62753 | P62753 | P62753 | P62753 | P62753 | P62753 |
| P09429 | P09429 | P09429 | P09429 | P09429 | P09429 | P09429 | P09429 |
| P83731 | P83731 | P83731 | P83731 | P83731 | P83731 | P83731 | P83731 |
| Q96S96 | Q96S96 | Q96S96 | Q96S96 | Q96S96 | Q96S96 | Q96S96 | Q96S96 |
| | Q5SNV9 | Q5SNV9 | Q5SNV9 | Q5SNV9 | Q5SNV9 | Q5SNV9 | Q5SNV9 |
| | Q6ZR08 | Q6ZR08 | Q6ZR08 | Q6ZR08 | Q6ZR08 | Q6ZR08 | Q6ZR08 |
| | P07910 | P07910 | P07910 | P07910 | P07910 | P07910 | P07910 |
| | Q9UKZ1 | Q9UKZ1 | Q9UKZ1 | Q9UKZ1 | Q9UKZ1 | Q9UKZ1 | Q9UKZ1 |

| | | | | | | | |
|---|---|---|---|---|---|---|---|
| P20933 | P20933 | P20933 | P20933 | P20933 | P20933 | P20933 | P20933 |
| | Q8IZL8 | Q8IZL8 | Q8IZL8 | Q8IZL8 | Q8IZL8 | Q8IZL8 | Q8IZL8 |
| | P00505 | P00505 | P00505 | P00505 | P00505 | P00505 | P00505 |
| | | P48634 | | P48634 | | P48634 | |
| | Q96G74 | Q96G74 | Q96G74 | Q96G74 | Q96G74 | Q96G74 | Q96G74 |
| | Q8IVF2 | Q8IVF2 | Q8IVF2 | Q8IVF2 | Q8IVF2 | Q8IVF2 | Q8IVF2 |
| | Q8TF72 | Q8TF72 | Q8TF72 | Q8TF72 | Q8TF72 | Q8TF72 | Q8TF72 |
| | Q14966 | Q14966 | Q14966 | Q14966 | Q14966 | Q14966 | Q14966 |
| | Q7LBE3 | Q7LBE3 | Q7LBE3 | Q7LBE3 | Q7LBE3 | Q7LBE3 | Q7LBE3 |
| | A6NKD9 | A6NKD9 | A6NKD9 | A6NKD9 | A6NKD9 | A6NKD9 | A6NKD9 |
| | A0A0B4J1U7 | A0A0B4J1U7 | A0A0B4J1U7 | A0A0B4J1U7 | A0A0B4J1U7 | A0A0B4J1U7 | A0A0B4J1U7 |
| | | Q6PID8 | Q6PID8 | | | Q6PID8 | |
| Q99574 | Q99574 | Q99574 | Q99574 | Q99574 | Q99574 | Q99574 | Q99574 |
| | Q14526 | Q14526 | Q14526 | Q14526 | Q14526 | Q14526 | Q14526 |
| | Q5VSY0 | Q5VSY0 | Q5VSY0 | Q5VSY0 | Q5VSY0 | Q5VSY0 | Q5VSY0 |
| | Q8TCU6 | Q8TCU6 | Q8TCU6 | Q8TCU6 | Q8TCU6 | Q8TCU6 | Q8TCU6 |
| | Q7Z2W7 | Q7Z2W7 | Q7Z2W7 | Q7Z2W7 | Q7Z2W7 | Q7Z2W7 | Q7Z2W7 |
| | Q14106 | Q14106 | Q14106 | Q14106 | Q14106 | Q14106 | Q14106 |
| | Q9GZQ3 | Q9GZQ3 | Q9GZQ3 | Q9GZQ3 | Q9GZQ3 | Q9GZQ3 | Q9GZQ3 |
| | Q8NHQ8 | Q8NHQ8 | Q8NHQ8 | Q8NHQ8 | Q8NHQ8 | Q8NHQ8 | Q8NHQ8 |
| | P01703 | P01703 | P01703 | P01703 | P01703 | P01703 | P01703 |
| | O75882 | O75882 | O75882 | O75882 | O75882 | O75882 | O75882 |
| | O60902 | O60902 | O60902 | O60902 | O60902 | O60902 | O60902 |
| Q5VSP4 | Q5VSP4 | Q5VSP4 | Q5VSP4 | Q5VSP4 | Q5VSP4 | Q5VSP4 | Q5VSP4 |
| P01861 | P01861 | P01861 | P01861 | P01861 | P01861 | P01861 | P01861 |
| O95678 | O95678 | O95678 | O95678 | O95678 | O95678 | O95678 | O95678 |
| P0DOX6 | P0DOX6 | P0DOX6 | P0DOX6 | P0DOX6 | P0DOX6 | P0DOX6 | P0DOX6 |
| Q86YZ3 | Q86YZ3 | Q86YZ3 | Q86YZ3 | Q86YZ3 | Q86YZ3 | Q86YZ3 | Q86YZ3 |
| Q14525 | Q14525 | Q14525 | Q14525 | Q14525 | Q14525 | Q14525 | Q14525 |
| Q562R1 | Q562R1 | Q562R1 | Q562R1 | Q562R1 | Q562R1 | Q562R1 | Q562R1 |
| P31946 | P31946 | P31946 | P31946 | P31946 | P31946 | P31946 | P31946 |
| Q7Z3Y8 | Q7Z3Y8 | Q7Z3Y8 | Q7Z3Y8 | Q7Z3Y8 | Q7Z3Y8 | Q7Z3Y8 | Q7Z3Y8 |
| P01780 | P01780 | P01780 | P01780 | P01780 | P01780 | P01780 | P01780 |

| | | | | | | | |
|---|---|---|---|---|---|---|---|
| A0A0J9YX35 | A0A0J9YX35 | A0A0J9YX35 | A0A0J9YX35 | A0A0J9YX35 | A0A0J9YX35 | A0A0J9YX35 | A0A0J9YX35 |
| A0A0J9YXX1 | A0A0J9YXX1 | A0A0J9YXX1 | A0A0J9YXX1 | A0A0J9YXX1 | A0A0J9YXX1 | A0A0J9YXX1 | A0A0J9YXX1 |
| A0A0C4DH38 | A0A0C4DH38 | A0A0C4DH38 | A0A0C4DH38 | A0A0C4DH38 | A0A0C4DH38 | A0A0C4DH38 | A0A0C4DH38 |
| P28799 | P28799 | P28799 | P28799 | P28799 | P28799 | P28799 | P28799 |
| P01762 | P01762 | P01762 | P01762 | P01762 | P01762 | P01762 | P01762 |
| Q7Z3Y9 | Q7Z3Y9 | Q7Z3Y9 | Q7Z3Y9 | Q7Z3Y9 | Q7Z3Y9 | Q7Z3Y9 | Q7Z3Y9 |
| Q15149 | Q15149 | Q15149 | Q15149 | Q15149 | Q15149 | Q15149 | Q15149 |
| Q7Z406 | Q7Z406 | Q7Z406 | Q7Z406 | Q7Z406 | Q7Z406 | Q7Z406 | Q7Z406 |
| P06731 | P06731 | P06731 | P06731 | P06731 | P06731 | P06731 | P06731 |
| P04632 | P04632 | P04632 | P04632 | P04632 | P04632 | P04632 | P04632 |
| Q07666 | Q07666 | Q07666 | Q07666 | Q07666 | Q07666 | Q07666 | Q07666 |
| | O60716 | O60716 | O60716 | O60716 | O60716 | O60716 | O60716 |
| P01611 | P01611 | P01611 | P01611 | P01611 | P01611 | P01611 | P01611 |
| A0A0C4DH72 | A0A0C4DH72 | A0A0C4DH72 | A0A0C4DH72 | A0A0C4DH72 | A0A0C4DH72 | A0A0C4DH72 | A0A0C4DH72 |
| P08246 | P08246 | P08246 | P08246 | P08246 | P08246 | P08246 | P08246 |
| A0A075B6R9 | A0A075B6R9 | A0A075B6R9 | A0A075B6R9 | A0A075B6R9 | A0A075B6R9 | A0A075B6R9 | A0A075B6R9 |
| A0A0C4DH68 | A0A0C4DH68 | A0A0C4DH68 | A0A0C4DH68 | A0A0C4DH68 | A0A0C4DH68 | A0A0C4DH68 | A0A0C4DH68 |
| P40121 | P40121 | P40121 | P40121 | P40121 | P40121 | P40121 | P40121 |
| | O76021 | O76021 | O76021 | O76021 | O76021 | O76021 | O76021 |
| A0A075B6H7 | A0A075B6H7 | A0A075B6H7 | A0A075B6H7 | A0A075B6H7 | A0A075B6H7 | A0A075B6H7 | A0A075B6H7 |
| P42357 | P42357 | P42357 | P42357 | | P42357 | P42357 | P42357 |
| P13797 | P13797 | P13797 | P13797 | P13797 | P13797 | P13797 | P13797 |
| P04211 | P04211 | P04211 | P04211 | P04211 | P04211 | P04211 | P04211 |
| A0A075B6I9 | A0A075B6I9 | A0A075B6I9 | A0A075B6I9 | A0A075B6I9 | A0A075B6I9 | A0A075B6I9 | A0A075B6I9 |
| P55064 | P55064 | P55064 | P55064 | P55064 | P55064 | P55064 | P55064 |
| P07951 | P07951 | P07951 | P07951 | P07951 | P07951 | P07951 | P07951 |
| P09493 | P09493 | P09493 | P09493 | P09493 | P09493 | P09493 | P09493 |
| P22735 | P22735 | P22735 | P22735 | P22735 | P22735 | P22735 | P22735 |
| P01701 | P01701 | P01701 | P01701 | P01701 | P01701 | P01701 | P01701 |
| P49840 | P49840 | P49840 | P49840 | P49840 | P49840 | P49840 | P49840 |
| P0DOX3 | P0DOX3 | P0DOX3 | P0DOX3 | P0DOX3 | P0DOX3 | P0DOX3 | P0DOX3 |
| | P10809 | P10809 | P10809 | P10809 | P10809 | P10809 | P10809 |
| P02679 | P02679 | P02679 | P02679 | P02679 | P02679 | P02679 | P02679 |

| | | | | | | | |
|---|---|---|---|---|---|---|---|
| Q9UPP2 | Q9UPP2 | Q9UPP2 | Q9UPP2 | Q9UPP2 | Q9UPP2 | Q9UPP2 | Q9UPP2 |
| A0A075B6I0 | A0A075B6I0 | A0A075B6I0 | A0A075B6I0 | A0A075B6I0 | A0A075B6I0 | A0A075B6I0 | A0A075B6I0 |
| Q86V81 | Q86V81 | Q86V81 | Q86V81 | Q86V81 | Q86V81 | Q86V81 | Q86V81 |
| P22681 | P22681 | P22681 | P22681 | P22681 | P22681 | P22681 | P22681 |
| | Q9BVC4 | | | | | | |
| Q9BWS9 | Q9BWS9 | | Q9BWS9 | Q9BWS9 | Q9BWS9 | | |
| Q7RTR2 | Q7RTR2 | Q7RTR2 | Q7RTR2 | Q7RTR2 | Q7RTR2 | Q7RTR2 | Q7RTR2 |
| Q8N6G6 | Q8N6G6 | Q8N6G6 | Q8N6G6 | Q8N6G6 | Q8N6G6 | Q8N6G6 | Q8N6G6 |
| P67809 | P67809 | P67809 | P67809 | P67809 | P67809 | P67809 | P67809 |
| Q8IZ21 | Q8IZ21 | Q8IZ21 | Q8IZ21 | Q8IZ21 | Q8IZ21 | Q8IZ21 | Q8IZ21 |
| | | Q8WXS5 | | | | | |
| Q9ULD2 | Q9ULD2 | Q9ULD2 | Q9ULD2 | Q9ULD2 | Q9ULD2 | Q9ULD2 | Q9ULD2 |
| Q86SM8 | Q86SM8 | Q86SM8 | Q86SM8 | Q86SM8 | Q86SM8 | Q86SM8 | Q86SM8 |
| | Q13283 | Q13283 | Q13283 | Q13283 | Q13283 | Q13283 | Q13283 |
| Q9NSA2 | Q9NSA2 | Q9NSA2 | Q9NSA2 | | Q9NSA2 | Q9NSA2 | Q9NSA2 |
| Q15323 | Q15323 | Q15323 | Q15323 | | Q15323 | Q15323 | Q15323 |
| O43790 | O43790 | O43790 | O43790 | | O43790 | O43790 | O43790 |
| Q14533 | Q14533 | Q14533 | Q14533 | | Q14533 | Q14533 | Q14533 |
| P62258 | P62258 | P62258 | P62258 | | P62258 | P62258 | P62258 |
| Q13228 | Q13228 | Q13228 | Q13228 | | Q13228 | Q13228 | Q13228 |
| P78386 | P78386 | P78386 | P78386 | | P78386 | P78386 | P78386 |
| P13929 | P13929 | P13929 | P13929 | | P13929 | P13929 | P13929 |
| A0A0C4DH42 | A0A0C4DH42 | A0A0C4DH42 | A0A0C4DH42 | | A0A0C4DH42 | A0A0C4DH42 | A0A0C4DH42 |
| P02790 | P02790 | P02790 | P02790 | | P02790 | P02790 | P02790 |
| P22392 | P22392 | P22392 | P22392 | | P22392 | P22392 | P22392 |
| P37802 | P37802 | P37802 | P37802 | | P37802 | P37802 | P37802 |
| P29508 | P29508 | P29508 | P29508 | | P29508 | P29508 | P29508 |
| P18510 | P18510 | P18510 | P18510 | | P18510 | P18510 | P18510 |
| P30041 | P30041 | P30041 | P30041 | | P30041 | P30041 | P30041 |
| P15259 | P15259 | P15259 | P15259 | | P15259 | P15259 | P15259 |
| P18669 | P18669 | P18669 | P18669 | | P18669 | P18669 | P18669 |
| P48594 | P48594 | P48594 | P48594 | | P48594 | P48594 | P48594 |
| P0DP24 | P0DP24 | P0DP24 | P0DP24 | | P0DP24 | P0DP24 | P0DP24 |

| | | | | | | | |
|---|---|---|---|---|---|---|---|
| P0DP23 | P0DP23 | P0DP23 | P0DP23 | | P0DP23 | P0DP23 | P0DP23 |
| P0DP25 | P0DP25 | P0DP25 | P0DP25 | | P0DP25 | P0DP25 | P0DP25 |
| P05387 | P05387 | P05387 | P05387 | | P05387 | P05387 | P05387 |
| P04080 | P04080 | P04080 | P04080 | | P04080 | P04080 | P04080 |
| P22626 | P22626 | P22626 | P22626 | | P22626 | P22626 | P22626 |
| O75223 | O75223 | O75223 | O75223 | | O75223 | O75223 | O75223 |
| P37837 | P37837 | P37837 | P37837 | | P37837 | P37837 | P37837 |
| P27482 | P27482 | P27482 | P27482 | | P27482 | P27482 | P27482 |
| O00299 | O00299 | O00299 | O00299 | | O00299 | O00299 | O00299 |
| P08582 | P08582 | P08582 | P08582 | | P08582 | P08582 | P08582 |
| | | Q9HC84 | Q9HC84 | | Q9HC84 | | |
| P0DME0 | P0DME0 | P0DME0 | P0DME0 | | P0DME0 | P0DME0 | P0DME0 |
| Q01105 | Q01105 | Q01105 | Q01105 | | Q01105 | Q01105 | Q01105 |
| P02511 | P02511 | P02511 | P02511 | | P02511 | P02511 | P02511 |
| P01008 | P01008 | P01008 | P01008 | | P01008 | P01008 | P01008 |
| P08571 | P08571 | P08571 | P08571 | | P08571 | P08571 | P08571 |
| A0A075B6S9 | A0A075B6S9 | A0A075B6S9 | A0A075B6S9 | | A0A075B6S9 | A0A075B6S9 | A0A075B6S9 |
| P0DSN7 | P0DSN7 | P0DSN7 | P0DSN7 | | P0DSN7 | P0DSN7 | P0DSN7 |
| P06454 | P06454 | P06454 | P06454 | | P06454 | P06454 | P06454 |
| P08294 | P08294 | P08294 | P08294 | | P08294 | P08294 | P08294 |
| Q9HAV0 | Q9HAV0 | Q9HAV0 | Q9HAV0 | | Q9HAV0 | Q9HAV0 | Q9HAV0 |
| P62873 | P62873 | P62873 | P62873 | | P62873 | P62873 | P62873 |
| P62879 | P62879 | P62879 | P62879 | | P62879 | P62879 | P62879 |
| P16520 | P16520 | P16520 | P16520 | | P16520 | P16520 | P16520 |
| P25705 | P25705 | P25705 | P25705 | | P25705 | P25705 | P25705 |
| P08670 | P08670 | P08670 | P08670 | | P08670 | P08670 | P08670 |
| P61158 | P61158 | P61158 | P61158 | | P61158 | P61158 | P61158 |
| Q9H0U4 | Q9H0U4 | Q9H0U4 | Q9H0U4 | | Q9H0U4 | Q9H0U4 | Q9H0U4 |
| Q92928 | Q92928 | Q92928 | Q92928 | | Q92928 | Q92928 | Q92928 |
| P62820 | P62820 | P62820 | P62820 | | P62820 | P62820 | P62820 |
| Q92930 | Q92930 | Q92930 | Q92930 | | Q92930 | Q92930 | Q92930 |
| P61026 | P61026 | P61026 | P61026 | | P61026 | P61026 | P61026 |
| P59190 | P59190 | P59190 | P59190 | | P59190 | P59190 | P59190 |

| | | | | | | | |
|---|---|---|---|---|---|---|---|
| Q15286 | Q15286 | Q15286 | Q15286 | | Q15286 | Q15286 | Q15286 |
| P51153 | P51153 | P51153 | P51153 | | P51153 | P51153 | P51153 |
| P61006 | P61006 | P61006 | P61006 | | P61006 | P61006 | P61006 |
| P61160 | P61160 | P61160 | P61160 | | P61160 | P61160 | P61160 |
| P06576 | P06576 | P06576 | P06576 | | P06576 | P06576 | |
| P60660 | P60660 | P60660 | P60660 | | P60660 | P60660 | P60660 |
| P14649 | P14649 | P14649 | P14649 | | P14649 | P14649 | P14649 |
| P05386 | P05386 | P05386 | P05386 | | P05386 | P05386 | P05386 |
| Q14764 | Q14764 | Q14764 | Q14764 | | Q14764 | Q14764 | Q14764 |
| Q14118 | Q14118 | Q14118 | Q14118 | | Q14118 | Q14118 | Q14118 |
| P04217 | P04217 | P04217 | P04217 | | P04217 | P04217 | P04217 |
| P36952 | P36952 | P36952 | P36952 | | P36952 | P36952 | P36952 |
| Q02809 | Q02809 | Q02809 | Q02809 | | Q02809 | Q02809 | Q02809 |
| Q6ZVX7 | Q6ZVX7 | Q6ZVX7 | Q6ZVX7 | | Q6ZVX7 | Q6ZVX7 | Q6ZVX7 |
| P14174 | P14174 | P14174 | P14174 | | P14174 | P14174 | P14174 |
| Q58FF3 | Q58FF3 | Q58FF3 | Q58FF3 | | Q58FF3 | Q58FF3 | Q58FF3 |
| P14625 | P14625 | P14625 | P14625 | | P14625 | P14625 | P14625 |
| P25786 | P25786 | P25786 | P25786 | | P25786 | P25786 | P25786 |
| O95436 | O95436 | O95436 | O95436 | | O95436 | O95436 | O95436 |
| P13489 | P13489 | P13489 | P13489 | | P13489 | P13489 | P13489 |
| Q9UL46 | Q9UL46 | Q9UL46 | Q9UL46 | | Q9UL46 | Q9UL46 | Q9UL46 |
| P36955 | P36955 | P36955 | P36955 | | P36955 | P36955 | P36955 |
| P60981 | P60981 | P60981 | P60981 | | P60981 | P60981 | P60981 |
| P62913 | P62913 | P62913 | P62913 | | P62913 | P62913 | P62913 |
| O75888 | O75888 | O75888 | O75888 | | O75888 | O75888 | O75888 |
| P57723 | P57723 | P57723 | P57723 | | P57723 | P57723 | P57723 |
| Q15365 | Q15365 | Q15365 | Q15365 | | Q15365 | Q15365 | Q15365 |
| P57721 | P57721 | P57721 | P57721 | | P57721 | P57721 | P57721 |
| Q15366 | Q15366 | Q15366 | Q15366 | | Q15366 | Q15366 | Q15366 |
| P15309 | | P15309 | P15309 | | P15309 | P15309 | P15309 |
| P62491 | P62491 | P62491 | P62491 | | P62491 | P62491 | P62491 |
| Q15907 | Q15907 | Q15907 | Q15907 | | Q15907 | Q15907 | Q15907 |
| Q13867 | Q13867 | Q13867 | Q13867 | | Q13867 | Q13867 | Q13867 |

| | | | | | | | |
|---|---|---|---|---|---|---|---|
| P33241 | P33241 | P33241 | P33241 | | P33241 | P33241 | P33241 |
| P27635 | P27635 | P27635 | P27635 | | P27635 | P27635 | P27635 |
| O75083 | O75083 | O75083 | O75083 | | O75083 | O75083 | O75083 |
| P09467 | P09467 | P09467 | P09467 | | P09467 | P09467 | P09467 |
| P02452 | | | | | | | |
| P18124 | P18124 | P18124 | P18124 | | P18124 | P18124 | P18124 |
| P62899 | P62899 | P62899 | P62899 | | P62899 | P62899 | P62899 |
| P25789 | P25789 | P25789 | P25789 | | P25789 | P25789 | P25789 |
| Q9UN76 | Q9UN76 | Q9UN76 | Q9UN76 | | Q9UN76 | Q9UN76 | Q9UN76 |
| P06748 | P06748 | P06748 | P06748 | | P06748 | P06748 | P06748 |
| O43516 | O43516 | O43516 | O43516 | | O43516 | O43516 | O43516 |
| Q99497 | Q99497 | Q99497 | Q99497 | | Q99497 | Q99497 | Q99497 |
| Q86X10 | Q86X10 | Q86X10 | | | Q86X10 | | |
| P40939 | P40939 | P40939 | P40939 | | P40939 | P40939 | P40939 |
| P09668 | P09668 | P09668 | P09668 | | P09668 | P09668 | P09668 |
| O14497 | O14497 | O14497 | O14497 | | O14497 | O14497 | O14497 |
| Q13200 | Q13200 | Q13200 | Q13200 | | Q13200 | Q13200 | Q13200 |
| Q96SC8 | Q96SC8 | Q96SC8 | Q96SC8 | | Q96SC8 | Q96SC8 | Q96SC8 |
| Q14574 | Q14574 | Q14574 | Q14574 | | Q14574 | Q14574 | Q14574 |
| Q9UPN9 | Q9UPN9 | Q9UPN9 | Q9UPN9 | | Q9UPN9 | Q9UPN9 | Q9UPN9 |
| P08174 | P08174 | P08174 | P08174 | | P08174 | P08174 | P08174 |
| P06727 | | P06727 | P06727 | | P06727 | P06727 | P06727 |
| | | Q5T1R4 | | | | | |
| Q96JM2 | Q96JM2 | Q96JM2 | Q96JM2 | | Q96JM2 | Q96JM2 | Q96JM2 |
| Q96RY5 | Q96RY5 | Q96RY5 | Q96RY5 | | Q96RY5 | Q96RY5 | Q96RY5 |
| P04179 | P04179 | P04179 | P04179 | | P04179 | P04179 | P04179 |

All proteins are listed by their Uniprot Accession

**Supplementary Table 3**

*Protein Identifications by Subject*

**Subject 1**

| Accession | Balafilcon A | Comfilcon A | Delefilcon A | Etafilcon A | Lotrafilcon B | Nelfilcon A | Nesofilcon A | Senofilcon A | Verofilcon A |
|---|---|---|---|---|---|---|---|---|---|
| Totals | 0 | 275 | 280 | 277 | 279 | 277 | 279 | 273 | 278 |
| P02788\|TRFL_HUMAN | | 5.690724 | 4.871878 | 4.971168 | 5.147946 | 4.729811 | 4.194721 | 5.29558 | 7.346494 |
| P35527\|K1C9_HUMAN | | 2.640408 | 2.653632 | 2.500222 | 2.682099 | 2.618872 | 2.715624 | 2.696039 | 2.639785 |
| P04264\|K2C1_HUMAN | | 3.655624 | 3.261086 | 3.113812 | 3.413111 | 3.583112 | 3.166784 | 3.271805 | 3.393056 |
| P13645\|K1C10_HUMAN | | 2.472302 | 1.775869 | 1.558176 | 2.033123 | 2.574902 | 1.281023 | 1.943836 | 2.087936 |
| P35908\|K22E_HUMAN | | 1.896558 | 1.339147 | 1.060623 | 1.916443 | 2.286693 | 0.769479 | 1.607368 | 1.54259 |
| P61626\|LYSC_HUMAN | | 6.122913 | 6.120219 | 7.936636 | 5.045267 | 5.649285 | 6.657784 | 5.443738 | 7.725464 |
| P02768\|ALBU_HUMAN | | 2.994 | 2.866583 | 2.947026 | 3.733974 | 3.375815 | 2.488064 | 3.027271 | 4.047957 |
| P31025\|LCN1_HUMAN | | 4.804439 | 4.450895 | 4.599402 | 4.515453 | 4.32522 | 3.899687 | 4.338181 | 4.429633 |
| P01833\|PIGR_HUMAN | | 1.169388 | 0.463922 | 0.790406 | 0.768693 | 1.556154 | 0.252294 | 0.244853 | 1.263555 |
| P01876\|IGHA1_HUMAN | | 0.744885 | 0.058396 | 0.305906 | 0.463582 | 1.062527 | -0.12497 | -0.2989 | 0.816341 |
| P01024\|CO3_HUMAN | | -1.73484 | -2.19835 | -1.79919 | -1.41695 | -0.34289 | -1.90514 | -1.96992 | -1.27763 |
| P25311\|ZA2G_HUMAN | | 2.824343 | 2.387976 | 2.357498 | 2.482827 | 2.806957 | 1.388468 | 2.425736 | 2.806497 |
| P04259\|K2C6B_HUMAN | | -9.47332 | -10.8151 | -10.2246 | -10.5327 | -9.29973 | -8.34516 | | -9.73089 |
| P19013\|K2C4_HUMAN | | -0.05136 | -0.04315 | -1.32839 | -0.21754 | 1.402761 | -0.67122 | -1.10658 | 0.18399 |
| P13647\|K2C5_HUMAN | | 0.989055 | 1.01902 | 0.360239 | 0.811495 | 2.139689 | 0.525628 | 0.412755 | 1.111652 |
| P02538\|K2C6A_HUMAN | | -6.79927 | -7.08096 | -7.42274 | -8.26161 | -6.37327 | -7.08513 | -8.62578 | -7.41315 |
| P08727\|K1C19_HUMAN | | -2.09435 | -1.91331 | -2.82852 | -1.80989 | -0.58766 | -2.18012 | -2.7141 | -1.55462 |

| | | | | | | | | |
|---|---|---|---|---|---|---|---|---|
| P13646\|K1C13_HUMAN | -1.05261 | -0.99042 | -2.36572 | -1.19364 | 0.468996 | -1.5962 | -2.43853 | -0.84505 |
| P08779\|K1C16_HUMAN | -1.02142 | -1.39949 | -0.98303 | -1.12571 | -0.7594 | -0.52679 | -1.47697 | -0.95396 |
| P02533\|K1C14_HUMAN | -1.35707 | -1.77041 | -1.85073 | -1.75414 | -1.15355 | -1.77524 | -1.73294 | -1.43775 |
| Q9UGM3\|DMBT1_HUMAN | -1.85694 | -2.29903 | -2.12636 | -2.11166 | -1.86316 | -2.80307 | -2.58966 | -1.99766 |
| P02787\|TRFE_HUMAN | -0.41073 | -0.72151 | 0.065129 | -0.16936 | -0.10705 | -0.8272 | -0.43361 | 0.692201 |
| P0DOX7\|IGK_HUMAN | 0.202038 | -0.37487 | -0.07498 | -0.11181 | 0.703482 | -0.59886 | -0.55347 | 0.386494 |
| P98160\|PGBM_HUMAN | -3.17445 | -3.32512 | -2.94325 | -3.389 | -2.63392 | -3.98916 | -3.66582 | -3.13169 |
| P60709\|ACTB_HUMAN | -2.77477 | -2.98564 | -2.78246 | -2.4187 | -0.39934 | -2.91089 | -3.55801 | -1.91192 |
| P63261\|ACTG_HUMAN | -2.77477 | -2.98564 | -2.78246 | -2.4187 | -0.39934 | -2.91089 | -3.55801 | -1.91192 |
| P15924\|DESP_HUMAN | -0.23747 | -0.77691 | -0.74279 | -0.49167 | -0.53948 | -0.60928 | -0.78958 | -0.64787 |
| P06733\|ENOA_HUMAN | -1.07151 | -1.5988 | -1.61221 | -0.8039 | 1.735394 | -1.50819 | -1.12058 | -0.1456 |
| P01036\|CYTS_HUMAN | 0.974397 | 0.494677 | 0.635129 | 0.651335 | 0.94984 | 1.078228 | 0.281506 | 1.127429 |
| P12273\|PIP_HUMAN | 2.299667 | 2.044471 | 2.048426 | 2.170166 | 2.288968 | 1.025234 | 1.844631 | 2.492993 |
| Q13421\|MSLN_HUMAN | -6.20067 | -6.48875 | -5.96648 | -5.99726 | -5.57447 | -6.98327 | -6.54741 | -5.65453 |
| P01037\|CYTN_HUMAN | 0.28956 | -0.20532 | 0.335586 | -0.33806 | 0.554615 | 1.42066 | -0.48346 | 0.884203 |
| P0DOX2\|IGA2_HUMAN | -4.27794 | -4.85752 | -4.70455 | -4.82345 | -3.70246 | -5.26845 | -5.23179 | -3.99639 |
| P07355\|ANXA2_HUMAN | -0.53242 | -0.99659 | -1.16652 | -0.6605 | 0.540356 | -1.39075 | -1.2241 | 0.639059 |
| P00450\|CERU_HUMAN | -3.55573 | -3.61227 | -3.26985 | -3.64878 | -2.81314 | -3.79041 | -3.84061 | -2.85352 |
| P06396\|GELS_HUMAN | -1.98107 | -2.21835 | -2.00675 | -1.5941 | -0.35907 | -2.30329 | -2.52792 | -1.40071 |

| | | | | | | | | |
|---|---|---|---|---|---|---|---|---|
| P00738\|HPT_HUMAN | 0.426634 | -0.11363 | 0.20588 | -0.00042 | 0.307992 | -1.0426 | 0.066075 | 0.187686 |
| P0DOY2\|IGLC2_HUMAN | -3.89535 | -4.60487 | -4.43465 | -4.1614 | -3.2187 | -4.7109 | -4.65177 | -3.96852 |
| B9A064\|IGLL5_HUMAN | -2.67315 | -3.35344 | -3.17717 | -3.05307 | -2.16989 | -3.51421 | -3.57848 | -2.67707 |
| P0DOX8\|IGL1_HUMAN | -2.67315 | -3.35344 | -3.17717 | -3.05307 | -2.16989 | -3.51421 | -3.57848 | -2.67707 |
| P0DOX5\|IGG1_HUMAN | -2.18369 | -2.35972 | -0.94885 | -2.59593 | -1.72468 | -2.02135 | -2.32062 | -0.95108 |
| P10909\|CLUS_HUMAN | -1.37049 | -1.59956 | -1.46922 | -1.48503 | -0.43213 | -2.22025 | -1.80876 | -1.44033 |
| P08729\|K2C7_HUMAN | -5.93867 | -5.49625 | -6.42654 | -5.0507 | -4.03791 | -5.3867 | -6.26519 | -4.4809 |
| P04083\|ANXA1_HUMAN | -1.38156 | -1.76514 | -1.9269 | -1.13614 | -0.40493 | -1.88704 | -2.32835 | -0.72259 |
| P60174\|TPIS_HUMAN | -3.5366 | -3.80917 | -3.90167 | -3.44593 | -1.46896 | -4.05468 | -3.88457 | -2.76901 |
| P14923\|PLAK_HUMAN | -0.85102 | -1.30489 | -1.18115 | -1.02291 | -0.90492 | -1.23083 | -1.1946 | -1.04739 |
| P00352\|AL1A1_HUMAN | -6.43791 | -6.34092 | -6.21831 | -5.69049 | -3.2025 | -6.33856 | -6.96144 | -5.14282 |
| P01871\|IGHM_HUMAN | -5.54578 | -6.33794 | -6.2071 | -5.79926 | -4.97737 | -6.27174 | -5.51038 | -5.63898 |
| P14618\|KPYM_HUMAN | -3.47721 | -4.3375 | -3.53152 | -3.66643 | -1.89729 | -4.48094 | -3.89682 | -3.26555 |
| P05787\|K2C8_HUMAN | -3.90425 | -3.56319 | -4.58262 | -3.36624 | -2.38692 | -3.84219 | -4.71206 | -3.02123 |
| O75556\|SG2A1_HUMAN | 0.052594 | -0.64659 | 1.573758 | -0.39696 | -0.03065 | 0.534324 | -0.96893 | -0.29723 |
| P30838\|AL3A1_HUMAN | -2.88163 | -3.30998 | -2.20401 | -3.30186 | -0.20268 | -3.16961 | -3.29099 | -1.70003 |
| P0DMV8\|HS71A_HUMAN | -5.17299 | -5.41332 | -5.11104 | -4.1203 | -3.14598 | -5.32743 | -5.67683 | -3.81856 |
| P0DMV9\|HS71B_HUMAN | -5.17299 | -5.41332 | -5.11104 | -4.1203 | -3.14598 | -5.32743 | -5.67683 | -3.81856 |
| Q08380\|LG3BP_HUMAN | -1.41392 | -2.06269 | -1.5077 | -1.55367 | -1.1898 | -2.47457 | -1.50929 | -1.42224 |
| Q16378\|PROL4_HUMAN | 2.126929 | 1.737121 | 2.766235 | 1.765878 | 0.991276 | 1.789382 | 1.570162 | 2.059822 |

| | | | | | | | | |
|---|---|---|---|---|---|---|---|---|
| Q04695\|K1C17_HUMAN | -3.70758 | -3.98088 | -4.19681 | -4.1619 | -3.85442 | -3.37553 | -4.32075 | -3.7047 |
| P68133\|ACTS_HUMAN | -6.06053 | -5.89285 | -6.45398 | -5.28255 | -3.10777 | -5.88664 | -6.49814 | -4.98677 |
| P68032\|ACTC_HUMAN | -6.06053 | -5.89285 | -6.45398 | -5.28255 | -3.10777 | -5.88664 | -6.49814 | -4.98677 |
| P04792\|HSPB1_HUMAN | -2.52593 | -2.62182 | -2.66703 | -0.60574 | -0.43266 | -2.57405 | -2.82838 | -1.6459 |
| P01009\|A1AT_HUMAN | -3.90883 | -4.01937 | -4.33026 | -3.40466 | -3.32414 | -4.50174 | -3.98187 | -2.9635 |
| P12035\|K2C3_HUMAN | -4.56971 | -5.20682 | -4.13358 | -3.68782 | -7.44879 | -5.73756 | -5.58615 | -3.95074 |
| P06702\|S10A9_HUMAN | -1.33814 | -1.94195 | -2.64455 | -1.46112 | -0.02424 | -2.13426 | -2.4551 | -0.96274 |
| P22079\|PERL_HUMAN | -1.84745 | -2.26613 | -2.07111 | -2.20279 | -1.96242 | -3.08691 | -2.48045 | -1.14724 |
| P01859\|IGHG2_HUMAN | -4.52429 | -5.03471 | -4.87573 | -4.52997 | -3.58385 | -5.16534 | -4.50471 | -4.17109 |
| P20061\|TCO1_HUMAN | -3.07965 | -3.3543 | -3.14413 | -3.2593 | -2.82857 | -3.85707 | -3.52066 | -2.70914 |
| Q02413\|DSG1_HUMAN | -2.00707 | -2.60048 | -2.13757 | -2.31299 | -2.56814 | -2.77426 | -2.44333 | -1.9919 |
| P19012\|K1C15_HUMAN | -9.72751 | -9.43751 | | -9.19824 | -8.12715 | -9.50229 | | -8.71101 |
| P30740\|ILEU_HUMAN | -3.16718 | -3.54897 | -3.38423 | -2.95012 | -1.04522 | -3.65465 | -3.84648 | -2.13675 |
| P80188\|NGAL_HUMAN | -2.06072 | -2.36404 | -2.36524 | -1.53363 | -0.97769 | -2.10894 | -2.22978 | -1.36809 |
| Q99935\|PROL1_HUMAN | -0.59318 | -0.68125 | -0.45332 | 0.46195 | -0.11033 | -1.42781 | -1.26743 | -0.90342 |
| P04406\|G3P_HUMAN | -1.65444 | -2.47494 | -2.43731 | -2.01191 | -0.10726 | -2.7829 | -2.1044 | -1.2342 |
| P03973\|SLPI_HUMAN | 0.9309 | 1.128068 | 2.947734 | 0.098507 | -0.22618 | 1.264906 | 0.230864 | 0.879523 |
| P98088\|MUC5A_HUMAN | -4.77874 | -6.16876 | -7.15076 | -5.5338 | -2.91712 | -5.73338 | -7.84162 | -5.03304 |
| P14555\|PA2GA_HUMAN | 0.306817 | -0.50239 | 1.319054 | -0.13116 | -1.08815 | -1.39368 | -1.36248 | -1.78186 |

| | | | | | | | | |
|---|---|---|---|---|---|---|---|---|
| P01591\|IGJ_HUMAN | 1.621904 | 0.863885 | 1.15314 | 1.177101 | 2.075459 | 0.632553 | 0.553693 | 1.581436 |
| O95968\|SG1D1_HUMAN | 1.336028 | 1.081845 | 2.505184 | 1.331934 | 2.026642 | 1.698593 | 0.593965 | 1.324656 |
| Q7Z794\|K2C1B_HUMAN | -5.18343 | -6.198 | -5.8329 | -5.50669 | -5.30382 | -6.47353 | -5.87595 | -4.58714 |
| Q8N1N4\|K2C78_HUMAN | -2.52361 | -2.875 | -3.18508 | -2.7936 | -2.2209 | -3.32568 | -3.00494 | -2.44434 |
| P63104\|1433Z_HUMAN | -3.29073 | -3.57023 | -3.43347 | -2.69514 | -0.65795 | -3.55786 | -3.55193 | -2.08525 |
| P31944\|CASPE_HUMAN | -3.45934 | -4.61478 | -4.25325 | -3.97926 | -3.09797 | -4.76212 | -4.04857 | -3.70049 |
| Q96P63\|SPB12_HUMAN | -3.55018 | -3.87356 | -3.2068 | -4.12846 | -3.83199 | -3.85661 | -3.99138 | -3.25183 |
| P09228\|CYTT_HUMAN | -6.46619 | -6.95475 | -6.87266 | -6.94278 | -6.37197 | -5.85611 | -7.16257 | -6.12861 |
| P09211\|GSTP1_HUMAN | -3.7241 | -4.0293 | -3.94405 | -3.27672 | -0.84625 | -3.88678 | -3.94917 | -2.56202 |
| Q9GZZ8\|LACRT_HUMAN | 3.472011 | 2.63853 | 4.123645 | 2.73628 | 3.551181 | 2.410184 | 2.261791 | 2.702174 |
| P02647\|APOA1_HUMAN | -4.9386 | -4.72009 | -4.73094 | -3.00322 | -3.41234 | -5.23459 | -4.43072 | -4.36827 |
| Q96DA0\|ZG16B_HUMAN | -1.4158 | -1.29429 | -1.55954 | -1.16113 | -0.88204 | -2.69355 | -1.7538 | -0.97523 |
| P05109\|S10A8_HUMAN | -0.33673 | -0.87101 | -1.96241 | -0.68769 | 0.692575 | -1.10502 | -1.54314 | -0.37074 |
| P07602\|SAP_HUMAN | -4.10545 | -4.33008 | -3.85803 | -4.02388 | -3.20576 | -4.47375 | -4.23739 | -3.91163 |
| P05090\|APOD_HUMAN | -2.60645 | 0.472118 | -3.06546 | 0.208537 | -1.88969 | -1.87049 | -0.98809 | -1.7284 |
| P11142\|HSP7C_HUMAN | -6.84932 | -6.98051 | -6.13591 | -6.13285 | -5.32629 | -6.85378 | -7.2469 | -5.56337 |
| Q5D862\|FILA2_HUMAN | -3.45339 | -4.19164 | -4.29676 | -3.82616 | -3.713 | -4.67146 | -4.05533 | -3.60361 |
| P61769\|B2MG_HUMAN | -2.266 | -2.8712 | -2.11933 | -2.49525 | -2.7227 | -0.42384 | -2.8545 | -1.13285 |
| Q01469\|FABP5_HUMAN | -4.07993 | -4.89848 | -4.62833 | -4.43179 | -1.80265 | -4.03495 | -4.44972 | -3.71897 |
| P34096\|RNAS4_HUMAN | -1.79812 | -2.25694 | -0.06668 | -2.37712 | -2.32576 | -1.73408 | -2.64449 | -0.97136 |

| | | | | | | | | |
|---|---|---|---|---|---|---|---|---|
| P07339\|CATD_HUMAN | -2.82259 | -3.5013 | -3.59386 | -3.1189 | -2.03175 | -3.5444 | -3.75442 | -3.2793 |
| P30044\|PRDX5_HUMAN | -8.43586 | -8.33797 | -7.52547 | -7.2939 | -4.70065 | -7.29509 | -8.29004 | -6.61543 |
| P04075\|ALDOA_HUMAN | -5.01314 | -5.71448 | -5.25698 | -5.41228 | -3.28951 | -5.75177 | -6.04533 | -4.40567 |
| P31947\|1433S_HUMAN | -5.76043 | -6.20323 | -6.22168 | -5.21762 | -2.89751 | -6.53682 | -5.86259 | -4.94573 |
| Q5VTE0\|EF1A3_HUMAN | -3.93521 | -4.65827 | -4.38751 | -4.00814 | -2.15874 | -4.64359 | -5.13883 | -2.99986 |
| P68104\|EF1A1_HUMAN | -3.93521 | -4.65827 | -4.38751 | -4.00814 | -2.15874 | -4.64359 | -5.13883 | -2.99986 |
| P62937\|PPIA_HUMAN | -3.75927 | -4.10819 | -4.24366 | -3.64695 | -1.83623 | -3.84448 | -4.07722 | -3.109 |
| P62805\|H4_HUMAN | -2.26928 | -2.21202 | -2.75529 | -2.1139 | -0.77948 | -2.50499 | -3.57244 | -1.88503 |
| P07737\|PROF1_HUMAN | -5.93239 | -6.11779 | -6.59324 | -5.53 | -3.07451 | -4.58952 | -6.26976 | -4.55283 |
| A0M8Q6\|IGLC7_HUMAN | -5.42765 | -6.22212 | -5.83748 | -5.36167 | -4.72615 | -6.52915 | -6.32444 | -5.07396 |
| Q99456\|K1C12_HUMAN | -3.08698 | -3.88216 | -2.65622 | -4.123 | -3.72699 | -4.92127 | -3.73865 | -3.28486 |
| Q06830\|PRDX1_HUMAN | -3.22509 | -3.57376 | -3.52978 | -3.14392 | -1.82424 | -3.5307 | -3.75569 | -2.73589 |
| P30086\|PEBP1_HUMAN | -6.14624 | -6.22659 | -6.64371 | -6.10364 | -3.74296 | -6.12667 | -6.08065 | -5.1581 |
| Q3SY84\|K2C71_HUMAN | -6.23188 | -6.57254 | -6.46216 | -7.01353 | -6.02593 | -6.7254 | -6.37259 | -5.85209 |
| P02545\|LMNA_HUMAN | -5.7672 | -6.83154 | -6.74614 | -6.42017 | -3.65068 | -6.67215 | -6.44965 | -6.94772 |
| P00338\|LDHA_HUMAN | -4.89224 | -4.99802 | -3.44951 | -5.03417 | -3.29983 | -4.63912 | -5.36746 | -3.35053 |
| P01034\|CYTC_HUMAN | -2.61858 | -3.4995 | -2.08727 | -2.99307 | -2.48555 | -1.90227 | -2.96617 | -2.27967 |
| P68363\|TBA1B_HUMAN | -8.52451 | -9.80431 | -8.95348 | -9.47495 | -7.58726 | -9.43201 | | -8.57345 |
| P52209\|6PGD_HUMAN | -5.31828 | -5.42321 | -5.15869 | -5.1509 | -2.57053 | -5.56956 | -5.63648 | -4.13067 |

| | | | | | | | | |
|---|---|---|---|---|---|---|---|---|
| Q8N474\|SFRP1_HUMAN | -2.63913 | -2.89885 | -2.6992 | -2.4562 | -3.19564 | -4.30895 | -3.51572 | -1.85957 |
| P01011\|AACT_HUMAN | -4.20971 | -5.07306 | -4.79196 | -4.76584 | -4.98769 | -5.61692 | -5.17631 | -3.42858 |
| P17931\|LEG3_HUMAN | -3.26511 | -3.59554 | -3.53072 | -2.94364 | -1.00052 | -3.21334 | -4.01438 | -2.05523 |
| P01619\|KV320_HUMAN | -2.95575 | -3.77475 | -3.50397 | -3.37169 | -2.60349 | -4.0326 | -3.62907 | -3.01424 |
| P11021\|BIP_HUMAN | -7.59759 | -7.80965 | -7.35912 | -6.69029 | -5.79285 | -7.22836 | -7.63084 | -6.73653 |
| P00558\|PGK1_HUMAN | -6.18238 | -6.95512 | -7.40982 | -6.32053 | -3.71454 | -6.83127 | -7.30515 | -5.51342 |
| P07858\|CATB_HUMAN | -5.28787 | -5.65641 | -5.56706 | -5.62379 | -5.00652 | -6.43944 | -5.52113 | -5.05581 |
| P29401\|TKT_HUMAN | -4.51089 | -5.16193 | -4.37116 | -3.52702 | -3.03039 | -4.85298 | -3.32041 | -3.7238 |
| P80303\|NUCB2_HUMAN | -2.88998 | -3.01567 | -2.38836 | -2.99837 | -2.27174 | -3.82074 | -3.53209 | -2.89735 |
| P15311\|EZRI_HUMAN | -5.88459 | -6.24443 | -5.88572 | -5.53874 | -3.681 | -5.77949 | -6.83174 | -4.93884 |
| Q08554\|DSC1_HUMAN | -3.59756 | -4.21717 | -4.06422 | -3.70378 | -4.03273 | -4.21519 | -3.97503 | -3.48879 |
| Q93079\|H2B1H_HUMAN | -3.30787 | -3.3345 | -3.67062 | -3.24569 | -1.95825 | -3.60132 | -4.19319 | -2.67934 |
| Q5QNW6\|H2B2F_HUMAN | -3.30787 | -3.3345 | -3.67062 | -3.24569 | -1.95825 | -3.60132 | -4.19319 | -2.67934 |
| Q99877\|H2B1N_HUMAN | -3.30787 | -3.3345 | -3.67062 | -3.24569 | -1.95825 | -3.60132 | -4.19319 | -2.67934 |
| O60814\|H2B1K_HUMAN | -3.30787 | -3.3345 | -3.67062 | -3.24569 | -1.95825 | -3.60132 | -4.19319 | -2.67934 |
| P62807\|H2B1C_HUMAN | -3.30787 | -3.3345 | -3.67062 | -3.24569 | -1.95825 | -3.60132 | -4.19319 | -2.67934 |
| P58876\|H2B1D_HUMAN | -3.30787 | -3.3345 | -3.67062 | -3.24569 | -1.95825 | -3.60132 | -4.19319 | -2.67934 |
| Q99880\|H2B1L_HUMAN | -3.30787 | -3.3345 | -3.67062 | -3.24569 | -1.95825 | -3.60132 | -4.19319 | -2.67934 |
| P57053\|H2BFS_HUMAN | -3.30787 | -3.3345 | -3.67062 | -3.24569 | -1.95825 | -3.60132 | -4.19319 | -2.67934 |

| | | | | | | | | |
|---|---|---|---|---|---|---|---|---|
| Q99879\|H2B1M_HUMAN | -3.30787 | -3.3345 | -3.67062 | -3.24569 | -1.95825 | -3.60132 | -4.19319 | -2.67934 |
| P27797\|CALR_HUMAN | -6.40666 | -6.42052 | -6.16831 | -5.66978 | -5.09841 | -6.11477 | -6.28456 | -5.30541 |
| Q6UXB2\|CXL17_HUMAN | -2.06737 | -1.3318 | -0.05492 | -3.06382 | -2.45974 | -2.62511 | -1.86024 | -2.12608 |
| A0A0B4J1X5\|HV374_HUMAN | -5.35614 | -6.26571 | -6.13828 | -6.09139 | -4.94031 | -6.37771 | -6.18354 | -5.68743 |
| P06744\|G6PI_HUMAN | -7.12776 | -7.56928 | -7.20427 | -7.16287 | -5.40295 | -7.68219 | -6.92837 | -6.56802 |
| P05783\|K1C18_HUMAN | -4.95249 | -5.81751 | -4.55832 | -4.48979 | -5.92273 | -6.24389 | -5.30919 | -4.6039 |
| P01040\|CYTA_HUMAN | -3.7952 | -4.50224 | -3.94738 | -3.85134 | -3.43438 | -4.14814 | -4.09167 | -4.10725 |
| P40394\|ADH7_HUMAN | -7.99795 | -8.52093 | -8.47589 | -7.46493 | -5.35525 | -8.43529 | -8.50708 | -6.90951 |
| A0A0B4J1V0\|HV315_HUMAN | -4.8302 | -5.2689 | -5.37528 | -5.18315 | -4.306 | -5.70011 | -5.9516 | -4.57127 |
| P80748\|LV321_HUMAN | -5.81698 | -6.37941 | -6.26232 | -5.90678 | -5.24504 | -6.75034 | -6.55175 | -5.67631 |
| P62979\|RS27A_HUMAN | -4.17845 | -4.84773 | -4.42964 | -4.64528 | -3.78408 | -4.86315 | -5.1185 | -4.17022 |
| P62987\|RL40_HUMAN | -4.17845 | -4.84773 | -4.42964 | -4.64528 | -3.78408 | -4.86315 | -5.1185 | -4.17022 |
| P0CG47\|UBB_HUMAN | -4.17845 | -4.84773 | -4.42964 | -4.64528 | -3.78408 | -4.86315 | -5.1185 | -4.17022 |
| P0CG48\|UBC_HUMAN | -4.17845 | -4.84773 | -4.42964 | -4.64528 | -3.78408 | -4.86315 | -5.1185 | -4.17022 |
| P16403\|H12_HUMAN | -7.12728 | -7.182 | -7.81785 | -7.43823 | -5.46556 | -7.41372 | -7.41288 | -7.45873 |
| P27348\|1433T_HUMAN | -7.5434 | -7.63403 | -8.02748 | -7.79028 | -6.38579 | -8.17398 | -8.21127 | -7.30352 |
| P07237\|PDIA1_HUMAN | -3.40544 | -3.91176 | -3.68454 | -2.98399 | -2.8057 | -3.97561 | -3.6883 | -2.83648 |
| P00751\|CFAB_HUMAN | -4.39531 | -4.6608 | -4.36991 | -3.6078 | -2.85891 | -4.40044 | -4.50635 | -3.6107 |
| P06312\|KV401_HUMAN | -5.27464 | -5.767 | -5.35878 | -5.80417 | -4.89817 | -6.09766 | -5.91321 | -4.5662 |
| P02765\|FETUA_HUMAN | -10.0997 | -9.50727 | -10.2918 | -9.06 | -7.57328 | -10.1091 | -9.81007 | -9.33276 |

| | | | | | | | | |
|---|---|---|---|---|---|---|---|---|
| P10412\|H14_HUMAN | -6.36481 | -6.50306 | -6.85291 | -6.04108 | -4.54528 | -6.50328 | -6.74719 | -6.23389 |
| P31941\|ABC3A_HUMAN | -6.84932 | -7.38035 | -7.20347 | -5.63356 | -4.66749 | -6.10245 | -7.52826 | -5.72423 |
| P55058\|PLTP_HUMAN | -4.96764 | -4.64785 | -3.98743 | -4.7799 | -4.51512 | -5.07309 | -5.10835 | -4.00868 |
| P01615\|KVD28_HUMAN | -7.24782 | -8.08874 | -7.68618 | -7.50392 | -6.83045 | -8.28748 | -8.40148 | -7.0075 |
| A0A075B6P5\|KV228_HUMAN | -7.24782 | -8.08874 | -7.68618 | -7.50392 | -6.83045 | -8.28748 | -8.40148 | -7.0075 |
| P81605\|DCD_HUMAN | -1.48236 | -0.87684 | -1.54624 | -2.271 | -1.42739 | -2.1374 | -1.73847 | -2.1881 |
| P02774\|VTDB_HUMAN | | -10.4691 | | -10.5688 | -10.2148 | -11.1945 | -10.2602 | -9.77017 |
| P14550\|AK1A1_HUMAN | -7.16815 | -7.83016 | -8.20162 | -7.06732 | -4.58319 | -7.70488 | -8.15324 | -6.47025 |
| A0A0B4J1Y9\|HV372_HUMAN | -8.69843 | -9.64439 | -9.43168 | -9.51602 | -8.84577 | -9.77457 | -9.71884 | -8.61135 |
| P32119\|PRDX2_HUMAN | -5.03226 | -5.41225 | -5.38166 | -5.20697 | -4.29407 | -5.52519 | -5.25787 | -5.02459 |
| P26447\|S10A4_HUMAN | -2.86549 | -3.02412 | -3.36986 | -2.21228 | -0.87866 | -2.85145 | -3.33252 | -1.82475 |
| P31151\|S10A7_HUMAN | -4.5115 | -5.77325 | -6.10479 | -4.54766 | -5.72983 | -5.9085 | -5.09182 | -6.02424 |
| Q15517\|CDSN_HUMAN | -5.68125 | -6.37697 | -6.44393 | -6.17292 | -5.89661 | -6.42879 | -6.12887 | -5.80945 |
| P20930\|FILA_HUMAN | -5.25284 | -5.93456 | -6.17291 | -5.82975 | -5.70182 | -6.33405 | -5.86554 | -5.72725 |
| P0DP08\|HVD82_HUMAN | -2.54937 | -3.0618 | -2.83871 | -2.85438 | -1.85524 | -3.1457 | -3.55941 | -2.34482 |
| P01825\|HV459_HUMAN | -2.54937 | -3.0618 | -2.83871 | -2.85438 | -1.85524 | -3.1457 | -3.55941 | -2.34482 |
| P0DP07\|HV431_HUMAN | -2.54937 | -3.0618 | -2.83871 | -2.85438 | -1.85524 | -3.1457 | -3.55941 | -2.34482 |
| A0A0C4DH41\|HV461_HUMAN | -2.54937 | -3.0618 | -2.83871 | -2.85438 | -1.85524 | -3.1457 | -3.55941 | -2.34482 |
| P0DP06\|HVD34_HUMAN | -2.54937 | -3.0618 | -2.83871 | -2.85438 | -1.85524 | -3.1457 | -3.55941 | -2.34482 |

| | | | | | | | | |
|---|---|---|---|---|---|---|---|---|
| P06331\|HV434_HUMAN | -2.54937 | -3.0618 | -2.83871 | -2.85438 | -1.85524 | -3.1457 | -3.55941 | -2.34482 |
| P01824\|HV439_HUMAN | -2.54937 | -3.0618 | -2.83871 | -2.85438 | -1.85524 | -3.1457 | -3.55941 | -2.34482 |
| A0A0A0MRZ8\|KVD11_HUMAN | -5.71144 | -6.23968 | -6.05472 | -6.24162 | -5.1309 | -6.76314 | -6.35898 | -5.62449 |
| P04433\|KV311_HUMAN | -5.71144 | -6.23968 | -6.05472 | -6.24162 | -5.1309 | -6.76314 | -6.35898 | -5.62449 |
| P31949\|S10AB_HUMAN | -5.31176 | -5.45169 | -6.12217 | -5.10304 | -3.72335 | -5.39545 | -6.71989 | -4.39476 |
| P60953\|CDC42_HUMAN | -6.58616 | -6.39147 | -6.82827 | -5.82575 | -4.56609 | -6.71603 | -7.29434 | -5.81848 |
| Q6KB66\|K2C80_HUMAN | -8.6678 | -8.9854 | -8.66817 | -8.50298 | -8.83253 | -9.22202 | -9.3679 | -8.50507 |
| P10599\|THIO_HUMAN | -3.66851 | -4.23336 | -4.40905 | -3.83814 | -1.46361 | -3.76825 | -4.42179 | -3.54292 |
| P40925\|MDHC_HUMAN | -6.71157 | -6.73842 | -7.04832 | -6.26346 | -4.46956 | -6.62107 | -6.75754 | -5.72258 |
| Q14515\|SPRL1_HUMAN | -3.2758 | -3.80876 | -3.77507 | -4.21336 | -3.97923 | -4.41994 | -3.68344 | -2.2857 |
| A0A075B6S5\|KV127_HUMAN | -5.0498 | -5.59981 | -5.41182 | -5.39444 | -5.15883 | -4.8985 | -5.73801 | -4.88592 |
| P01742\|HV169_HUMAN | -3.38891 | -4.00144 | -3.74552 | -3.71183 | -2.74427 | -4.19856 | -4.44511 | -3.31083 |
| A0A0C4DH31\|HV118_HUMAN | -5.93225 | -6.75694 | -6.29461 | -6.2108 | -5.25537 | -6.82069 | -6.81902 | -5.76985 |
| P23083\|HV102_HUMAN | -5.93225 | -6.75694 | -6.29461 | -6.2108 | -5.25537 | -6.82069 | -6.81902 | -5.76985 |
| P07900\|HS90A_HUMAN | -7.17481 | -7.12507 | -7.49967 | -7.19445 | -6.38177 | -7.54872 | -7.65819 | -5.83089 |
| P84243\|H33_HUMAN | -5.38144 | -5.63307 | -5.76599 | -5.62379 | -4.4813 | -5.90019 | -6.70789 | -5.21518 |
| P01700\|LV147_HUMAN | -5.09284 | -5.73612 | -5.34049 | -5.25019 | -4.71078 | -5.84335 | -5.64433 | -4.69205 |
| P23528\|COF1_HUMAN | -6.41264 | -6.69024 | -6.32327 | -5.9089 | -3.29867 | -6.7345 | -6.93139 | -5.88777 |
| P21980\|TGM2_HUMAN | -6.69092 | -6.11991 | -5.96997 | -5.02705 | -2.85984 | -5.84432 | -7.08259 | -4.8303 |
| Q09666\|AHNK_HUMAN | -5.12781 | -5.18978 | -5.4342 | -5.35433 | -3.46434 | -5.67116 | -5.59102 | -4.80682 |

| | | | | | | | | |
|---|---|---|---|---|---|---|---|---|
| P01624\|KV315_HUMAN | -5.22639 | -5.80216 | -5.59554 | -5.58831 | -4.55938 | -6.13954 | -6.1637 | -5.21378 |
| P07384\|CAN1_HUMAN | -6.74383 | -6.93144 | -6.41886 | -6.28678 | -5.1499 | -7.02036 | -7.37794 | -5.89148 |
| Q6UWP8\|SBSN_HUMAN | -8.35535 | -9.13308 | -9.51612 | -9.11153 | -8.09129 | -8.97319 | -9.55717 | -8.51171 |
| Q8NBJ4\|GOLM1_HUMAN | -3.93553 | -4.33025 | -4.12253 | -4.21388 | -3.55641 | -5.0821 | -4.45114 | -3.98212 |
| P40926\|MDHM_HUMAN | -6.10485 | -6.77644 | -6.27297 | -5.63351 | -5.16947 | -6.41185 | -6.13743 | -5.58447 |
| P68371\|TBB4B_HUMAN | -5.8466 | -6.38338 | -6.31187 | -6.05051 | -5.05668 | -7.42498 | -6.55967 | -5.43531 |
| A0A0A0MS15\|HV349_HUMAN | -6.00548 | -6.65234 | -6.50363 | -6.13148 | -6.02567 | -7.01524 | -6.04369 | -5.93646 |
| P01593\|KVD33_HUMAN | -7.4147 | -7.83731 | -6.71859 | -7.80045 | -7.22002 | -7.46696 | -7.8617 | -6.50321 |
| P01594\|KV133_HUMAN | -7.4147 | -7.83731 | -6.71859 | -7.80045 | -7.22002 | -7.46696 | -7.8617 | -6.50321 |
| P02766\|TTHY_HUMAN | -5.48347 | -5.57957 | -5.70222 | -4.7882 | -4.84292 | -5.98276 | -5.3704 | -4.53205 |
| Q08188\|TGM3_HUMAN | -7.27875 | -8.22653 | -8.67002 | -8.05235 | -7.98997 | -8.63642 | -8.37007 | -8.00155 |
| P05089\|ARGI1_HUMAN | -4.36059 | -4.9659 | -5.07895 | -4.92842 | -4.41203 | -5.22403 | -4.58677 | -4.47135 |
| Q9UBT3\|DKK4_HUMAN | | -10.8211 | -9.7002 | -9.11164 | | -10.8336 | | -8.53731 |
| O00584\|RNT2_HUMAN | -7.16858 | -7.65369 | -7.53956 | -7.91179 | -7.03712 | -8.67246 | -7.69309 | -7.35016 |
| A0A0B4J2D9\|KVD13_HUMAN | -6.76274 | -7.20959 | -5.68054 | -7.29353 | -6.60405 | -7.37245 | -7.35499 | -6.63479 |
| P0DP09\|KV113_HUMAN | -6.76274 | -7.20959 | -5.68054 | -7.29353 | -6.60405 | -7.37245 | -7.35499 | -6.63479 |
| O75874\|IDHC_HUMAN | -8.57235 | -9.0018 | -8.01177 | -8.4579 | -5.66305 | -8.1838 | -8.81793 | -7.24922 |
| P15814\|IGLL1_HUMAN | -1.22278 | -2.00807 | -1.8108 | -1.57018 | -0.73475 | -2.17013 | -2.43905 | -1.26027 |
| P30085\|KCY_HUMAN | -7.82263 | -7.86357 | -9.04732 | -7.78972 | -5.14652 | -8.34283 | -8.27369 | -7.16237 |
| A0A0C4DH34\|HV428_HUMAN | -5.55215 | -6.32039 | -5.97925 | -5.98519 | -5.21187 | -6.4086 | -6.75241 | -5.50841 |

| | | | | | | | | |
|---|---|---|---|---|---|---|---|---|
| P30101\|PDIA3_HUMAN | -7.11005 | -7.39414 | -6.19394 | -7.36082 | -6.77934 | -6.92359 | -7.61638 | -5.94746 |
| Q14697\|GANAB_HUMAN | | -11.0586 | -10.0431 | -9.74076 | -10.7397 | | | |
| P12830\|CADH1_HUMAN | -5.56963 | -5.68427 | -4.32576 | -5.819 | -5.9842 | -6.28645 | -5.92136 | -5.09635 |
| Q16651\|PRSS8_HUMAN | -5.17261 | -5.3124 | -5.27172 | -4.53104 | -4.47944 | -5.78037 | -5.26114 | -4.30494 |
| P13987\|CD59_HUMAN | -6.95568 | -6.96621 | -7.24699 | -6.99895 | -5.89386 | -7.76882 | -7.33792 | -6.59257 |
| O43653\|PSCA_HUMAN | -5.23123 | -5.16124 | -5.13579 | -4.96359 | -2.57738 | -5.21533 | -6.18174 | -4.75298 |
| P02763\|A1AG1_HUMAN | -8.66832 | -9.23387 | -8.91938 | -8.363 | -8.46434 | -9.19836 | -8.44463 | -7.52723 |
| A0A0B4J1V6\|HV373_HUMAN | -6.75625 | -7.32907 | -6.99541 | -7.36107 | -5.91565 | -7.83755 | -7.35578 | -6.7685 |
| O43852\|CALU_HUMAN | -5.87137 | -6.17875 | -5.85043 | -5.70424 | -5.37985 | -6.8695 | -6.57881 | -5.53956 |
| P08758\|ANXA5_HUMAN | -6.16225 | -6.61341 | -6.82458 | -6.09292 | -5.78611 | -6.43174 | -6.4031 | -5.99975 |
| Q5T749\|KPRP_HUMAN | -5.17286 | -5.9887 | -6.00506 | -6.20077 | -5.87857 | -6.47662 | -6.27134 | -5.78196 |
| Q7Z5P9\|MUC19_HUMAN | -5.27027 | -6.20724 | -4.91221 | -5.85476 | -5.28385 | -6.74969 | -5.43951 | -3.87094 |
| P01714\|LV319_HUMAN | -9.02416 | -9.44793 | -9.46087 | -9.16026 | -8.26173 | -10.0031 | -10.194 | -9.02911 |
| P01717\|LV325_HUMAN | -5.6369 | -6.34466 | -6.23239 | -5.8831 | -5.22351 | -6.6104 | -6.38164 | -5.64145 |
| A0A075B6K4\|LV310_HUMAN | -5.6369 | -6.34466 | -6.23239 | -5.8831 | -5.22351 | -6.6104 | -6.38164 | -5.64145 |
| P23284\|PPIB_HUMAN | -3.28802 | -4.20493 | -2.42915 | -4.78103 | -4.64324 | -4.09193 | -3.85678 | -1.79811 |
| P12814\|ACTN1_HUMAN | -6.34292 | -6.58848 | -6.74864 | -5.75661 | -5.42567 | -6.29365 | -7.71446 | -5.33556 |
| P40199\|CEAM6_HUMAN | -6.0355 | -6.30324 | -5.01341 | -5.67632 | -4.68274 | -6.45369 | -6.05053 | -5.90221 |
| Q02818\|NUCB1_HUMAN | -5.16519 | -5.52408 | -5.21282 | -5.47965 | -4.7566 | -6.32543 | -5.91081 | -5.39556 |
| P49788\|TIG1_HUMAN | -8.64523 | -8.60231 | -8.65626 | -9.0191 | -7.6098 | -8.94308 | -8.76308 | -7.63557 |

| | | | | | | | | |
|---|---|---|---|---|---|---|---|---|
| P06703\|S10A6_HUMAN | -3.70841 | -3.99943 | -3.31009 | -2.85984 | -2.23263 | -3.60962 | -3.92036 | -3.05362 |
| P16401\|H15_HUMAN | -6.31906 | -6.36685 | -7.04766 | -6.43177 | -4.62089 | -6.49455 | -6.78688 | -6.39094 |
| O60437\|PEPL_HUMAN | -6.41009 | -6.73338 | -6.60541 | -6.8131 | -5.34855 | -7.69353 | -7.34372 | -6.19388 |
| P47929\|LEG7_HUMAN | -4.8597 | -6.26449 | -5.72841 | -5.21901 | -4.98152 | -6.07949 | -6.15853 | -5.58743 |
| Q14508\|WFDC2_HUMAN | -7.24486 | -6.51606 | -6.41956 | -6.07153 | -6.31294 | -6.26811 | -6.59425 | -5.90763 |
| Q13835\|PKP1_HUMAN | -6.42676 | -6.49026 | -6.55865 | -6.43118 | -5.88512 | -7.15824 | -6.12729 | -6.52063 |
| P02750\|A2GL_HUMAN | -8.8396 | -9.1117 | -9.454 | -8.60875 | -7.31447 | -8.83956 | -9.05208 | -7.4578 |
| P21926\|CD9_HUMAN | -4.88626 | -4.97864 | -4.24529 | -5.26891 | -3.34706 | -4.69998 | -5.30419 | -4.13642 |
| P19961\|AMY2B_HUMAN | -5.38664 | -6.57653 | -6.12371 | -6.35846 | -6.17991 | -7.10629 | -6.24363 | -5.66286 |
| P0DTE7\|AMY1B_HUMAN | -5.38664 | -6.57653 | -6.12371 | -6.35846 | -6.17991 | -7.10629 | -6.24363 | -5.66286 |
| P04746\|AMYP_HUMAN | -5.38664 | -6.57653 | -6.12371 | -6.35846 | -6.17991 | -7.10629 | -6.24363 | -5.66286 |
| P04745\|AMY1A_HUMAN | -5.38664 | -6.57653 | -6.12371 | -6.35846 | -6.17991 | -7.10629 | -6.24363 | -5.66286 |
| P0DTE8\|AMY1C_HUMAN | -5.38664 | -6.57653 | -6.12371 | -6.35846 | -6.17991 | -7.10629 | -6.24363 | -5.66286 |
| Q9UBC9\|SPRR3_HUMAN | -4.64326 | -5.53079 | -7.45205 | -5.77754 | -3.58893 | -5.74193 | -6.79356 | -5.47883 |
| Q96QA5\|GSDMA_HUMAN | -4.69146 | -5.53602 | -5.07734 | -5.23402 | -4.69534 | -5.88529 | -5.38087 | -4.81229 |
| Q13296\|SG2A2_HUMAN | -3.92156 | -4.72639 | -3.89247 | -4.56964 | -4.6491 | -4.39205 | -5.19716 | -4.26395 |
| Q14002\|CEAM7_HUMAN | -8.4064 | -8.36751 | -8.22688 | -8.47568 | -6.97174 | -8.57095 | -9.48277 | -7.66684 |
| P62753\|RS6_HUMAN | -5.37324 | -6.0273 | -5.49103 | -5.41294 | -5.15756 | -6.2748 | -5.58988 | -5.72303 |
| P09429\|HMGB1_HUMAN | -3.32488 | -5.53764 | -3.6334 | -4.22501 | -4.09273 | -4.96954 | -3.61606 | -4.3485 |

| | | | | | | | | |
|---|---|---|---|---|---|---|---|---|
| P83731\|RL24_HUMAN | -3.50667 | -5.52218 | -3.50134 | -3.85346 | -4.39129 | -4.90005 | -3.7431 | -4.93323 |
| Q96S96\|PEBP4_HUMAN | -7.25227 | -7.50747 | -7.46673 | -8.26347 | -7.59006 | -8.54728 | -7.49101 | -6.70571 |
| Q5SNV9\|CA167_HUMAN | -8.74368 | -9.62675 | -8.38498 | -8.83633 | -9.48695 | -9.81425 | -9.21232 | -7.04987 |
| Q6ZR08\|DYH12_HUMAN | -4.90204 | -5.72288 | -5.85493 | -5.02249 | -4.65515 | -6.16844 | -5.61661 | -5.55265 |
| P07910\|HNRPC_HUMAN | -8.68341 | -8.46397 | -8.93329 | -6.10687 | -7.47185 | -8.7381 | -9.32875 | -7.12046 |
| Q9UKZ1\|CNO11_HUMAN | -6.98712 | -7.46015 | -6.34341 | -7.35543 | -7.23032 | -7.0368 | -7.5629 | -5.83488 |
| P20933\|ASPG_HUMAN | -7.08566 | -8.77682 | -7.76528 | -7.64066 | -7.75364 | -8.59597 | -7.31794 | -8.16054 |
| Q8IZL8\|PELP1_HUMAN | -8.54875 | -9.85582 | -9.31881 | -8.7408 | -8.77594 | -9.82372 | -8.3143 | -8.89414 |
| P00505\|AATM_HUMAN | -6.07554 | -5.30337 | -5.84087 | -4.65004 | -4.64383 | -5.54525 | -5.58931 | -5.3518 |
| P48634\|PRC2A_HUMAN | | -10.9782 | | -11.0087 | | -11.8276 | | |
| Q96G74\|OTUD5_HUMAN | -8.66004 | -9.12975 | -8.54188 | -9.48592 | -8.40249 | -9.55808 | -8.85419 | -7.81671 |
| Q8IVF2\|AHNK2_HUMAN | -5.4026 | -6.15203 | -5.49963 | -6.04856 | -6.06265 | -3.41921 | -5.75259 | -4.65657 |
| Q8TF72\|SHRM3_HUMAN | -10.6705 | -8.9997 | -8.40182 | -8.53049 | -10.3623 | -9.87405 | -8.71353 | -8.6235 |
| Q14966\|ZN638_HUMAN | -4.94594 | -5.67779 | -4.42239 | -5.45581 | -4.96838 | -5.85507 | -5.35082 | -5.51751 |
| Q7LBE3\|S26A9_HUMAN | -3.5856 | -4.47892 | -4.45898 | -4.39823 | -4.36689 | -5.1575 | -4.0614 | -1.98143 |
| A6NKD9\|CC85C_HUMAN | -6.59761 | -6.80393 | -5.45449 | -8.02423 | -7.14666 | -6.41757 | -7.64823 | -5.24513 |
| A0A0B4J1U7\|HV601_HUMAN | -3.5085 | -4.13771 | -3.9008 | -3.76547 | -2.8582 | -4.3949 | -4.25919 | -2.87077 |
| Q6PID8\|KLD10_HUMAN | | -11.4141 | -9.11924 | | | -10.616 | | -9.19522 |
| Q99574\|NEUS_HUMAN | -8.25175 | -8.76881 | -8.66751 | -8.97838 | -8.46542 | -9.16544 | -8.95868 | -7.17077 |
| Q14526\|HIC1_HUMAN | -6.86121 | -7.53072 | -7.70944 | -7.55867 | -7.59338 | -8.32828 | -7.07015 | -5.08482 |

| | | | | | | | | |
|---|---|---|---|---|---|---|---|---|
| Q5VSY0\|GKAP1_HUMAN | -7.30782 | -7.90108 | -8.14322 | -7.16989 | -5.76703 | -8.04889 | -8.12672 | -6.62636 |
| Q8TCU6\|PREX1_HUMAN | -6.41865 | -5.79701 | -7.36593 | -6.36682 | -5.18473 | -6.99055 | -7.12253 | -5.89757 |
| Q7Z2W7\|TRPM8_HUMAN | -7.0547 | -7.7975 | -7.5493 | -8.19477 | -8.15266 | -7.76552 | -8.41114 | -5.94919 |
| Q14106\|TOB2_HUMAN | -6.88595 | -7.54463 | -7.43574 | -6.79204 | -6.90826 | -7.95579 | -7.57233 | -6.65584 |
| Q9GZQ3\|COMD5_HUMAN | -8.32065 | -9.54956 | -8.22953 | -9.06282 | -8.75948 | -9.39306 | -8.9227 | -8.40295 |
| Q8NHQ8\|RASF8_HUMAN | -2.44132 | -7.56089 | -6.69636 | -7.51004 | -7.17849 | -7.90284 | -7.20873 | -6.37483 |
| P01703\|LV140_HUMAN | -7.09701 | -7.7642 | -7.2964 | -7.37469 | -6.51802 | -7.98004 | -7.4098 | -6.50998 |
| O75882\|ATRN_HUMAN | -8.61462 | -8.92089 | -8.22028 | -9.63949 | -9.14574 | -8.83735 | -9.36946 | -7.89296 |
| O60902\|SHOX2_HUMAN | -6.82715 | -7.64228 | -7.26171 | -8.17867 | -7.61399 | -7.98286 | -7.96445 | -8.04109 |
| Q5VSP4\|LC1L1_HUMAN | | | | | | | | |
| P01861\|IGHG4_HUMAN | | | | | | | | |
| O95678\|K2C75_HUMAN | | | | | | | | |
| P0DOX6\|IGM_HUMAN | | | | | | | | |
| Q86YZ3\|HORN_HUMAN | | | | | | | | |
| Q14525\|KT33B_HUMAN | | | | | | | | |
| Q562R1\|ACTBL_HUMAN | | | | | | | | |
| P31946\|1433B_HUMAN | | | | | | | | |
| Q7Z3Y8\|K1C27_HUMAN | | | | | | | | |
| P01780\|HV307_HUMAN | | | | | | | | |
| A0A0J9YX35\|HV64D_HUMAN | | | | | | | | |
| A0A0J9YXX1\|HV5X1_HUMAN | | | | | | | | |
| A0A0C4DH38\|HV551_HUMAN | | | | | | | | |
| P28799\|GRN_HUMAN | | | | | | | | |
| P01762\|HV311_HUMAN | | | | | | | | |
| Q7Z3Y9\|K1C26_HUMAN | | | | | | | | |
| Q15149\|PLEC_HUMAN | | | | | | | | |

Q7Z406|MYH14_HUMAN
P06731|CEAM5_HUMAN
P04632|CPNS1_HUMAN
Q07666|KHDR1_HUMAN
O60716|CTND1_HUMAN
P01611|KVD12_HUMAN
A0A0C4DH72|KV106_HUMAN
P08246|ELNE_HUMAN
A0A075B6R9|KVD24_HUMAN
A0A0C4DH68|KV224_HUMAN
P40121|CAPG_HUMAN
O76021|RL1D1_HUMAN
A0A075B6H7|KV37_HUMAN
P42357|HUTH_HUMAN
P13797|PLST_HUMAN
P04211|LV743_HUMAN
A0A075B6I9|LV746_HUMAN
P55064|AQP5_HUMAN
P07951|TPM2_HUMAN
P09493|TPM1_HUMAN
P22735|TGM1_HUMAN
P01701|LV151_HUMAN
P49840|GSK3A_HUMAN
P0DOX3|IGD_HUMAN
P10809|CH60_HUMAN
P02679|FIBG_HUMAN
Q9UPP2|IQEC3_HUMAN
A0A075B6I0|LV861_HUMAN
Q86V81|THOC4_HUMAN
P22681|CBL_HUMAN
Q9BVC4|LST8_HUMAN
Q9BWS9|CHID1_HUMAN
Q7RTR2|NLRC3_HUMAN

Q8N6G6|ATL1_HUMAN
P67809|YBOX1_HUMAN
Q8IZ21|PHAR4_HUMAN
Q8WXS5|CCG8_HUMAN
Q9ULD2|MTUS1_HUMAN
Q86SM8|MRGRE_HUMAN
Q13283|G3BP1_HUMAN
Q9NSA2|KCND1_HUMAN
Q15323|K1H1_HUMAN
O43790|KRT86_HUMAN
Q14533|KRT81_HUMAN
P62258|1433E_HUMAN
Q13228|SBP1_HUMAN
P78386|KRT85_HUMAN
P13929|ENOB_HUMAN
A0A0C4DH42|HV366_HUMAN
P02790|HEMO_HUMAN
P22392|NDKB_HUMAN
P37802|TAGL2_HUMAN
P29508|SPB3_HUMAN
P18510|IL1RA_HUMAN
P30041|PRDX6_HUMAN
P15259|PGAM2_HUMAN
P18669|PGAM1_HUMAN
P48594|SPB4_HUMAN
P0DP24|CALM2_HUMAN
P0DP23|CALM1_HUMAN
P0DP25|CALM3_HUMAN
P05387|RLA2_HUMAN
P04080|CYTB_HUMAN
P22626|ROA2_HUMAN
O75223|GGCT_HUMAN
P37837|TALDO_HUMAN

P27482|CALL3_HUMAN
O00299|CLIC1_HUMAN
P08582|TRFM_HUMAN
Q9HC84|MUC5B_HUMAN
P0DME0|SETLP_HUMAN
Q01105|SET_HUMAN
P02511|CRYAB_HUMAN
P01008|ANT3_HUMAN
P08571|CD14_HUMAN
A0A075B6S9|KV137_HUMAN
P0DSN7|KVD37_HUMAN
P06454|PTMA_HUMAN
P08294|SODE_HUMAN
Q9HAV0|GBB4_HUMAN
P62873|GBB1_HUMAN
P62879|GBB2_HUMAN
P16520|GBB3_HUMAN
P25705|ATPA_HUMAN
P08670|VIME_HUMAN
P61158|ARP3_HUMAN
Q9H0U4|RAB1B_HUMAN
Q92928|RAB1C_HUMAN
P62820|RAB1A_HUMAN
Q92930|RAB8B_HUMAN
P61026|RAB10_HUMAN
P59190|RAB15_HUMAN
Q15286|RAB35_HUMAN
P51153|RAB13_HUMAN
P61006|RAB8A_HUMAN
P61160|ARP2_HUMAN
P06576|ATPB_HUMAN
P60660|MYL6_HUMAN
P14649|MYL6B_HUMAN

P05386|RLA1_HUMAN
Q14764|MVP_HUMAN
Q14118|DAG1_HUMAN
P04217|A1BG_HUMAN
P36952|SPB5_HUMAN
Q02809|PLOD1_HUMAN
Q6ZVX7|FBX50_HUMAN
P14174|MIF_HUMAN
Q58FF3|ENPLL_HUMAN
P14625|ENPL_HUMAN
P25786|PSA1_HUMAN
O95436|NPT2B_HUMAN
P13489|RINI_HUMAN
Q9UL46|PSME2_HUMAN
P36955|PEDF_HUMAN
P60981|DEST_HUMAN
P62913|RL11_HUMAN
O75888|TNF13_HUMAN
P57723|PCBP4_HUMAN
Q15365|PCBP1_HUMAN
P57721|PCBP3_HUMAN
Q15366|PCBP2_HUMAN
P15309|PPAP_HUMAN
P62491|RB11A_HUMAN
Q15907|RB11B_HUMAN
Q13867|BLMH_HUMAN
P33241|LSP1_HUMAN
P27635|RL10_HUMAN
O75083|WDR1_HUMAN
P09467|F16P1_HUMAN
P02452|CO1A1_HUMAN
P18124|RL7_HUMAN
P62899|RL31_HUMAN

P25789|PSA4_HUMAN
Q9UN76|S6A14_HUMAN
P06748|NPM_HUMAN
O43516|WIPF1_HUMAN
Q99497|PARK7_HUMAN
Q86X10|RLGPB_HUMAN
P40939|ECHA_HUMAN
P09668|CATH_HUMAN
O14497|ARI1A_HUMAN
Q13200|PSMD2_HUMAN
Q96SC8|DMTA2_HUMAN
Q14574|DSC3_HUMAN
Q9UPN9|TRI33_HUMAN
P08174|DAF_HUMAN
P06727|APOA4_HUMAN
Q5T1R4|ZEP3_HUMAN
Q96JM2|ZN462_HUMAN
Q96RY5|CRML_HUMAN
P04179|SODM_HUMAN

**Subject 2**

| MASTER+BS1BS1:CB440 | 258 | 262 | 262 | 259 | 260 | 262 | 261 | 261 | 260 |
|---|---|---|---|---|---|---|---|---|---|
| **Accession** | **Balafilcon A** | **Comfilcon A** | **Delefilcon A** | **Etafilcon A** | **Lotrafilcon B** | **Nelfilcon A** | **Nesofilcon A** | **Senofilcon A** | **Verofilcon A** |
| P02788\|TRFL_HUMAN | 5.644685 | 6.48063 | 6.093431 | 5.977888 | 5.404873 | 4.9491 | 5.29803 | 5.527466 | 8.567646 |
| P35527\|K1C9_HUMAN | 2.767031 | 2.743706 | 2.7649 | 2.727741 | 2.771539 | 2.762243 | 2.756836 | 2.762481 | 2.739341 |
| P04264\|K2C1_HUMAN | 4.436022 | 4.384302 | 4.213545 | 4.206775 | 4.333576 | 4.27544 | 4.402743 | 4.320868 | 4.318792 |
| P13645\|K1C10_HUMAN | 2.773293 | 2.47186 | 2.357781 | 2.628089 | 2.371568 | 2.477037 | 2.781749 | 2.361848 | 2.795523 |
| P35908\|K22E_HUMAN | 3.078768 | 2.591395 | 2.497434 | 2.930976 | 2.578695 | 2.753581 | 3.065732 | 2.852403 | 2.99409 |

| | | | | | | | | | |
|---|---|---|---|---|---|---|---|---|---|
| P61626\|LYSC_HUMAN | 6.048334 | 6.526546 | 6.697116 | 7.713213 | 5.062951 | 5.760944 | 7.566889 | 5.266971 | 7.339411 |
| P02768\|ALBU_HUMAN | 2.8478 | 3.790709 | 3.017425 | 3.453482 | 4.348905 | 3.664437 | 3.364637 | 2.851145 | 2.585931 |
| P31025\|LCN1_HUMAN | 5.578297 | 6.717542 | 5.48904 | 6.526489 | 5.842403 | 5.671717 | 5.741495 | 5.979437 | 5.699131 |
| P01833\|PIGR_HUMAN | 1.873089 | 2.630185 | 1.399917 | 2.252679 | 1.843134 | 2.014817 | 1.597878 | 2.189736 | 1.742629 |
| P01876\|IGHA1_HUMAN | 2.599109 | 3.228501 | 1.963902 | 2.765867 | 2.483815 | 2.613586 | 2.145384 | 2.797302 | 2.227979 |
| P01024\|CO3_HUMAN | -1.51409 | -0.51946 | -2.00516 | -0.91546 | -0.93243 | -0.92462 | -1.49667 | -1.47418 | -1.28412 |
| P25311\|ZA2G_HUMAN | 2.432156 | 3.351309 | 2.510742 | 3.065541 | 2.529098 | 2.838123 | 2.166594 | 2.751667 | 2.647599 |
| P04259\|K2C6B_HUMAN | -4.90984 | -3.65618 | -3.44926 | -3.79353 | -4.71527 | -4.91189 | -3.43927 | -5.18306 | -4.95241 |
| P19013\|K2C4_HUMAN | -0.84705 | 1.217862 | -0.94502 | 3.493818 | -0.78918 | -0.1876 | 1.104256 | 1.46855 | -0.92118 |
| P13647\|K2C5_HUMAN | 0.423783 | 1.180108 | 0.701091 | 2.721039 | 0.326963 | 0.483643 | 1.646154 | 1.186967 | 0.084229 |
| P02538\|K2C6A_HUMAN | | | | | | | | | |
| P08727\|K1C19_HUMAN | -2.87189 | -0.91089 | -2.59906 | 0.998334 | -2.36533 | -1.99987 | -0.64067 | -0.96458 | -2.84328 |
| P13646\|K1C13_HUMAN | -2.09237 | 0.256151 | -1.7979 | 2.59544 | -1.53998 | -0.88481 | 0.450621 | 0.544745 | -1.943 |
| P08779\|K1C16_HUMAN | -1.0241 | 0.327736 | 0.265874 | -0.07238 | -1.06433 | -1.23328 | 0.253113 | -1.57667 | -1.14315 |
| P02533\|K1C14_HUMAN | -0.2122 | -0.11209 | 0.031439 | 0.073408 | -0.62075 | -0.741 | 0.099115 | -0.37152 | -0.55042 |
| Q9UGM3\|DMBT1_HUMAN | -1.28902 | -0.57451 | -2.15561 | -1.00418 | -2.21463 | -1.49831 | -2.0417 | -1.51002 | -0.655 |
| P02787\|TRFE_HUMAN | -1.32497 | -0.27019 | -0.69682 | -0.33522 | -0.05334 | -0.58759 | -0.70354 | -1.25537 | -0.89971 |
| P0DOX7\|IGK_HUMAN | 1.372198 | 2.001336 | 0.917551 | 1.670605 | 1.369322 | 1.571072 | 1.14258 | 1.619624 | 1.259968 |
| P98160\|PGBM_HUMAN | -2.2288 | -1.75316 | -2.80896 | -1.41454 | -2.66245 | -2.21594 | -2.37559 | -2.39417 | -1.74311 |

| | | | | | | | | | |
|---|---|---|---|---|---|---|---|---|---|
| P60709\|ACTB_HUMAN | -1.68028 | -0.85546 | -1.53587 | -0.13485 | -1.44707 | -1.40734 | -1.71454 | -1.27949 | -1.7432 |
| P63261\|ACTG_HUMAN | -1.68028 | -0.85546 | -1.53587 | -0.13485 | -1.44707 | -1.40734 | -1.71454 | -1.27949 | -1.7432 |
| P15924\|DESP_HUMAN | -0.09018 | 0.140108 | 0.152834 | 0.620474 | -0.2453 | -0.18169 | 0.005607 | -0.13489 | -0.20525 |
| P06733\|ENOA_HUMAN | -1.35098 | -0.31273 | -0.44513 | -0.49115 | -0.75045 | -0.71021 | -1.14322 | -1.1627 | -1.52151 |
| P01036\|CYTS_HUMAN | 1.673771 | 2.664357 | 1.014903 | 2.081326 | 1.438902 | 1.801074 | 2.332546 | 1.772268 | 1.504994 |
| P12273\|PIP_HUMAN | 1.787567 | 3.547882 | 2.673246 | 3.493162 | 2.713623 | 3.206077 | 2.51323 | 3.08396 | 3.091125 |
| Q13421\|MSLN_HUMAN | -0.21506 | 0.39987 | -1.06696 | 0.504967 | -0.22428 | 0.109976 | -0.5661 | -0.02246 | -0.10503 |
| P01037\|CYTN_HUMAN | -1.04155 | -0.02197 | -1.82018 | -0.68586 | -1.33237 | -0.8218 | 0.288115 | -1.02389 | -1.15816 |
| P0DOX2\|IGA2_HUMAN | -3.27986 | -2.54846 | -3.77971 | -2.81424 | -3.26913 | -3.18006 | -3.53508 | -2.96269 | -3.45217 |
| P07355\|ANXA2_HUMAN | -1.10167 | 0.088107 | -0.73487 | 1.328026 | -0.05478 | -0.51442 | -0.16396 | -0.17276 | -1.20527 |
| P00450\|CERU_HUMAN | -1.08398 | -1.22513 | -2.32886 | -0.89797 | -1.90128 | -1.7017 | -1.56616 | -1.69257 | -1.53728 |
| P06396\|GELS_HUMAN | -2.45076 | -1.89898 | -2.91194 | -1.55216 | -2.29695 | -2.22284 | -2.72167 | -2.506 | -2.57695 |
| P00738\|HPT_HUMAN | -3.63585 | -2.88588 | -3.62611 | -3.23159 | -2.49562 | -2.85286 | -3.62132 | -3.73575 | -3.73069 |
| P0DOY2\|IGLC2_HUMAN | -3.95952 | -3.6127 | -4.65383 | -3.80065 | -4.15232 | -4.01195 | -4.48538 | -3.99538 | -4.07281 |
| B9A064\|IGLL5_HUMAN | -1.71277 | -1.0116 | -2.3233 | -1.3341 | -1.6516 | -1.67042 | -1.99616 | -1.42376 | -1.84845 |
| P0DOX8\|IGL1_HUMAN | -1.71277 | -1.0116 | -2.3233 | -1.3341 | -1.6516 | -1.67042 | -1.99616 | -1.42376 | -1.84845 |
| P0DOX5\|IGG1_HUMAN | -1.45212 | -0.44148 | -0.91415 | -0.16174 | -0.38121 | -0.60717 | -0.37333 | -1.36935 | -0.62883 |
| P10909\|CLUS_HUMAN | -0.29484 | 0.24836 | -1.11741 | 0.378115 | -0.6512 | -0.05792 | -0.79715 | -0.07192 | -0.23469 |
| P08729\|K2C7_HUMAN | -3.46816 | -2.42507 | -3.49049 | -0.6562 | -2.61862 | -2.87241 | -2.11651 | -2.83432 | -3.79806 |
| P04083\|ANXA1_HUMAN | -2.45417 | -1.06282 | -1.91146 | 0.447318 | -1.07215 | -1.65833 | -0.9889 | -1.22419 | -1.98398 |

| | | | | | | | | | |
|---|---|---|---|---|---|---|---|---|---|
| P60174\|TPIS_HUMAN | -3.96358 | -2.95834 | -2.56056 | -2.90629 | -2.87327 | -2.99782 | -3.64193 | -3.3939 | -4.68376 |
| P14923\|PLAK_HUMAN | -0.08951 | 0.110752 | 0.06473 | 0.233927 | -0.23665 | -0.22192 | -0.11518 | -0.0155 | -0.28578 |
| P00352\|AL1A1_HUMAN | -6.97835 | -6.17881 | -6.08349 | -5.99133 | -6.3822 | -6.35897 | -6.6226 | -6.80718 | -7.00306 |
| P01871\|IGHM_HUMAN | -1.91298 | -0.78771 | -2.49821 | -1.6053 | -1.79053 | -1.61036 | -2.02652 | -1.65778 | -1.8845 |
| P14618\|KPYM_HUMAN | -2.87086 | -2.27689 | -2.6555 | -2.20074 | -3.01214 | -2.9717 | -3.15282 | -3.24392 | -3.94036 |
| P05787\|K2C8_HUMAN | -7.03801 | -5.09182 | -6.35486 | -2.92717 | -6.91777 | -5.74161 | -4.19199 | -4.87529 | -8.06203 |
| O75556\|SG2A1_HUMAN | 1.378868 | 1.925618 | 1.068987 | 3.256906 | 0.939005 | 2.025649 | 1.975582 | 1.823692 | 1.77353 |
| P30838\|AL3A1_HUMAN | -4.0715 | -3.15118 | -3.04131 | -2.86536 | -3.66239 | -3.46313 | -3.2128 | -3.80783 | -2.84348 |
| P0DMV8\|HS71A_HUMAN | -4.51857 | -3.79781 | -4.33048 | -3.17538 | -4.0476 | -4.24685 | -3.99136 | -4.03436 | -5.11322 |
| P0DMV9\|HS71B_HUMAN | -4.51857 | -3.79781 | -4.33048 | -3.17538 | -4.0476 | -4.24685 | -3.99136 | -4.03436 | -5.11322 |
| Q08380\|LG3BP_HUMAN | -2.15835 | -1.15496 | -2.46874 | -0.89248 | -2.07774 | -1.87867 | -1.8641 | -1.34784 | -1.55492 |
| Q16378\|PROL4_HUMAN | 2.629207 | 3.698001 | 2.218897 | 3.854139 | 3.154548 | 3.330334 | 3.443252 | 2.857933 | 3.655827 |
| Q04695\|K1C17_HUMAN | -2.03527 | -0.86561 | -0.9908 | -0.93438 | -1.95875 | -2.0235 | -0.67201 | -2.24994 | -0.19053 |
| P68133\|ACTS_HUMAN | | | | | | | | | |
| P68032\|ACTC_HUMAN | | | | | | | | | |
| P04792\|HSPB1_HUMAN | -1.69828 | -1.0015 | -1.24841 | -0.37762 | -1.05892 | -1.66204 | -1.28061 | -1.24158 | -1.77573 |
| P01009\|A1AT_HUMAN | -2.70756 | -1.99198 | -2.53909 | -2.20573 | -1.85892 | -2.31746 | -2.86108 | -2.74299 | -3.12715 |
| P12035\|K2C3_HUMAN | -3.92167 | -4.05976 | -5.09638 | -4.69795 | -7.04664 | -2.52367 | -4.89626 | -3.20044 | -4.17452 |
| P06702\|S10A9_HUMAN | -0.82622 | -0.11437 | -0.26323 | -0.10302 | -0.0404 | -0.74337 | -0.91919 | -0.63551 | -1.3726 |
| P22079\|PERL_HUMAN | -3.28722 | -2.26227 | -3.52918 | -2.17374 | -3.0738 | -2.84544 | -3.03493 | -2.96317 | -2.33361 |
| P01859\|IGHG2_HUMAN | -5.22339 | -4.00489 | -4.80035 | -4.67087 | -3.52118 | -3.98933 | -5.06136 | -5.22296 | -4.89706 |

| | | | | | | | | | |
|---|---|---|---|---|---|---|---|---|---|
| P20061\|TCO1_HUMAN | -1.99674 | -0.7597 | -2.15411 | -0.89106 | -1.60081 | -1.23249 | -1.7196 | -1.34659 | -1.48863 |
| Q02413\|DSG1_HUMAN | -1.15487 | -1.07689 | -1.19763 | -1.06183 | -1.53836 | -1.23407 | -1.16581 | -1.0757 | -1.30648 |
| P19012\|K1C15_HUMAN | | | | | | | | | |
| P30740\|ILEU_HUMAN | -2.26739 | -1.38575 | -1.46359 | -1.43176 | -1.74725 | -1.99281 | -1.78752 | -1.93854 | -2.64822 |
| P80188\|NGAL_HUMAN | -1.90321 | -0.80315 | -1.24198 | -0.934 | -0.97624 | -1.09198 | -1.43914 | -1.52624 | -1.31007 |
| Q99935\|PROL1_HUMAN | 0.373643 | 0.515045 | -0.71502 | 0.717137 | -0.79343 | 0.025428 | -0.7377 | 0.025538 | 0.67665 |
| P04406\|G3P_HUMAN | -4.85448 | -3.69051 | -3.54094 | -3.88655 | -4.5912 | -4.22906 | -4.45774 | -4.15334 | -3.11141 |
| P03973\|SLPI_HUMAN | -0.00745 | -0.10723 | 0.652627 | 1.875622 | -0.90086 | -1.01466 | 1.040857 | -1.09155 | -0.51143 |
| P98088\|MUC5A_HUMAN | | | | | | | | | |
| P14555\|PA2GA_HUMAN | 1.139439 | 0.103566 | -1.53472 | 0.795659 | -1.15275 | -2.23551 | -0.27264 | -0.71926 | -1.12419 |
| P01591\|IGJ_HUMAN | 2.602039 | 3.16467 | 1.936933 | 2.650483 | 2.352173 | 2.507502 | 2.129472 | 2.774271 | 2.357535 |
| O95968\|SG1D1_HUMAN | 3.403019 | 3.676479 | 2.059463 | 4.365015 | 2.391109 | 3.366912 | 3.292515 | 3.676723 | 3.323772 |
| Q7Z794\|K2C1B_HUMAN | -2.25886 | -2.49607 | -2.81941 | -2.73425 | -2.65949 | -2.7683 | -2.4858 | -2.89038 | -2.14404 |
| Q8N1N4\|K2C78_HUMAN | -2.61985 | -2.42434 | -2.887 | -1.62961 | -2.86665 | -2.80934 | -2.31612 | -2.65254 | -2.74103 |
| P63104\|1433Z_HUMAN | -3.26407 | -2.65932 | -2.75807 | -2.29554 | -2.85384 | -2.86876 | -3.19585 | -3.01522 | -4.40163 |
| P31944\|CASPE_HUMAN | -4.32054 | -4.33271 | -4.35362 | -4.34684 | -4.35247 | -4.43614 | -4.50888 | -4.3373 | -4.67773 |
| Q96P63\|SPB12_HUMAN | -3.94711 | -4.03626 | -3.33119 | -2.90146 | -4.52016 | -4.11305 | -2.99931 | -4.29591 | -3.31733 |
| P09228\|CYTT_HUMAN | -3.5896 | -2.46113 | -4.30754 | -3.18222 | -3.74492 | -3.42134 | -2.89878 | -3.66262 | -3.9489 |
| P09211\|GSTP1_HUMAN | -3.19793 | -2.20636 | -2.08623 | -2.31255 | -2.36541 | -2.19626 | -2.95343 | -2.5657 | -3.67738 |
| Q9GZZ8\|LACRT_HUMAN | 2.740472 | 3.170036 | 1.767726 | 4.101622 | 1.983699 | 3.226014 | 2.1001 | 3.066889 | 2.659902 |

| | | | | | | | | | |
|---|---|---|---|---|---|---|---|---|---|
| P02647\|APOA1_HUMAN | -4.08951 | -3.62751 | -4.17617 | -2.89445 | -2.55776 | -2.80528 | -3.8254 | -3.87578 | -3.72193 |
| Q96DA0\|ZG16B_HUMAN | -1.62763 | -0.65038 | -1.92939 | -1.00581 | -1.68351 | -1.66373 | -1.57194 | -1.29859 | -0.14399 |
| P05109\|S10A8_HUMAN | -1.04264 | 0.099812 | 0.091498 | 0.197266 | 0.176802 | -0.54041 | -0.64761 | -0.39432 | -1.53503 |
| P07602\|SAP_HUMAN | -2.80335 | -2.0001 | -3.28677 | -1.44048 | -2.74144 | -2.25252 | -2.59554 | -2.42533 | -2.38555 |
| P05090\|APOD_HUMAN | | | | | | | | | |
| P11142\|HSP7C_HUMAN | -5.93795 | -4.85885 | -6.03591 | -5.12531 | -5.61851 | -6.01439 | -6.03839 | -6.1863 | -5.25254 |
| Q5D862\|FILA2_HUMAN | -2.70228 | -2.8529 | -2.79638 | -2.71428 | -2.96121 | -3.00052 | -2.63062 | -2.95945 | -2.76573 |
| P61769\|B2MG_HUMAN | -3.61403 | -2.82262 | -3.42675 | -2.60614 | -3.25344 | -3.26084 | -1.06619 | -3.20157 | -2.87063 |
| Q01469\|FABP5_HUMAN | | | | | | | | | |
| P34096\|RNAS4_HUMAN | -3.04251 | -2.18398 | -2.74553 | -1.326 | -3.21638 | -2.91184 | -1.94964 | -3.04807 | -2.21153 |
| P07339\|CATD_HUMAN | -3.75435 | -3.51573 | -4.04014 | -2.39726 | -3.38697 | -3.5719 | -3.34621 | -2.90807 | -3.90044 |
| P30044\|PRDX5_HUMAN | -7.57058 | -7.05324 | -6.9499 | -6.72597 | -7.18935 | -7.17565 | -7.45487 | -6.74686 | -4.70574 |
| P04075\|ALDOA_HUMAN | | | | | | | | | |
| P31947\|1433S_HUMAN | -8.7286 | -8.0668 | -8.03227 | -7.91549 | -7.93493 | -7.94259 | -8.56552 | -7.78342 | -9.58834 |
| Q5VTE0\|EF1A3_HUMAN | -3.4592 | -2.75905 | -1.91804 | -1.97859 | -3.5599 | -3.3613 | -3.07845 | -3.10718 | -3.59262 |
| P68104\|EF1A1_HUMAN | -3.4592 | -2.75905 | -1.91804 | -1.97859 | -3.5599 | -3.3613 | -3.07845 | -3.10718 | -3.59262 |
| P62937\|PPIA_HUMAN | -4.02542 | -2.36508 | -2.8784 | -2.27431 | -2.91055 | -2.56756 | -3.03314 | -3.04629 | -3.1156 |
| P62805\|H4_HUMAN | -3.56256 | -2.39084 | -3.75454 | -0.23141 | -3.63627 | -3.15166 | -1.92611 | -2.51741 | -4.54409 |
| P07737\|PROF1_HUMAN | -9.05087 | -7.23484 | -7.53566 | -7.80912 | -8.89478 | -8.75115 | -7.35628 | -8.20691 | -6.93831 |
| A0M8Q6\|IGLC7_HUMAN | -3.75494 | -2.88963 | -4.04226 | -3.28725 | -3.53407 | -3.70944 | -3.70557 | -3.43078 | -3.92577 |
| Q99456\|K1C12_HUMAN | -4.86072 | -5.45489 | -6.342 | -6.03028 | -7.99881 | -3.96988 | -5.48752 | -4.70912 | -4.85192 |
| Q06830\|PRDX1_HUMAN | -2.68563 | -2.18624 | -1.63455 | -1.63817 | -2.49256 | -2.50766 | -2.54816 | -2.48319 | -3.30826 |

| | | | | | | | | | |
|---|---|---|---|---|---|---|---|---|---|
| P30086\|PEBP1_HUMAN | -6.54923 | -5.27878 | -5.12286 | -5.85175 | -5.70406 | -5.71744 | -6.05424 | -6.03939 | -6.00143 |
| Q3SY84\|K2C71_HUMAN | | | | | | | | | |
| P02545\|LMNA_HUMAN | -5.76165 | -5.09948 | -5.64691 | -2.71867 | -5.81297 | -5.77604 | -4.25156 | -4.83753 | -5.5724 |
| P00338\|LDHA_HUMAN | | | | | | | | | |
| P01034\|CYTC_HUMAN | -3.20424 | -2.48767 | -3.53325 | -2.08336 | -3.41005 | -2.80281 | -1.46504 | -3.25749 | -3.36496 |
| P68363\|TBA1B_HUMAN | -7.33328 | -6.14605 | -5.61889 | -5.61132 | -6.90767 | -6.60486 | -7.25839 | -6.52109 | -7.93212 |
| P52209\|6PGD_HUMAN | -6.85425 | -5.75065 | -5.94496 | -5.91607 | -6.81849 | -6.43492 | -6.80925 | -6.60953 | -5.36007 |
| Q8N474\|SFRP1_HUMAN | -1.93762 | -1.77957 | -2.10606 | -2.20766 | -2.47829 | -2.83518 | -3.31619 | -2.43887 | -0.27591 |
| P01011\|AACT_HUMAN | -4.57803 | -3.56693 | -4.79484 | -3.55951 | -4.21433 | -4.21162 | -4.50566 | -4.49587 | -3.97301 |
| P17931\|LEG3_HUMAN | -4.01362 | -3.56553 | -4.17359 | -3.00765 | -4.24258 | -3.70761 | -3.75596 | -4.17236 | -4.31421 |
| P01619\|KV320_HUMAN | -2.8913 | -2.05378 | -3.25251 | -2.53437 | -2.95266 | -2.8201 | -3.16572 | -2.82306 | -3.1793 |
| P11021\|BIP_HUMAN | | | | | | | | | |
| P00558\|PGK1_HUMAN | | | | | | | | | |
| P07858\|CATB_HUMAN | -4.64401 | -4.05908 | -4.54329 | -3.14816 | -4.36361 | -4.40501 | -4.38203 | -4.38696 | -4.23835 |
| P29401\|TKT_HUMAN | -4.91486 | -3.72346 | -3.93347 | -3.73525 | -3.5982 | -4.33464 | -4.47568 | -4.5163 | -4.82459 |
| P80303\|NUCB2_HUMAN | -2.31221 | -1.74943 | -3.54961 | -1.41106 | -2.82256 | -2.08309 | -2.80051 | -2.26274 | -2.27322 |
| P15311\|EZRI_HUMAN | -4.07399 | -3.4161 | -3.82236 | -2.5067 | -4.07754 | -3.84371 | -3.84523 | -3.63362 | -4.6417 |
| Q08554\|DSC1_HUMAN | -2.54524 | -2.46525 | -2.26747 | -2.25551 | -2.60543 | -2.44216 | -2.3469 | -2.67772 | -2.30088 |
| Q93079\|H2B1H_HUMAN | -3.09656 | -2.2951 | -3.42596 | 0.169944 | -3.47398 | -2.85986 | -1.58586 | -2.11349 | -4.72906 |
| Q5QNW6\|H2B2F_HUMAN | -3.09656 | -2.2951 | -3.42596 | 0.169944 | -3.47398 | -2.85986 | -1.58586 | -2.11349 | -4.72906 |
| Q99877\|H2B1N_HUMAN | -3.09656 | -2.2951 | -3.42596 | 0.169944 | -3.47398 | -2.85986 | -1.58586 | -2.11349 | -4.72906 |

| | | | | | | | | | |
|---|---|---|---|---|---|---|---|---|---|
| O60814\|H2B1K_HUMAN | -3.09656 | -2.2951 | -3.42596 | 0.169944 | -3.47398 | -2.85986 | -1.58586 | -2.11349 | -4.72906 |
| P62807\|H2B1C_HUMAN | -3.09656 | -2.2951 | -3.42596 | 0.169944 | -3.47398 | -2.85986 | -1.58586 | -2.11349 | -4.72906 |
| P58876\|H2B1D_HUMAN | -3.09656 | -2.2951 | -3.42596 | 0.169944 | -3.47398 | -2.85986 | -1.58586 | -2.11349 | -4.72906 |
| Q99880\|H2B1L_HUMAN | -3.09656 | -2.2951 | -3.42596 | 0.169944 | -3.47398 | -2.85986 | -1.58586 | -2.11349 | -4.72906 |
| P57053\|H2BFS_HUMAN | -3.09656 | -2.2951 | -3.42596 | 0.169944 | -3.47398 | -2.85986 | -1.58586 | -2.11349 | -4.72906 |
| Q99879\|H2B1M_HUMAN | -3.09656 | -2.2951 | -3.42596 | 0.169944 | -3.47398 | -2.85986 | -1.58586 | -2.11349 | -4.72906 |
| P27797\|CALR_HUMAN | -8.01761 | -6.8962 | -6.81224 | -5.96565 | -6.50505 | -6.90615 | -6.94246 | -7.47916 | -9.2837 |
| Q6UXB2\|CXL17_HUMAN | -1.63304 | -1.92634 | -0.98942 | -0.29777 | -3.06961 | -2.39602 | -2.19723 | -1.87241 | -1.38537 |
| A0A0B4J1X5\|HV374_HUMAN | -4.3052 | -3.03999 | -4.51325 | -3.87347 | -4.03914 | -3.9945 | -4.30317 | -4.11183 | -4.28621 |
| P06744\|G6PI_HUMAN | -7.41049 | -7.28605 | -6.48306 | -7.38829 | -6.90467 | -6.91108 | -7.44318 | -7.23629 | -7.31589 |
| P05783\|K1C18_HUMAN | -3.8259 | -3.79595 | -4.76641 | -4.18104 | -5.69169 | -3.20481 | -4.33113 | -3.5517 | -4.00871 |
| P01040\|CYTA_HUMAN | -6.22753 | -5.93206 | -5.27813 | -6.37859 | -6.37617 | -5.90137 | -6.52378 | -5.96185 | -6.17407 |
| P40394\|ADH7_HUMAN | | | | | | | | | |
| A0A0B4J1V0\|HV315_HUMAN | | | | | | | | | |
| P80748\|LV321_HUMAN | -4.3328 | -3.62857 | -4.32415 | -3.71282 | -4.12178 | -4.21482 | -4.29745 | -3.948 | -4.47178 |
| P62979\|RS27A_HUMAN | -3.32143 | -2.99934 | -3.3064 | -2.2145 | -3.47555 | -3.2169 | -3.15488 | -3.03729 | -3.78072 |
| P62987\|RL40_HUMAN | -3.32143 | -2.99934 | -3.3064 | -2.2145 | -3.47555 | -3.2169 | -3.15488 | -3.03729 | -3.78072 |
| P0CG47\|UBB_HUMAN | -3.32143 | -2.99934 | -3.3064 | -2.2145 | -3.47555 | -3.2169 | -3.15488 | -3.03729 | -3.78072 |
| P0CG48\|UBC_HUMAN | -3.32143 | -2.99934 | -3.3064 | -2.2145 | -3.47555 | -3.2169 | -3.15488 | -3.03729 | -3.78072 |
| P16403\|H12_HUMAN | | | | | | | | | |
| P27348\|1433T_HUMAN | -7.54949 | -7.38668 | -7.43786 | -6.95956 | -7.82178 | -7.30544 | -8.07236 | -7.93493 | -7.90675 |
| P07237\|PDIA1_HUMAN | -5.50881 | -4.72194 | -5.80477 | -4.03654 | -5.09328 | -5.55623 | -5.04945 | -5.17542 | -6.03105 |

| | | | | | | | | | |
|---|---|---|---|---|---|---|---|---|---|
| P00751\|CFAB_HUMAN | -4.83629 | -3.99068 | -4.68382 | -3.59202 | -4.33559 | -4.36828 | -4.47666 | -4.70346 | -4.25816 |
| P06312\|KV401_HUMAN | -4.99956 | -4.16459 | -5.61335 | -4.70859 | -4.83591 | -4.80126 | -5.17777 | -4.61425 | -5.19094 |
| P02765\|FETUA_HUMAN | -6.15897 | -6.32132 | -6.95478 | -5.8656 | -6.44744 | -6.19753 | -6.82747 | -6.8604 | -6.87279 |
| P10412\|H14_HUMAN | -6.97356 | -5.70118 | -6.7559 | -4.02375 | -6.79607 | -6.31359 | -5.55048 | -6.01649 | -8.61271 |
| P31941\|ABC3A_HUMAN | -4.05254 | -4.03633 | -3.76587 | -3.64682 | -4.61031 | -4.21874 | -3.44855 | -4.20827 | -4.29985 |
| P55058\|PLTP_HUMAN | -4.14351 | -4.12887 | -5.59255 | -4.15863 | -5.95863 | -3.88149 | -5.13689 | -4.09753 | -4.48095 |
| P01615\|KVD28_HUMAN | -9.89306 | -8.03161 | -9.56396 | | -8.71988 | -9.04574 | -9.58374 | -8.33379 | -9.01603 |
| A0A075B6P5\|KV228_HUMAN | -9.89306 | -8.03161 | -9.56396 | | -8.71988 | -9.04574 | -9.58374 | -8.33379 | -9.01603 |
| P81605\|DCD_HUMAN | -1.44531 | -1.09814 | -1.89287 | -1.0491 | -1.89717 | -1.5677 | -1.82106 | -1.26955 | -1.5501 |
| P02774\|VTDB_HUMAN | | | | | | | | | |
| P14550\|AK1A1_HUMAN | | | | | | | | | |
| A0A0B4J1Y9\|HV372_HUMAN | | | | | | | | | |
| P32119\|PRDX2_HUMAN | -4.48851 | -4.558 | -4.28797 | -4.44671 | -4.63341 | -4.59216 | -4.42714 | -4.40367 | -4.57194 |
| P26447\|S10A4_HUMAN | -2.12036 | -1.07603 | -2.14628 | 0.482423 | -1.48022 | -1.77335 | -1.28134 | -1.32087 | -3.36325 |
| P31151\|S10A7_HUMAN | | | | | | | | | |
| Q15517\|CDSN_HUMAN | -4.39066 | -4.2852 | -4.12341 | -4.28768 | -4.60994 | -4.49695 | -4.40302 | -4.31269 | -4.00891 |
| P20930\|FILA_HUMAN | -2.74606 | -2.98885 | -2.79404 | -2.75008 | -2.85591 | -2.93222 | -2.94177 | -3.31425 | -2.81324 |
| P0DP08\|HVD82_HUMAN | -5.17815 | -3.55752 | -4.9783 | -3.88277 | -4.31272 | -4.08661 | -4.47 | -3.77851 | -4.43941 |
| P01825\|HV459_HUMAN | -5.17815 | -3.55752 | -4.9783 | -3.88277 | -4.31272 | -4.08661 | -4.47 | -3.77851 | -4.43941 |
| P0DP07\|HV431_HUMAN | -5.17815 | -3.55752 | -4.9783 | -3.88277 | -4.31272 | -4.08661 | -4.47 | -3.77851 | -4.43941 |
| A0A0C4DH41\|HV461_HUMAN | -5.17815 | -3.55752 | -4.9783 | -3.88277 | -4.31272 | -4.08661 | -4.47 | -3.77851 | -4.43941 |

| | | | | | | | | | |
|---|---|---|---|---|---|---|---|---|---|
| P0DP06\|HVD34_HUMAN | -5.17815 | -3.55752 | -4.9783 | -3.88277 | -4.31272 | -4.08661 | -4.47 | -3.77851 | -4.43941 |
| P06331\|HV434_HUMAN | -5.17815 | -3.55752 | -4.9783 | -3.88277 | -4.31272 | -4.08661 | -4.47 | -3.77851 | -4.43941 |
| P01824\|HV439_HUMAN | -5.17815 | -3.55752 | -4.9783 | -3.88277 | -4.31272 | -4.08661 | -4.47 | -3.77851 | -4.43941 |
| A0A0A0MRZ8\|KVD11_HUMAN | -3.62382 | -2.75842 | -3.82575 | -3.21733 | -3.45199 | -3.37961 | -4.02585 | -3.21023 | -3.83798 |
| P04433\|KV311_HUMAN | -3.62382 | -2.75842 | -3.82575 | -3.21733 | -3.45199 | -3.37961 | -4.02585 | -3.21023 | -3.83798 |
| P31949\|S10AB_HUMAN | -6.49841 | -5.64461 | -6.43327 | -3.3646 | -5.46659 | -5.8031 | -5.54138 | -5.40974 | -4.38652 |
| P60953\|CDC42_HUMAN | | | | | | | | | |
| Q6KB66\|K2C80_HUMAN | | | | | | | | | |
| P10599\|THIO_HUMAN | -3.98922 | -3.45196 | -3.886 | -3.59964 | -3.70098 | -3.53439 | -2.85072 | -3.88333 | -4.26743 |
| P40925\|MDHC_HUMAN | | | | | | | | | |
| Q14515\|SPRL1_HUMAN | -3.60219 | -3.15856 | -3.1492 | -2.92998 | -3.80616 | -3.54433 | -3.48561 | -3.54795 | -1.69571 |
| A0A075B6S5\|KV127_HUMAN | -5.67315 | -5.2259 | -6.4338 | -5.4315 | -6.38825 | -5.42701 | -4.46467 | -5.54763 | -6.00711 |
| P01742\|HV169_HUMAN | -2.66521 | -1.85928 | -3.25885 | -2.1369 | -2.47611 | -2.42676 | -2.96573 | -2.37178 | -2.69849 |
| A0A0C4DH31\|HV118_HUMAN | -5.49806 | -5.04781 | -6.87507 | -5.65591 | -5.90505 | -5.73998 | -6.27927 | -5.55426 | -6.307 |
| P23083\|HV102_HUMAN | -5.49806 | -5.04781 | -6.87507 | -5.65591 | -5.90505 | -5.73998 | -6.27927 | -5.55426 | -6.307 |
| P07900\|HS90A_HUMAN | | | | | | | | | |
| P84243\|H33_HUMAN | -5.18693 | -4.22632 | -5.81802 | -2.52882 | -5.9327 | -4.99728 | -4.11258 | -4.36105 | -6.89592 |
| P01700\|LV147_HUMAN | -4.2826 | -3.68311 | -4.79292 | -3.98263 | -4.47394 | -4.36898 | -4.61047 | -4.23752 | -4.83324 |
| P23528\|COF1_HUMAN | -4.26877 | -3.87108 | -4.16657 | -3.68555 | -4.07095 | -4.043 | -4.13036 | -4.37264 | -5.44196 |
| P21980\|TGM2_HUMAN | | | | | | | | | |
| Q09666\|AHNK_HUMAN | -4.67993 | -3.99446 | -4.8216 | -1.98413 | -4.50683 | -4.69093 | -4.04517 | -3.97776 | -5.49348 |
| P01624\|KV315_HUMAN | -5.46325 | -4.9535 | -6.3038 | -5.30014 | -5.66796 | -5.59507 | -5.97791 | -5.35638 | -5.9798 |

| | | | | | | | | | |
|---|---|---|---|---|---|---|---|---|---|
| P07384\|CAN1_HUMAN | -8.11176 | -7.70881 | -7.27626 | -7.76558 | -8.13608 | -8.53675 | -8.18625 | -7.9899 | -5.11624 |
| Q6UWP8\|SBSN_HUMAN | -7.26088 | -9.00928 | -6.65446 | -6.49573 | -7.52108 | -7.49844 | -7.48813 | -8.02811 | -6.93709 |
| Q8NBJ4\|GOLM1_HUMAN | -3.95918 | -3.20113 | -4.58587 | -3.18636 | -4.04794 | -3.99843 | -3.95802 | -3.80273 | -4.30525 |
| P40926\|MDHM_HUMAN | | | | | | | | | |
| P68371\|TBB4B_HUMAN | | | | | | | | | |
| A0A0A0MS15\|HV349_HUMAN | -6.23233 | -4.66726 | -6.15526 | -5.38777 | -6.11189 | -5.74982 | -6.34585 | -5.39309 | -5.68988 |
| P01593\|KVD33_HUMAN | -5.27848 | -4.93275 | -4.97787 | -4.96577 | -5.94854 | -6.28293 | -5.89557 | -5.43136 | -2.4922 |
| P01594\|KV133_HUMAN | -5.27848 | -4.93275 | -4.97787 | -4.96577 | -5.94854 | -6.28293 | -5.89557 | -5.43136 | -2.4922 |
| P02766\|TTHY_HUMAN | | | | | | | | | |
| Q08188\|TGM3_HUMAN | -6.18664 | -6.85132 | -6.74376 | -6.44831 | -6.78445 | -6.17526 | -6.24436 | -6.77573 | -6.44642 |
| P05089\|ARGI1_HUMAN | -5.08345 | -5.54309 | -5.84305 | -6.01484 | -5.84217 | -5.71381 | -5.56698 | -5.33983 | -5.74978 |
| Q9UBT3\|DKK4_HUMAN | -4.61828 | -5.37693 | -5.42407 | -5.08332 | -5.31641 | -6.34033 | -5.81898 | -5.68149 | -3.36796 |
| O00584\|RNT2_HUMAN | -7.36287 | -5.86128 | -7.06823 | -6.07311 | -6.76414 | -6.37973 | -7.00827 | -6.43829 | -6.59461 |
| A0A0B4J2D9\|KVD13_HUMAN | | | | | | | | | |
| P0DP09\|KV113_HUMAN | | | | | | | | | |
| O75874\|IDHC_HUMAN | | | | | | | | | |
| P15814\|IGLL1_HUMAN | -1.2897 | -0.64156 | -1.96492 | -0.81818 | -1.42151 | -1.34051 | -1.57917 | -0.97816 | -1.52283 |
| P30085\|KCY_HUMAN | | | | | | | | | |
| A0A0C4DH34\|HV428_HUMAN | -5.92066 | -4.63227 | -5.48728 | -4.89509 | -5.42763 | -5.11199 | -5.51386 | -5.09837 | -5.69898 |
| P30101\|PDIA3_HUMAN | -6.61721 | -5.81353 | -5.93559 | -4.59006 | -5.57371 | -5.84301 | -5.54271 | -5.78894 | -6.59167 |
| Q14697\|GANAB_HUMAN | | | | | | | | | |
| P12830\|CADH1_HUMAN | -5.44358 | -5.75053 | -6.04007 | -5.02103 | -6.52047 | -7.91992 | -5.13407 | -6.54863 | -4.76669 |

| | | | | | | | | | |
|---|---|---|---|---|---|---|---|---|---|
| Q16651\|PRSS8_HUMAN | -3.72599 | -3.18484 | -4.26402 | -3.31165 | -3.4568 | -3.66928 | -4.33152 | -3.58223 | -4.12473 |
| P13987\|CD59_HUMAN | -5.38566 | -4.87716 | -5.7195 | -4.31926 | -5.8659 | -5.50629 | -5.52724 | -5.31089 | -5.77939 |
| O43653\|PSCA_HUMAN | -5.49089 | -5.16512 | -5.17474 | -3.33056 | -5.79703 | -4.76615 | -5.39521 | -4.99047 | -5.56734 |
| P02763\|A1AG1_HUMAN | | | | | | | | | |
| A0A0B4J1V6\|HV373_HUMAN | -5.83353 | -5.47468 | -6.18378 | -5.68245 | -6.01039 | -5.6414 | -6.06715 | -5.70362 | -5.93903 |
| O43852\|CALU_HUMAN | -9.24874 | -8.37664 | -6.74344 | -7.75013 | -8.08858 | -8.30305 | -9.24476 | -9.25096 | -9.34513 |
| P08758\|ANXA5_HUMAN | | | | | | | | | |
| Q5T749\|KPRP_HUMAN | -5.18148 | -6.3469 | -6.24756 | -6.04658 | -6.29434 | -6.85159 | -5.98212 | -6.13587 | -6.08766 |
| Q7Z5P9\|MUC19_HUMAN | -4.61507 | -4.05332 | -4.08289 | -3.46831 | -4.8003 | -4.82876 | -3.77325 | -4.71829 | -2.38295 |
| P01714\|LV319_HUMAN | -5.29614 | -4.57128 | -5.46803 | -4.89671 | -5.39859 | -5.05133 | -5.43998 | -5.23346 | -5.35384 |
| P01717\|LV325_HUMAN | -4.87676 | -4.41881 | -5.82474 | -4.83486 | -5.15219 | -5.08055 | -5.46052 | -4.88312 | -5.17802 |
| A0A075B6K4\|LV310_HUMAN | -4.87676 | -4.41881 | -5.82474 | -4.83486 | -5.15219 | -5.08055 | -5.46052 | -4.88312 | -5.17802 |
| P23284\|PPIB_HUMAN | -4.06157 | -4.3104 | -4.83947 | -4.14589 | -4.52023 | -4.34697 | -4.36835 | -4.13568 | -4.05177 |
| P12814\|ACTN1_HUMAN | | | | | | | | | |
| P40199\|CEAM6_HUMAN | | | | | | | | | |
| Q02818\|NUCB1_HUMAN | -5.69124 | -5.52485 | -7.27724 | -5.15481 | -6.09189 | -5.54785 | -6.18925 | -5.71959 | -5.56485 |
| P49788\|TIG1_HUMAN | | | | | | | | | |
| P06703\|S10A6_HUMAN | -5.85421 | -4.79576 | -5.43012 | -4.41891 | -4.80464 | -4.83776 | -5.06646 | -5.57781 | -6.57036 |
| P16401\|H15_HUMAN | -6.33192 | -6.10909 | -6.68398 | -3.39384 | -6.99376 | -6.79095 | -5.46629 | -5.85368 | -7.82121 |
| O60437\|PEPL_HUMAN | -7.32376 | -6.80164 | -6.85193 | -5.10704 | -7.46575 | -7.08692 | -6.87395 | -6.67536 | -8.06299 |
| P47929\|LEG7_HUMAN | | | | | | | | | |
| Q14508\|WFDC2_HUMAN | -4.33381 | -6.35575 | -8.21145 | -7.96192 | -5.82737 | -7.73848 | -6.54284 | -8.3149 | -7.38706 |

| | | | | | | | | | |
|---|---|---|---|---|---|---|---|---|---|
| Q13835\|PKP1_HUMAN | | | | | | | | | |
| P02750\|A2GL_HUMAN | | | | | | | | | |
| P21926\|CD9_HUMAN | | | | | | | | | |
| P19961\|AMY2B_HUMAN | | | | | | | | | |
| P0DTE7\|AMY1B_HUMAN | | | | | | | | | |
| P04746\|AMYP_HUMAN | | | | | | | | | |
| P04745\|AMY1A_HUMAN | | | | | | | | | |
| P0DTE8\|AMY1C_HUMAN | | | | | | | | | |
| Q9UBC9\|SPRR3_HUMAN | -4.96292 | -3.88817 | -4.66907 | -2.93945 | -4.55817 | -4.48638 | -4.33264 | -4.49716 | -4.71037 |
| Q96QA5\|GSDMA_HUMAN | -6.41383 | -6.61264 | -6.94885 | -7.11929 | -6.75398 | -6.74054 | -6.9464 | -6.81629 | -6.52153 |
| Q13296\|SG2A2_HUMAN | | | | | | | | | |
| Q14002\|CEAM7_HUMAN | | | | | | | | | |
| P62753\|RS6_HUMAN | -6.46497 | -5.53206 | -6.54263 | -4.94145 | -6.76313 | -6.1819 | -5.93411 | -6.15132 | -6.82296 |
| P09429\|HMGB1_HUMAN | -4.61973 | -3.52519 | -5.50652 | -3.91226 | -4.90374 | -4.99179 | -4.90028 | -4.40279 | -6.92552 |
| P83731\|RL24_HUMAN | -3.64261 | -2.78345 | -3.9637 | -2.75841 | -3.54812 | -3.82614 | -3.43422 | -3.56745 | -5.51431 |
| Q96S96\|PEBP4_HUMAN | | | | | | | | | |
| Q5SNV9\|CA167_HUMAN | | | | | | | | | |
| Q6ZR08\|DYH12_HUMAN | | | | | | | | | |
| P07910\|HNRPC_HUMAN | | | | | | | | | |
| Q9UKZ1\|CNO11_HUMAN | | | | | | | | | |
| P20933\|ASPG_HUMAN | -6.00335 | -5.22531 | -6.17846 | -4.89334 | -5.38554 | -5.97185 | -5.66205 | -5.71521 | -7.01675 |
| Q8IZL8\|PELP1_HUMAN | | | | | | | | | |
| P00505\|AATM_HUMAN | | | | | | | | | |
| P48634\|PRC2A_HUMAN | | | | | | | | | |
| Q96G74\|OTUD5_HUMAN | | | | | | | | | |
| Q8IVF2\|AHNK2_HUMAN | | | | | | | | | |
| Q8TF72\|SHRM3_HUMAN | | | | | | | | | |
| Q14966\|ZN638_HUMAN | | | | | | | | | |
| Q7LBE3\|S26A9_HUMAN | | | | | | | | | |

| | | | | | | | | | |
|---|---|---|---|---|---|---|---|---|---|
| A6NKD9\|CC85C_HUMAN | | | | | | | | | |
| A0A0B4J1U7\|HV601_HUMAN | | | | | | | | | |
| Q6PID8\|KLD10_HUMAN | | | | | | | | | |
| Q99574\|NEUS_HUMAN | -8.52674 | -7.77156 | -7.93474 | -7.23589 | -8.2585 | -8.00804 | -7.8688 | -7.83724 | -8.03226 |
| Q14526\|HIC1_HUMAN | | | | | | | | | |
| Q5VSY0\|GKAP1_HUMAN | | | | | | | | | |
| Q8TCU6\|PREX1_HUMAN | | | | | | | | | |
| Q7Z2W7\|TRPM8_HUMAN | | | | | | | | | |
| Q14106\|TOB2_HUMAN | | | | | | | | | |
| Q9GZQ3\|COMD5_HUMAN | | | | | | | | | |
| Q8NHQ8\|RASF8_HUMAN | | | | | | | | | |
| P01703\|LV140_HUMAN | | | | | | | | | |
| O75882\|ATRN_HUMAN | | | | | | | | | |
| O60902\|SHOX2_HUMAN | | | | | | | | | |
| Q5VSP4\|LC1L1_HUMAN | -4.5276 | -3.86731 | -4.87759 | -3.7678 | -4.4547 | -4.50446 | -5.08391 | -4.49022 | -4.2846 |
| P01861\|IGHG4_HUMAN | -6.33406 | -5.08766 | -5.85803 | -4.98139 | -4.69056 | -4.96464 | -5.78652 | -5.9092 | -5.81118 |
| O95678\|K2C75_HUMAN | -4.27939 | -4.06688 | -4.93008 | -4.43863 | -6.14438 | -3.14709 | -4.49012 | -3.82082 | -4.07595 |
| P0DOX6\|IGM_HUMAN | -3.7822 | -3.23599 | -4.14569 | -2.98589 | -3.94393 | -3.60506 | -4.31534 | -3.69323 | -4.12857 |
| Q86YZ3\|HORN_HUMAN | -5.12948 | -5.74736 | -5.3587 | -5.60053 | -5.80899 | -5.06065 | -5.90778 | -5.80279 | -4.69507 |
| Q14525\|KT33B_HUMAN | -5.49679 | -6.03605 | -7.32674 | -6.83609 | -8.66626 | -5.57636 | -6.57689 | -5.33472 | -5.66064 |
| Q562R1\|ACTBL_HUMAN | -7.70596 | -5.5056 | -7.05125 | -6.71323 | -7.41089 | -6.63564 | -7.06945 | -6.7263 | -7.31477 |
| P31946\|1433B_HUMAN | -8.59244 | -8.25918 | -8.30963 | -8.08167 | -8.10245 | -7.97417 | -9.11546 | -8.59602 | -10.0516 |
| Q7Z3Y8\|K1C27_HUMAN | -3.93016 | -3.06254 | -3.99271 | -3.62046 | -4.27784 | -2.85182 | -3.76216 | -3.61096 | -3.944 |
| P01780\|HV307_HUMAN | -4.71971 | -4.28631 | -5.11316 | -4.52874 | -4.84144 | -4.76795 | -4.83208 | -4.58348 | -4.68604 |

| | | | | | | | | | |
|---|---|---|---|---|---|---|---|---|---|
| A0A0J9YX35\|HV64D_HUMAN | -4.34356 | -3.52108 | -4.82345 | -3.98193 | -4.3827 | -4.09228 | -4.80167 | -3.90306 | -4.47966 |
| A0A0J9YXX1\|HV5X1_HUMAN | -6.04259 | -4.74196 | -6.57115 | -5.13202 | -6.17185 | -5.60193 | -6.14068 | -5.90007 | -6.05779 |
| A0A0C4DH38\|HV551_HUMAN | -4.13295 | -3.68476 | -4.79969 | -4.11811 | -4.23194 | -4.43096 | -4.71867 | -4.35439 | -4.46492 |
| P28799\|GRN_HUMAN | -4.83206 | -4.2053 | -5.51538 | -4.13396 | -4.8907 | -4.53817 | -4.90358 | -4.7883 | -4.71494 |
| P01762\|HV311_HUMAN | -7.06962 | -6.44133 | -8.08236 | -6.8623 | -7.40071 | -7.31374 | -7.66004 | -6.74766 | -7.31228 |
| Q7Z3Y9\|K1C26_HUMAN | -4.29899 | -5.09886 | -6.32418 | -6.03684 | -6.97923 | -4.45437 | -6.00669 | -4.90989 | -4.27801 |
| Q15149\|PLEC_HUMAN | -6.00897 | -4.99072 | -5.02309 | -3.61484 | -5.79636 | -5.83895 | -4.74214 | -5.45662 | -5.62718 |
| Q7Z406\|MYH14_HUMAN | -9.23599 | -8.48729 | -9.41594 | -7.23875 | -9.20901 | -9.34018 | -8.27705 | -8.82427 | |
| P06731\|CEAM5_HUMAN | -4.90208 | -4.02343 | -5.46462 | -3.68337 | -5.9798 | -4.67267 | -3.89381 | -4.56851 | -4.80579 |
| P04632\|CPNS1_HUMAN | -7.65178 | -8.09023 | -7.81741 | -6.07824 | -7.88049 | -7.9177 | -7.166 | -7.39895 | -5.98068 |
| Q07666\|KHDR1_HUMAN | -6.48146 | -4.1332 | -3.70315 | -4.10789 | -5.4187 | -5.55449 | -5.76406 | -4.97548 | -4.73494 |
| O60716\|CTND1_HUMAN | | -3.98036 | -3.82668 | -4.09438 | -6.13449 | -5.26265 | -6.89705 | -5.10933 | |
| P01611\|KVD12_HUMAN | -8.20977 | -6.89057 | -8.69797 | -7.15567 | -8.25947 | -8.27202 | -7.8957 | -7.39672 | -7.48036 |
| A0A0C4DH72\|KV106_HUMAN | -6.39876 | -6.52472 | -7.82566 | -6.59772 | -7.22524 | -7.02059 | -7.2586 | -7.39859 | -7.53316 |
| P08246\|ELNE_HUMAN | -6.20688 | -7.3458 | -6.27442 | -5.8725 | -6.5735 | -7.4095 | -6.83358 | -7.94989 | -7.41833 |
| A0A075B6R9\|KVD24_HUMAN | -7.08838 | -6.09244 | -6.82254 | -6.57223 | -6.69268 | -6.67298 | -6.95909 | -6.42143 | -7.12268 |
| A0A0C4DH68\|KV224_HUMAN | -7.08838 | -6.09244 | -6.82254 | -6.57223 | -6.69268 | -6.67298 | -6.95909 | -6.42143 | -7.12268 |
| P40121\|CAPG_HUMAN | -5.30394 | -4.53232 | -5.04785 | -4.08616 | -4.91161 | -4.94835 | -4.99627 | -5.09653 | -5.90549 |
| O76021\|RL1D1_HUMAN | | -7.9904 | -6.90927 | -7.57424 | -9.47659 | -8.62442 | -9.42683 | -9.15607 | -10.2 |
| A0A075B6H7\|KV37_HUMAN | -6.68794 | -6.11383 | -7.38272 | -6.80342 | -7.13297 | -6.9296 | -7.62515 | -6.83478 | -7.29292 |

| | | | | | | | | | |
|---|---|---|---|---|---|---|---|---|---|
| P42357\|HUTH_HUMAN | -9.99312 | | -8.93402 | | | -9.94252 | -9.7031 | -9.26993 | -9.13606 |
| P13797\|PLST_HUMAN | -6.54708 | -6.47048 | -6.12106 | -6.17777 | -5.76501 | -6.43911 | -6.42112 | -6.66661 | -8.1963 |
| P04211\|LV743_HUMAN | -4.20688 | -3.36093 | -4.55042 | -3.86973 | -3.98216 | -3.99061 | -4.32686 | -3.85257 | -4.6045 |
| A0A075B6I9\|LV746_HUMAN | -4.20688 | -3.36093 | -4.55042 | -3.86973 | -3.98216 | -3.99061 | -4.32686 | -3.85257 | -4.6045 |
| P55064\|AQP5_HUMAN | -3.77447 | -3.78543 | -4.53367 | -2.7301 | -5.00583 | -4.21482 | -4.57637 | -4.02614 | -4.88084 |
| P07951\|TPM2_HUMAN | -6.11573 | -6.01546 | -6.30295 | -4.86639 | -6.25281 | -6.23608 | -5.69519 | -6.66511 | -6.56158 |
| P09493\|TPM1_HUMAN | -6.11573 | -6.01546 | -6.30295 | -4.86639 | -6.25281 | -6.23608 | -5.69519 | -6.66511 | -6.56158 |
| P22735\|TGM1_HUMAN | -6.19873 | -5.87317 | -6.6017 | -5.83789 | -6.65129 | -6.54469 | -6.19244 | -6.36834 | -6.09709 |
| P01701\|LV151_HUMAN | -5.61438 | -4.87909 | -5.08584 | -4.38356 | -5.84486 | -5.21811 | -4.72622 | -5.71521 | -4.78396 |
| P49840\|GSK3A_HUMAN | -7.20523 | -6.19377 | -6.13869 | -6.07937 | -6.44942 | -6.39957 | -6.57338 | -6.65002 | -5.92998 |
| P0DOX3\|IGD_HUMAN | -5.7669 | -4.16317 | -5.49522 | -4.67414 | -5.06453 | -4.68972 | -5.37472 | -4.85631 | -5.25898 |
| P10809\|CH60_HUMAN | | -8.20535 | -6.8819 | -8.01071 | -9.37645 | -9.38952 | -9.49389 | -9.44992 | |
| P02679\|FIBG_HUMAN | -6.86003 | -5.83224 | -6.80972 | -5.78279 | -5.87137 | -5.71254 | -6.87339 | -6.47273 | -5.59247 |
| Q9UPP2\|IQEC3_HUMAN | -5.22013 | -4.51923 | -5.02496 | -5.16431 | -5.12399 | -5.49564 | -5.88073 | -5.23526 | -2.44936 |
| A0A075B6I0\|LV861_HUMAN | -3.49525 | -2.52952 | -3.31874 | -2.91493 | -3.2434 | -3.09514 | -3.423 | -3.13773 | -3.90906 |
| Q86V81\|THOC4_HUMAN | -8.24703 | -7.9458 | -7.47535 | -7.5388 | -8.46695 | -7.58402 | -7.69348 | -8.27908 | -9.01438 |
| P22681\|CBL_HUMAN | -7.53727 | -6.87269 | -6.94633 | -7.06589 | -7.43734 | -7.6032 | -8.57561 | -7.99719 | -5.05779 |
| Q9BVC4\|LST8_HUMAN | | -8.95616 | | | | | | | |
| Q9BWS9\|CHID1_HUMAN | -10.0248 | -7.74539 | | -8.98007 | -9.01323 | -9.10672 | | | -6.35056 |
| Q7RTR2\|NLRC3_HUMAN | -6.60015 | -5.48042 | -7.37671 | -7.02116 | -7.15808 | -6.46518 | -7.30151 | -6.00478 | -1.72842 |

| | | | | | | | | | |
|---|---|---|---|---|---|---|---|---|---|
| Q8N6G6\|ATL1_HUMAN | -6.89804 | -6.07194 | -6.10256 | -6.05751 | -6.72836 | -6.44148 | -6.58628 | -6.74784 | -5.48071 |
| P67809\|YBOX1_HUMAN | -7.5985 | -7.06926 | -4.22303 | -6.40621 | -7.05964 | -7.39339 | -8.64999 | -6.84876 | -6.30241 |
| Q8IZ21\|PHAR4_HUMAN | -7.21164 | -6.88038 | -7.78519 | -6.74849 | -7.555 | -7.31959 | -7.18459 | -6.76771 | -6.49186 |
| Q8WXS5\|CCG8_HUMAN | | | -9.71987 | | | | | | -8.73639 |
| Q9ULD2\|MTUS1_HUMAN | -4.60612 | -4.07578 | -5.29717 | -5.01951 | -6.63726 | -4.66869 | -5.99067 | -4.1851 | -2.2853 |
| Q86SM8\|MRGRE_HUMAN | -4.85859 | -4.67493 | -5.85126 | -4.66818 | -5.03367 | -5.20659 | -6.02787 | -5.03181 | -5.06198 |
| Q13283\|G3BP1_HUMAN | | -8.25904 | -5.97818 | -7.41723 | -7.8649 | -8.86782 | -9.10117 | -8.7714 | -9.26291 |
| Q9NSA2\|KCND1_HUMAN | -7.09666 | -6.89081 | -7.88705 | -7.81238 | | -5.65148 | -7.74884 | -6.34201 | -6.43023 |
| Q15323\|K1H1_HUMAN | | | | | | | | | |
| O43790\|KRT86_HUMAN | | | | | | | | | |
| Q14533\|KRT81_HUMAN | | | | | | | | | |
| P62258\|1433E_HUMAN | | | | | | | | | |
| Q13228\|SBP1_HUMAN | | | | | | | | | |
| P78386\|KRT85_HUMAN | | | | | | | | | |
| P13929\|ENOB_HUMAN | | | | | | | | | |
| A0A0C4DH42\|HV366_HUMAN | | | | | | | | | |
| P02790\|HEMO_HUMAN | | | | | | | | | |
| P22392\|NDKB_HUMAN | | | | | | | | | |
| P37802\|TAGL2_HUMAN | | | | | | | | | |
| P29508\|SPB3_HUMAN | | | | | | | | | |
| P18510\|IL1RA_HUMAN | | | | | | | | | |
| P30041\|PRDX6_HUMAN | | | | | | | | | |
| P15259\|PGAM2_HUMAN | | | | | | | | | |
| P18669\|PGAM1_HUMAN | | | | | | | | | |
| P48594\|SPB4_HUMAN | | | | | | | | | |
| P0DP24\|CALM2_HUMAN | | | | | | | | | |
| P0DP23\|CALM1_HUMAN | | | | | | | | | |
| P0DP25\|CALM3_HUMAN | | | | | | | | | |

P05387|RLA2_HUMAN
P04080|CYTB_HUMAN
P22626|ROA2_HUMAN
O75223|GGCT_HUMAN
P37837|TALDO_HUMAN
P27482|CALL3_HUMAN
O00299|CLIC1_HUMAN
P08582|TRFM_HUMAN
Q9HC84|MUC5B_HUMAN
P0DME0|SETLP_HUMAN
Q01105|SET_HUMAN
P02511|CRYAB_HUMAN
P01008|ANT3_HUMAN
P08571|CD14_HUMAN
A0A075B6S9|KV137_HUMAN
P0DSN7|KVD37_HUMAN
P06454|PTMA_HUMAN
P08294|SODE_HUMAN
Q9HAV0|GBB4_HUMAN
P62873|GBB1_HUMAN
P62879|GBB2_HUMAN
P16520|GBB3_HUMAN
P25705|ATPA_HUMAN
P08670|VIME_HUMAN
P61158|ARP3_HUMAN
Q9H0U4|RAB1B_HUMAN
Q92928|RAB1C_HUMAN
P62820|RAB1A_HUMAN
Q92930|RAB8B_HUMAN
P61026|RAB10_HUMAN
P59190|RAB15_HUMAN
Q15286|RAB35_HUMAN
P51153|RAB13_HUMAN

P61006|RAB8A_HUMAN
P61160|ARP2_HUMAN
P06576|ATPB_HUMAN
P60660|MYL6_HUMAN
P14649|MYL6B_HUMAN
P05386|RLA1_HUMAN
Q14764|MVP_HUMAN
Q14118|DAG1_HUMAN
P04217|A1BG_HUMAN
P36952|SPB5_HUMAN
Q02809|PLOD1_HUMAN
Q6ZVX7|FBX50_HUMAN
P14174|MIF_HUMAN
Q58FF3|ENPLL_HUMAN
P14625|ENPL_HUMAN
P25786|PSA1_HUMAN
O95436|NPT2B_HUMAN
P13489|RINI_HUMAN
Q9UL46|PSME2_HUMAN
P36955|PEDF_HUMAN
P60981|DEST_HUMAN
P62913|RL11_HUMAN
O75888|TNF13_HUMAN
P57723|PCBP4_HUMAN
Q15365|PCBP1_HUMAN
P57721|PCBP3_HUMAN
Q15366|PCBP2_HUMAN
P15309|PPAP_HUMAN
P62491|RB11A_HUMAN
Q15907|RB11B_HUMAN
Q13867|BLMH_HUMAN
P33241|LSP1_HUMAN
P27635|RL10_HUMAN

O75083|WDR1_HUMAN
P09467|F16P1_HUMAN
P02452|CO1A1_HUMAN
P18124|RL7_HUMAN
P62899|RL31_HUMAN
P25789|PSA4_HUMAN
Q9UN76|S6A14_HUMAN
P06748|NPM_HUMAN
O43516|WIPF1_HUMAN
Q99497|PARK7_HUMAN
Q86X10|RLGPB_HUMAN
P40939|ECHA_HUMAN
P09668|CATH_HUMAN
O14497|ARI1A_HUMAN
Q13200|PSMD2_HUMAN
Q96SC8|DMTA2_HUMAN
Q14574|DSC3_HUMAN
Q9UPN9|TRI33_HUMAN
P08174|DAF_HUMAN
P06727|APOA4_HUMAN
Q5T1R4|ZEP3_HUMAN
Q96JM2|ZN462_HUMAN
Q96RY5|CRML_HUMAN
P04179|SODM_HUMAN

**Subject 3**

| MASTER | 357 | 351 | 358 | 356 | 0 | 357 | 355 | 351 | 358 |
|---|---|---|---|---|---|---|---|---|---|
| **Accession** | **Balafilcon A** | **Comfilcon A** | **Delefilcon A** | **Etafilcon A** | **Lotrafilcon B** | **Nelfilcon A** | **Nesofilcon A** | **Senofilcon A** | **Verofilcon A** |
| P02788|TRFL_HUMAN | 4.800005 | 6.156228 | 6.312946 | 6.762103 | | 5.054875 | 6.492151 | 6.194909 | 7.773583 |
| P35527|K1C9_HUMAN | 2.585276 | 2.514096 | 2.568126 | 2.105827 | | 2.428701 | 2.090198 | 2.461693 | 2.039006 |

| | | | | | | | | |
|---|---|---|---|---|---|---|---|---|
| P04264\|K2C1_HUMAN | 4.110809 | 3.839497 | 4.121533 | 3.484943 | 3.78259 | 3.289165 | 3.924756 | 3.507104 |
| P13645\|K1C10_HUMAN | 2.942014 | 2.458142 | 3.021598 | 2.474332 | 2.295162 | 1.98468 | 2.608324 | 2.559571 |
| P35908\|K22E_HUMAN | 2.833021 | 2.202806 | 2.508756 | 2.102852 | 1.965376 | 1.743535 | 2.49954 | 2.329474 |
| P61626\|LYSC_HUMAN | 5.512686 | 6.124158 | 7.422039 | 7.764503 | 5.629909 | 8.30401 | 5.889723 | 7.113527 |
| P02768\|ALBU_HUMAN | 3.98486 | 3.231274 | 5.71633 | 6.012968 | 5.123586 | 6.050282 | 4.854313 | 4.862141 |
| P31025\|LCN1_HUMAN | 4.275856 | 5.585627 | 5.283541 | 6.398207 | 4.793559 | 6.010642 | 5.700904 | 5.268306 |
| P01833\|PIGR_HUMAN | 0.126032 | 3.556766 | 0.737658 | 2.417547 | 0.681695 | 2.438787 | 2.387517 | 1.728485 |
| P01876\|IGHA1_HUMAN | 0.281815 | 3.973085 | 0.506806 | 2.66401 | 0.876044 | 2.637669 | 2.60462 | 2.013204 |
| P01024\|CO3_HUMAN | 0.108485 | 2.513872 | 1.043191 | 2.873718 | 1.361551 | 2.809156 | 1.959542 | 1.535403 |
| P25311\|ZA2G_HUMAN | 1.216807 | 2.880192 | 2.267228 | 2.979824 | 1.666018 | 2.599341 | 2.700533 | 2.296197 |
| P04259\|K2C6B_HUMAN | -8.22675 | -8.3962 | -6.81063 | -8.19433 | -7.65801 | -7.1529 | -7.75243 | -8.41604 |
| P19013\|K2C4_HUMAN | 1.683415 | -0.12448 | 0.874693 | 1.455002 | 0.606898 | 1.547567 | -0.65232 | 3.075705 |
| P13647\|K2C5_HUMAN | 2.013755 | 0.433737 | 1.187466 | 0.985375 | 0.806564 | 0.941813 | 0.424825 | 2.895792 |
| P02538\|K2C6A_HUMAN | -3.64411 | -3.18673 | -1.61936 | -3.13734 | -3.07505 | -2.40938 | -3.32524 | -2.76278 |
| P08727\|K1C19_HUMAN | 0.435295 | -1.44803 | -0.37635 | -0.04139 | -0.80245 | 0.470194 | -1.48637 | 1.842214 |
| P13646\|K1C13_HUMAN | 0.70238 | -1.05085 | -0.4643 | 0.051303 | -0.65171 | 0.000261 | -1.56818 | 1.979043 |
| P08779\|K1C16_HUMAN | -1.0044 | -0.78656 | 0.540628 | -0.73975 | -0.63557 | -0.50417 | -0.77215 | -0.7883 |
| P02533\|K1C14_HUMAN | -1.69876 | -1.69586 | -1.08302 | -1.58344 | -1.71803 | -1.45996 | -1.66108 | -1.6341 |
| Q9UGM3\|DMBT1_HUMAN | -2.02621 | -0.08508 | -1.8093 | -0.01587 | -2.09728 | -0.05817 | -1.31912 | -0.43729 |

| | | | | | | | | |
|---|---|---|---|---|---|---|---|---|
| P02787\|TRFE_HUMAN | 0.245195 | 0.082233 | 1.862285 | 2.116209 | 1.312962 | 2.109926 | 1.19545 | 1.075453 |
| P0DOX7\|IGK_HUMAN | -0.33394 | 2.930351 | 0.558056 | 1.957464 | 0.41909 | 2.061526 | 1.781693 | 1.379221 |
| P98160\|PGBM_HUMAN | -2.27802 | -0.93946 | -1.83581 | -0.243 | -1.69786 | -0.77208 | -1.37097 | -0.91035 |
| P60709\|ACTB_HUMAN | -1.91896 | -1.84126 | -0.92135 | 0.015836 | -0.80296 | -0.00596 | -0.6236 | -0.11577 |
| P63261\|ACTG_HUMAN | -1.91896 | -1.84126 | -0.92135 | 0.015836 | -0.80296 | -0.00596 | -0.6236 | -0.11577 |
| P15924\|DESP_HUMAN | -0.79824 | -0.76968 | -0.48152 | -0.98317 | -0.72852 | -1.1641 | -0.90445 | -0.83223 |
| P06733\|ENOA_HUMAN | 0.08953 | 0.538745 | 1.323305 | 1.838814 | 1.685207 | 2.357201 | 2.343917 | 2.00138 |
| P01036\|CYTS_HUMAN | -1.74548 | -0.31802 | -0.96203 | -0.26634 | -1.91204 | 0.555054 | -0.94645 | -0.96664 |
| P12273\|PIP_HUMAN | 1.618114 | 3.302933 | 2.976601 | 3.839629 | 2.454612 | 3.247345 | 3.294509 | 3.108825 |
| Q13421\|MSLN_HUMAN | -3.57716 | -2.04043 | -3.30616 | -1.66607 | -3.25354 | -2.31643 | -2.22784 | -2.24695 |
| P01037\|CYTN_HUMAN | -4.16712 | -2.2272 | -2.7988 | -2.45559 | -3.84077 | -0.72975 | -2.86978 | -2.95871 |
| P0DOX2\|IGA2_HUMAN | -3.77751 | -0.33277 | -3.33999 | -1.6018 | -3.199 | -1.58365 | -1.50945 | -2.17961 |
| P07355\|ANXA2_HUMAN | 0.002726 | -0.84222 | 0.565851 | 0.579469 | 0.171211 | 0.810806 | -0.34688 | 1.400492 |
| P00450\|CERU_HUMAN | -2.13379 | -0.66917 | -1.40026 | -0.32187 | -1.73112 | -0.36416 | -0.93778 | -0.64606 |
| P06396\|GELS_HUMAN | -1.06588 | 0.268628 | -0.27527 | 1.242263 | 0.075183 | 1.102327 | 0.367966 | 0.506509 |
| P00738\|HPT_HUMAN | -2.72398 | -1.6496 | -1.29816 | -0.61838 | -1.84416 | -0.86265 | -1.59402 | -1.30005 |
| P0DOY2\|IGLC2_HUMAN | -3.12798 | 0.533905 | -2.62949 | -0.80564 | -2.45705 | -0.608 | -0.94015 | -1.38097 |
| B9A064\|IGLL5_HUMAN | -2.92144 | 0.445099 | -2.36375 | -0.77881 | -2.35208 | -0.60048 | -0.88832 | -1.29838 |
| P0DOX8\|IGL1_HUMAN | -2.92144 | 0.445099 | -2.36375 | -0.77881 | -2.35208 | -0.60048 | -0.88832 | -1.29838 |

| | | | | | | | | |
|---|---|---|---|---|---|---|---|---|
| P0DOX5\|IGG1_HUMAN | -1.72274 | -1.46849 | -0.08258 | 0.372793 | -1.08212 | 0.650402 | -1.12985 | -0.11957 |
| P10909\|CLUS_HUMAN | -2.29213 | -0.59376 | -2.01416 | 0.051959 | -1.44239 | -0.52975 | -0.94438 | -1.14061 |
| P08729\|K2C7_HUMAN | -1.75636 | -3.39507 | -2.32002 | -1.58932 | -2.69065 | -1.02507 | -3.47016 | -0.10724 |
| P04083\|ANXA1_HUMAN | -1.57385 | -2.3486 | -1.51046 | -0.54039 | -1.35263 | -0.51565 | -1.67401 | 0.117453 |
| P60174\|TPIS_HUMAN | -2.19305 | -1.39495 | -0.84963 | -0.31029 | -0.73395 | -0.04282 | 0.128662 | -0.3042 |
| P14923\|PLAK_HUMAN | -0.45149 | -0.55999 | 0.020167 | -0.51363 | -0.49887 | -0.60469 | -0.39638 | -0.33152 |
| P00352\|AL1A1_HUMAN | -2.32033 | -2.12452 | -1.43258 | -0.10534 | -0.77245 | 0.272302 | -0.76489 | -0.49835 |
| P01871\|IGHM_HUMAN | -8.26829 | -5.22076 | -6.44858 | -5.6998 | -7.02985 | -5.64533 | -6.35413 | -5.95476 |
| P14618\|KPYM_HUMAN | -1.64349 | -1.33013 | -1.1277 | 0.083873 | -0.7384 | 0.150999 | -0.59619 | 0.060216 |
| P05787\|K2C8_HUMAN | -2.22727 | -3.66557 | -2.92753 | -1.92047 | -2.90161 | -1.49405 | -3.94786 | -0.62872 |
| O75556\|SG2A1_HUMAN | -0.60184 | -0.56837 | -1.46868 | 0.538028 | -0.93532 | 0.925312 | 0.200054 | 0.206935 |
| P30838\|AL3A1_HUMAN | -1.56752 | -1.34925 | -1.23262 | 0.171278 | -0.21546 | 0.274414 | -0.16917 | 0.194401 |
| P0DMV8\|HS71A_HUMAN | -3.48455 | -3.87117 | -2.54792 | -1.71349 | -2.43537 | -1.68705 | -2.87444 | -1.86379 |
| P0DMV9\|HS71B_HUMAN | -3.48455 | -3.87117 | -2.54792 | -1.71349 | -2.43537 | -1.68705 | -2.87444 | -1.86379 |
| Q08380\|LG3BP_HUMAN | -2.1923 | -0.99878 | -1.65512 | -0.16481 | -1.61543 | -0.43542 | -1.11768 | -1.03584 |
| Q16378\|PROL4_HUMAN | 1.498458 | 1.793953 | 1.743367 | 2.756624 | 1.274367 | 3.178135 | 1.400755 | 1.834332 |
| Q04695\|K1C17_HUMAN | -4.46327 | -3.97887 | -2.78052 | -3.89078 | -3.68305 | -3.93596 | -4.15726 | -3.5928 |
| P68133\|ACTS_HUMAN | -5.40626 | -5.02725 | -4.35827 | -3.3088 | -3.81364 | -3.31494 | -4.00443 | -3.58662 |
| P68032\|ACTC_HUMAN | -5.40626 | -5.02725 | -4.35827 | -3.3088 | -3.81364 | -3.31494 | -4.00443 | -3.58662 |

| | | | | | | | | | |
|---|---|---|---|---|---|---|---|---|---|
| P04792\|HSPB1_HUMAN | -0.76949 | -1.09173 | -0.40613 | 1.048972 | | 0.426092 | 0.959667 | 0.123092 | 0.694445 |
| P01009\|A1AT_HUMAN | -0.45985 | -1.18869 | 0.970926 | 1.437831 | | 0.444303 | 1.427596 | 0.072118 | 0.340497 |
| P12035\|K2C3_HUMAN | -8.33374 | | -4.91505 | -5.66858 | | -7.62766 | -5.66281 | -6.59801 | -5.38895 |
| P06702\|S10A9_HUMAN | 0.441999 | 1.126935 | 1.82549 | 2.367029 | | 1.583733 | 2.991694 | 2.539499 | 2.53644 |
| P22079\|PERL_HUMAN | -3.33572 | -2.48655 | -2.3749 | -1.37753 | | -2.86083 | -1.84606 | -2.63781 | -2.10372 |
| P01859\|IGHG2_HUMAN | -3.969 | -3.85244 | -2.37511 | -1.84675 | | -2.4702 | -1.68359 | -2.79698 | -2.81146 |
| P20061\|TCO1_HUMAN | -2.34998 | -0.36295 | -1.29865 | 0.038621 | | -1.50243 | -0.52929 | -0.93242 | -0.82178 |
| Q02413\|DSG1_HUMAN | -2.3411 | -1.98109 | -1.63676 | -1.94851 | | -1.99909 | -2.36637 | -1.98932 | -1.52168 |
| P19012\|K1C15_HUMAN | -7.46788 | | -10.3988 | -9.3348 | | -9.21385 | -8.48691 | -8.33269 | -6.62935 |
| P30740\|ILEU_HUMAN | -2.5447 | -1.26568 | -1.40355 | -0.334 | | -1.11503 | -0.02934 | -0.14443 | -0.79959 |
| P80188\|NGAL_HUMAN | -2.86867 | 0.275363 | -1.25187 | 0.600125 | | -0.98394 | 0.419177 | 0.23064 | -0.96603 |
| Q99935\|PROL1_HUMAN | -0.79945 | -0.38204 | -1.08605 | 0.290059 | | -0.85082 | -0.18894 | -0.86383 | -0.17322 |
| P04406\|G3P_HUMAN | -2.86542 | -1.99792 | -1.50665 | -0.86165 | | -1.44741 | -1.12708 | -1.3502 | -0.62804 |
| P03973\|SLPI_HUMAN | -1.03242 | 0.36979 | 1.053617 | 1.688631 | | -1.04186 | 1.404423 | -0.39002 | -0.4571 |
| P98088\|MUC5A_HUMAN | -3.45543 | -1.48307 | -2.2069 | -2.12988 | | -2.34061 | -0.74789 | -2.29672 | 0.18286 |
| P14555\|PA2GA_HUMAN | -1.79717 | -0.70874 | -3.28196 | -1.33713 | | -3.8017 | -1.87269 | -1.54416 | -1.55995 |
| P01591\|IGJ_HUMAN | 0.227894 | 3.976488 | 0.123046 | 2.499061 | | 0.601177 | 2.646839 | 2.539309 | 1.912824 |
| O95968\|SG1D1_HUMAN | 0.026188 | 1.116905 | -0.31522 | 1.642025 | | -0.36444 | 1.582755 | 1.289144 | 1.313007 |
| Q7Z794\|K2C1B_HUMAN | -1.07974 | -1.26019 | -0.78371 | -1.20549 | | -1.26734 | -1.5858 | -1.26088 | -1.09147 |
| Q8N1N4\|K2C78_HUMAN | -3.30894 | -3.43208 | -3.26133 | -3.18424 | | -3.40036 | -3.58032 | -3.41819 | -2.90022 |

| | | | | | | | | |
|---|---|---|---|---|---|---|---|---|
| P63104\|1433Z_HUMAN | -2.94595 | -2.88146 | -2.07206 | -0.63136 | -1.38247 | -0.47438 | -1.79249 | -1.1654 |
| P31944\|CASPE_HUMAN | -2.26184 | -1.81128 | -0.84306 | -1.6081 | -2.09195 | -1.92294 | -1.65465 | 0.290627 |
| Q96P63\|SPB12_HUMAN | -4.15452 | -4.01481 | -3.51798 | -3.95036 | -4.19614 | -4.18356 | -4.27624 | -3.27305 |
| P09228\|CYTT_HUMAN | -7.69634 | -6.35281 | -6.79754 | -5.67493 | -7.07024 | -6.53447 | -6.35949 | -6.41746 |
| P09211\|GSTP1_HUMAN | -2.83076 | -2.02972 | -1.58343 | -0.86843 | -1.32347 | -0.91717 | -0.7435 | -0.85901 |
| Q9GZZ8\|LACRT_HUMAN | 1.798373 | 2.650169 | 1.604141 | 3.418638 | 1.955554 | 2.409913 | 2.887773 | 2.505009 |
| P02647\|APOA1_HUMAN | -1.54669 | -1.97603 | -1.25432 | -0.24187 | -0.78768 | -0.84749 | -1.5981 | -0.3936 |
| Q96DA0\|ZG16B_HUMAN | -1.25944 | -0.83399 | 0.470602 | 0.485333 | -1.309 | 0.35523 | -0.74392 | -0.04273 |
| P05109\|S10A8_HUMAN | -0.43319 | -0.44287 | 0.57618 | 1.095786 | 0.666425 | 1.759707 | 1.264626 | 1.228104 |
| P07602\|SAP_HUMAN | -3.44603 | -2.93885 | -2.9812 | -1.4636 | -2.68084 | -2.06062 | -2.93318 | -2.39639 |
| P05090\|APOD_HUMAN | -4.50764 | -2.35067 | -3.03415 | -2.34237 | -3.04874 | -2.10876 | -3.02142 | -0.44136 |
| P11142\|HSP7C_HUMAN | -4.16793 | -3.69981 | -2.83496 | -2.08392 | -2.8827 | -2.15306 | -3.07402 | -2.05312 |
| Q5D862\|FILA2_HUMAN | -3.58874 | -3.46236 | -2.72435 | -3.73837 | -3.8799 | -3.61122 | -3.51791 | -1.75439 |
| P61769\|B2MG_HUMAN | -2.70339 | -1.8904 | -2.07384 | -1.09318 | -1.77877 | 1.738462 | -1.21642 | -1.91023 |
| Q01469\|FABP5_HUMAN | -2.71656 | -2.36361 | -1.11938 | -1.13859 | -1.11296 | -0.5611 | -1.31748 | -0.58908 |
| P34096\|RNAS4_HUMAN | -3.82823 | -1.41833 | -2.29625 | -1.14688 | -2.95063 | -1.13205 | -1.76382 | -2.48256 |
| P07339\|CATD_HUMAN | -3.21181 | -2.84571 | -2.34288 | -1.78831 | -2.26459 | -1.88605 | -3.00965 | -1.5778 |
| P30044\|PRDX5_HUMAN | -3.02418 | -1.12695 | -2.62028 | -1.23656 | -2.19584 | -0.83794 | -1.82936 | -1.16428 |
| P04075\|ALDOA_HUMAN | -3.16701 | -2.84796 | -1.78494 | -1.32098 | -1.92663 | -1.20013 | -1.86358 | -1.41237 |
| P31947\|1433S_HUMAN | -5.05647 | -4.96343 | -4.2936 | -3.06293 | -3.64184 | -3.03629 | -3.84613 | -3.47467 |

| | | | | | | | | |
|---|---|---|---|---|---|---|---|---|
| Q5VTE0\|EF1A3_HUMAN | -1.90147 | -2.06582 | -1.02843 | -0.30665 | -1.18452 | -0.38071 | -1.08954 | 0.192648 |
| P68104\|EF1A1_HUMAN | -1.90147 | -2.06582 | -1.02843 | -0.30665 | -1.18452 | -0.38071 | -1.08954 | 0.192648 |
| P62937\|PPIA_HUMAN | -3.11208 | -2.30935 | -2.01852 | -1.22713 | -1.52852 | 0.515831 | -0.64778 | -1.26215 |
| P62805\|H4_HUMAN | -1.26947 | -2.82535 | -1.38895 | -1.26465 | -2.1565 | -0.8211 | -2.73209 | 0.470982 |
| P07737\|PROF1_HUMAN | -3.62909 | -2.97713 | -1.96837 | -1.35127 | -1.63042 | 0.439883 | -1.29598 | -1.32839 |
| A0M8Q6\|IGLC7_HUMAN | -8.18744 | -4.86342 | -8.08227 | -5.62597 | -7.47462 | -5.38989 | -6.16445 | -6.46658 |
| Q99456\|K1C12_HUMAN | -8.17792 | -8.82345 | -5.57474 | -6.14322 | -8.11091 | -6.17323 | -7.55336 | -5.8101 |
| Q06830\|PRDX1_HUMAN | -1.41501 | -1.71836 | -0.90889 | 0.184019 | -0.52242 | 0.549613 | -0.55612 | 0.00224 |
| P30086\|PEBP1_HUMAN | -3.10794 | -1.65849 | -1.46659 | -1.3576 | -1.65306 | -0.38782 | -0.28521 | -1.20941 |
| Q3SY84\|K2C71_HUMAN | | | | | | | | |
| P02545\|LMNA_HUMAN | -1.88812 | -3.06642 | -1.81892 | -1.88985 | -2.60975 | -1.575 | -3.59595 | -0.4652 |
| P00338\|LDHA_HUMAN | -2.19252 | -1.04656 | -0.84821 | -0.38234 | -0.95795 | 0.08776 | -0.38459 | -0.61859 |
| P01034\|CYTC_HUMAN | -3.67797 | -1.93569 | -2.80487 | -1.37535 | -2.80808 | 0.03116 | -1.91732 | -2.35578 |
| P68363\|TBA1B_HUMAN | -4.22756 | -3.5156 | -2.75342 | -2.30444 | -3.32155 | -2.43699 | -3.15687 | -1.9175 |
| P52209\|6PGD_HUMAN | -4.09393 | -3.86439 | -3.26721 | -1.93297 | -2.74265 | -1.96664 | -2.88657 | -2.33316 |
| Q8N474\|SFRP1_HUMAN | -4.09981 | -3.54076 | -2.78809 | -2.52165 | -4.07434 | -3.07158 | -3.58027 | -0.72192 |
| P01011\|AACT_HUMAN | -2.09367 | -1.60634 | -1.40629 | -0.93501 | -1.91577 | -1.03894 | -1.544 | -1.43453 |
| P17931\|LEG3_HUMAN | -1.7651 | -1.53497 | -1.11864 | 0.372543 | -0.25673 | 1.279666 | -0.56249 | -0.38335 |
| P01619\|KV320_HUMAN | -4.67457 | -1.71765 | -3.43087 | -2.50979 | -3.84732 | -2.41565 | -2.57792 | -3.32771 |
| P11021\|BIP_HUMAN | -3.39826 | -3.01078 | -2.49442 | -1.81592 | -2.79559 | -1.88318 | -3.15121 | -1.90896 |

| | | | | | | | | |
|---|---|---|---|---|---|---|---|---|
| P00558\|PGK1_HUMAN | -1.91905 | -1.87768 | -1.39843 | -0.49646 | -1.5736 | 0.148319 | -1.13226 | -0.7272 |
| P07858\|CATB_HUMAN | -4.47751 | -3.35423 | -3.31243 | -2.53397 | -3.75725 | -2.85849 | -3.39839 | -3.11983 |
| P29401\|TKT_HUMAN | -3.34967 | -2.8289 | -2.04523 | -1.46964 | -1.78564 | -1.32575 | -2.05068 | -1.78717 |
| P80303\|NUCB2_HUMAN | -2.88126 | -2.40132 | -2.72093 | -1.11437 | -2.76437 | -1.82421 | -2.57679 | -2.21287 |
| P15311\|EZRI_HUMAN | -4.19339 | -3.18595 | -3.49244 | -2.2152 | -2.95296 | -2.18717 | -2.91357 | -2.62238 |
| Q08554\|DSC1_HUMAN | -2.19157 | -2.11736 | -1.98411 | -2.26189 | -2.19194 | -2.38469 | -2.02907 | -2.01663 |
| Q93079\|H2B1H_HUMAN | -2.0224 | -3.61535 | -2.71799 | -2.29273 | -3.29325 | -1.74511 | -3.76608 | -0.38091 |
| Q5QNW6\|H2B2F_HUMAN | -2.0224 | -3.61535 | -2.71799 | -2.29273 | -3.29325 | -1.74511 | -3.76608 | -0.38091 |
| Q99877\|H2B1N_HUMAN | -2.0224 | -3.61535 | -2.71799 | -2.29273 | -3.29325 | -1.74511 | -3.76608 | -0.38091 |
| O60814\|H2B1K_HUMAN | -2.0224 | -3.61535 | -2.71799 | -2.29273 | -3.29325 | -1.74511 | -3.76608 | -0.38091 |
| P62807\|H2B1C_HUMAN | -2.0224 | -3.61535 | -2.71799 | -2.29273 | -3.29325 | -1.74511 | -3.76608 | -0.38091 |
| P58876\|H2B1D_HUMAN | -2.0224 | -3.61535 | -2.71799 | -2.29273 | -3.29325 | -1.74511 | -3.76608 | -0.38091 |
| Q99880\|H2B1L_HUMAN | -2.0224 | -3.61535 | -2.71799 | -2.29273 | -3.29325 | -1.74511 | -3.76608 | -0.38091 |
| P57053\|H2BFS_HUMAN | -2.0224 | -3.61535 | -2.71799 | -2.29273 | -3.29325 | -1.74511 | -3.76608 | -0.38091 |
| Q99879\|H2B1M_HUMAN | -2.0224 | -3.61535 | -2.71799 | -2.29273 | -3.29325 | -1.74511 | -3.76608 | -0.38091 |
| P27797\|CALR_HUMAN | -3.62191 | -3.50699 | -2.61983 | -1.54849 | -2.65958 | -1.3284 | -2.92065 | -2.26346 |
| Q6UXB2\|CXL17_HUMAN | -3.99912 | -3.15207 | -2.46356 | -2.45053 | -4.21932 | -3.04976 | -3.61376 | -2.80068 |
| A0A0B4J1X5\|HV374_HUMAN | -6.4682 | -3.36926 | -4.78129 | -4.26085 | -5.58404 | -3.9721 | -4.29162 | -4.87556 |
| P06744\|G6PI_HUMAN | -4.61077 | -3.73193 | -3.33329 | -2.97017 | -3.38112 | -2.64246 | -2.58434 | -2.8935 |
| P05783\|K1C18_HUMAN | -6.10729 | -6.91188 | -4.80289 | -5.12757 | -6.4657 | -4.99982 | -6.79535 | -4.36536 |

| | | | | | | | | |
|---|---|---|---|---|---|---|---|---|
| P01040\|CYTA_HUMAN | -3.13869 | -2.46547 | -1.42621 | -2.36772 | -2.08758 | -1.59048 | -1.68803 | -1.44009 |
| P40394\|ADH7_HUMAN | -2.53456 | -2.61464 | -1.82372 | -1.3173 | -1.67387 | -0.66781 | -1.2924 | -1.41783 |
| A0A0B4J1V0\|HV315_HUMAN | -7.77395 | -5.23294 | -6.64927 | -5.17148 | -6.56704 | -6.42499 | -6.27191 | -5.73496 |
| P80748\|LV321_HUMAN | -11.0074 | -8.11264 | -10.0993 | -8.47088 | -10.3204 | -8.81531 | | -8.90665 |
| P62979\|RS27A_HUMAN | -2.83742 | -2.6411 | -1.72775 | -1.86823 | -2.56011 | -1.60127 | -2.26068 | -1.49819 |
| P62987\|RL40_HUMAN | -2.83742 | -2.6411 | -1.72775 | -1.86823 | -2.56011 | -1.60127 | -2.26068 | -1.49819 |
| P0CG47\|UBB_HUMAN | -2.83742 | -2.6411 | -1.72775 | -1.86823 | -2.56011 | -1.60127 | -2.26068 | -1.49819 |
| P0CG48\|UBC_HUMAN | -2.83742 | -2.6411 | -1.72775 | -1.86823 | -2.56011 | -1.60127 | -2.26068 | -1.49819 |
| P16403\|H12_HUMAN | -5.25994 | -6.46641 | -6.18837 | -5.03431 | -5.91184 | -4.59746 | -6.75852 | -3.64387 |
| P27348\|1433T_HUMAN | -5.65074 | -4.67703 | -4.56887 | -4.03502 | -4.94904 | -3.74478 | -4.73286 | -4.38695 |
| P07237\|PDIA1_HUMAN | -5.24732 | -5.26369 | -4.42129 | -3.41253 | -4.5232 | -3.22759 | -4.95398 | -4.18481 |
| P00751\|CFAB_HUMAN | -4.11775 | -1.78952 | -2.82824 | -1.1933 | -2.5439 | -1.24836 | -1.67269 | -2.65231 |
| P06312\|KV401_HUMAN | -6.05144 | -3.62435 | -5.12825 | -3.84807 | -5.07528 | -3.84332 | -4.19889 | -4.45717 |
| P02765\|FETUA_HUMAN | -4.17819 | -4.8068 | -2.81337 | -1.685 | -2.77712 | -2.15058 | -4.15933 | -3.07417 |
| P10412\|H14_HUMAN | -5.66139 | -7.88212 | -7.03223 | -5.92495 | -6.86172 | -5.43086 | -8.24863 | -4.26334 |
| P31941\|ABC3A_HUMAN | -4.8811 | -3.71452 | -4.14859 | -2.08247 | -3.17633 | -0.66959 | -2.89778 | -3.13027 |
| P55058\|PLTP_HUMAN | -4.88002 | -4.36095 | -3.71751 | -3.28771 | -5.08861 | -3.54861 | -4.46156 | -3.8141 |
| P01615\|KVD28_HUMAN | -8.492 | -5.16733 | -7.51784 | -6.10082 | -7.68864 | -5.74515 | -6.19798 | -6.64324 |
| A0A075B6P5\|KV228_HUMAN | -8.492 | -5.16733 | -7.51784 | -6.10082 | -7.68864 | -5.74515 | -6.19798 | -6.64324 |
| P81605\|DCD_HUMAN | -2.11713 | -1.74379 | -2.1919 | -2.43819 | -2.26757 | -1.03575 | -2.32538 | -2.43797 |

| | | | | | | | | |
|---|---|---|---|---|---|---|---|---|
| P02774\|VTDB_HUMAN | -4.03001 | -3.73221 | -2.57825 | -2.02566 | -2.83799 | -2.16276 | -3.33107 | -3.06393 |
| P14550\|AK1A1_HUMAN | -5.06111 | -4.53755 | -4.15681 | -3.11879 | -4.03823 | -2.79969 | -3.19171 | -3.30804 |
| A0A0B4J1Y9\|HV372_HUMAN | | | | | | | | |
| P32119\|PRDX2_HUMAN | -3.46924 | -3.6043 | -3.19375 | -2.62274 | -2.9414 | -2.50002 | -2.96331 | -2.57306 |
| P26447\|S10A4_HUMAN | -2.18036 | -3.00308 | -2.14699 | -0.75659 | -1.45686 | -0.46799 | -1.54963 | -0.45839 |
| P31151\|S10A7_HUMAN | -4.41322 | -2.80393 | -2.43222 | -2.69408 | -3.39682 | -2.48856 | -2.38118 | -1.46128 |
| Q15517\|CDSN_HUMAN | -5.63585 | -5.08716 | -5.46681 | -5.54784 | -5.74312 | -5.13516 | -5.09417 | -5.50839 |
| P20930\|FILA_HUMAN | -2.07227 | -1.87375 | -1.38144 | -1.91527 | -1.96286 | -2.06178 | -1.67462 | -1.03199 |
| P0DP08\|HVD82_HUMAN | -5.56828 | -1.67425 | -4.15109 | -2.65481 | -4.31727 | -2.51385 | -2.89813 | -2.55396 |
| P01825\|HV459_HUMAN | -5.56828 | -1.67425 | -4.15109 | -2.65481 | -4.31727 | -2.51385 | -2.89813 | -2.55396 |
| P0DP07\|HV431_HUMAN | -5.56828 | -1.67425 | -4.15109 | -2.65481 | -4.31727 | -2.51385 | -2.89813 | -2.55396 |
| A0A0C4DH41\|HV461_HUMAN | -5.56828 | -1.67425 | -4.15109 | -2.65481 | -4.31727 | -2.51385 | -2.89813 | -2.55396 |
| P0DP06\|HVD34_HUMAN | -5.56828 | -1.67425 | -4.15109 | -2.65481 | -4.31727 | -2.51385 | -2.89813 | -2.55396 |
| P06331\|HV434_HUMAN | -5.56828 | -1.67425 | -4.15109 | -2.65481 | -4.31727 | -2.51385 | -2.89813 | -2.55396 |
| P01824\|HV439_HUMAN | -5.56828 | -1.67425 | -4.15109 | -2.65481 | -4.31727 | -2.51385 | -2.89813 | -2.55396 |
| A0A0A0MRZ8\|KVD11_HUMAN | | | | | | | | |
| P04433\|KV311_HUMAN | | | | | | | | |
| P31949\|S10AB_HUMAN | -3.83581 | -5.5023 | -4.00393 | -3.11262 | -3.67731 | -2.58844 | -4.44253 | -2.22611 |
| P60953\|CDC42_HUMAN | -5.14796 | -4.50172 | -4.25352 | -2.92828 | -4.16609 | -3.3848 | -4.10044 | -3.09495 |
| Q6KB66\|K2C80_HUMAN | -5.92816 | -6.16966 | -5.1843 | -5.63874 | -6.42275 | -5.31834 | -5.88164 | -5.25079 |
| P10599\|THIO_HUMAN | -3.72634 | -3.26574 | -2.5977 | -2.14613 | -2.23458 | -0.97606 | -2.89858 | -1.89604 |

| | | | | | | | | |
|---|---|---|---|---|---|---|---|---|
| P40925\|MDHC_HUMAN | -4.23394 | -3.24348 | -3.00523 | -2.45994 | -2.8659 | -2.22547 | -2.24339 | -2.546 |
| Q14515\|SPRL1_HUMAN | -6.61093 | -5.06981 | -5.78886 | -4.8622 | -6.14279 | -5.20739 | -5.45081 | -5.45607 |
| A0A075B6S5\|KV127_HUMAN | | | | | | | | |
| P01742\|HV169_HUMAN | | | | | | | | |
| A0A0C4DH31\|HV118_HUMAN | -7.33929 | -3.89548 | -6.57316 | -4.99201 | -6.52022 | -4.89485 | -5.21514 | -5.43996 |
| P23083\|HV102_HUMAN | -7.33929 | -3.89548 | -6.57316 | -4.99201 | -6.52022 | -4.89485 | -5.21514 | -5.43996 |
| P07900\|HS90A_HUMAN | -5.49927 | -4.83017 | -4.56185 | -3.11964 | -4.09028 | -3.08936 | -4.48779 | -3.77174 |
| P84243\|H33_HUMAN | -3.79291 | -3.95292 | -4.24754 | -3.55746 | -4.60805 | -3.02034 | -4.48718 | -2.07183 |
| P01700\|LV147_HUMAN | -5.18602 | -2.68171 | -4.6803 | -3.17646 | -4.52158 | -3.63704 | -3.87821 | -3.65582 |
| P23528\|COF1_HUMAN | -5.44678 | -4.94776 | -4.66902 | -3.44698 | -4.23626 | -3.34505 | -4.15594 | -3.61557 |
| P21980\|TGM2_HUMAN | -3.78539 | -3.74154 | -3.56899 | -1.78775 | -2.82714 | -1.64078 | -2.80033 | -1.80542 |
| Q09666\|AHNK_HUMAN | -2.78606 | -4.17084 | -3.14561 | -2.11003 | -2.81685 | -2.07198 | -3.54861 | -1.7327 |
| P01624\|KV315_HUMAN | -7.08842 | -4.18548 | -6.44922 | -5.27457 | -6.59033 | -4.82201 | -5.17484 | -5.901 |
| P07384\|CAN1_HUMAN | -7.9708 | -8.16592 | -7.4165 | -6.34127 | -7.26664 | -6.46193 | -7.582 | -6.25071 |
| Q6UWP8\|SBSN_HUMAN | -8.64419 | | -8.97286 | -10.0943 | -8.38343 | -8.76654 | -8.40506 | -8.55536 |
| Q8NBJ4\|GOLM1_HUMAN | | | | | | | | |
| P40926\|MDHM_HUMAN | -7.04291 | -8.22143 | -5.86882 | -5.1802 | -5.82289 | -4.77472 | -6.54722 | -5.40882 |
| P68371\|TBB4B_HUMAN | -6.29501 | -5.02515 | -5.2199 | -4.17276 | -4.80447 | -4.07775 | -4.72353 | -4.18458 |
| A0A0A0MS15\|HV349_HUMAN | -8.82955 | -5.99066 | -7.21959 | -6.69647 | -8.19057 | -6.36145 | -6.50472 | -7.21776 |
| P01593\|KVD33_HUMAN | -7.35999 | -4.99686 | -6.90601 | -5.46348 | -7.42294 | -5.28172 | -5.78177 | -5.30423 |
| P01594\|KV133_HUMAN | -7.35999 | -4.99686 | -6.90601 | -5.46348 | -7.42294 | -5.28172 | -5.78177 | -5.30423 |

| | | | | | | | | |
|---|---|---|---|---|---|---|---|---|
| P02766\|TTHY_HUMAN | -4.92283 | -5.24263 | -3.2856 | -3.06618 | -3.81618 | -3.01426 | -4.10511 | -3.77182 |
| Q08188\|TGM3_HUMAN | -7.15779 | -6.35648 | -6.81635 | -6.71375 | -6.74769 | -7.22981 | -6.64615 | -5.79761 |
| P05089\|ARGI1_HUMAN | -3.30578 | -3.27533 | -2.95379 | -3.29602 | -3.28015 | -2.91304 | -2.98681 | -2.543 |
| Q9UBT3\|DKK4_HUMAN | | | | | | | | |
| O00584\|RNT2_HUMAN | | | | | | | | |
| A0A0B4J2D9\|KVD13_HUMAN | | | | | | | | |
| P0DP09\|KV113_HUMAN | | | | | | | | |
| O75874\|IDHC_HUMAN | -7.02702 | -6.79999 | -6.58247 | -5.33544 | -6.1165 | -4.91271 | -5.89462 | -5.58638 |
| P15814\|IGLL1_HUMAN | -3.97936 | -0.58667 | -3.70178 | -1.74979 | -3.29621 | -1.62735 | -1.90409 | -2.34393 |
| P30085\|KCY_HUMAN | -6.21785 | -5.23366 | -5.50265 | -4.2532 | -5.29509 | -4.13243 | -4.90602 | -4.68405 |
| A0A0C4DH34\|HV428_HUMAN | | | | | | | | |
| P30101\|PDIA3_HUMAN | -7.56498 | -8.2625 | -7.046 | -5.62206 | -6.7035 | -5.4514 | -7.26834 | -6.13995 |
| Q14697\|GANAB_HUMAN | -5.09232 | -5.45751 | -6.66507 | -4.52209 | -5.79562 | -4.87669 | -6.67344 | -4.79228 |
| P12830\|CADH1_HUMAN | -6.27779 | -5.47739 | -5.46823 | -5.18626 | -7.69265 | -5.13621 | -6.74147 | -4.43099 |
| Q16651\|PRSS8_HUMAN | | | | | | | | |
| P13987\|CD59_HUMAN | -4.29618 | -3.40431 | -3.45375 | -1.81518 | -4.06478 | -2.00712 | -3.7248 | -2.76329 |
| O43653\|PSCA_HUMAN | -5.34399 | -2.89335 | -5.50472 | -2.54433 | -3.41847 | -2.89589 | -3.92449 | -3.88256 |
| P02763\|A1AG1_HUMAN | -6.06584 | -7.09967 | -4.33823 | -4.14898 | -4.79643 | -4.17774 | -5.25073 | -5.01036 |
| A0A0B4J1V6\|HV373_HUMAN | | | | | | | | |
| O43852\|CALU_HUMAN | -6.07257 | -4.74598 | -5.11594 | -3.96688 | -5.13034 | -4.63891 | -5.32508 | -4.94063 |
| P08758\|ANXA5_HUMAN | -5.46976 | -5.2618 | -4.71191 | -4.3619 | -4.71419 | -4.21678 | -4.91272 | -4.08397 |
| Q5T749\|KPRP_HUMAN | -7.06845 | -6.8413 | -6.35243 | -6.33094 | -7.15004 | -6.08271 | -6.19063 | -6.19523 |
| Q7Z5P9\|MUC19_HUMAN | -8.16107 | -7.23729 | -6.50811 | -6.36403 | -8.23951 | -6.86123 | -6.82277 | -5.44727 |

| | | | | | | | | |
|---|---|---|---|---|---|---|---|---|
| P01714\|LV319_HUMAN | | | | | | | | |
| P01717\|LV325_HUMAN | -8.16025 | -5.04367 | -7.85282 | -6.26881 | -7.57526 | -6.17939 | -6.64872 | -6.88252 |
| A0A075B6K4\|LV310_HUMAN | -8.16025 | -5.04367 | -7.85282 | -6.26881 | -7.57526 | -6.17939 | -6.64872 | -6.88252 |
| P23284\|PPIB_HUMAN | -6.61723 | -5.24628 | -4.98606 | -4.33283 | -5.37252 | -3.54947 | -4.97001 | -3.87262 |
| P12814\|ACTN1_HUMAN | -4.97388 | -4.43516 | -3.44786 | -2.52677 | -4.08753 | -2.49518 | -4.25955 | -3.08232 |
| P40199\|CEAM6_HUMAN | | | | | | | | |
| Q02818\|NUCB1_HUMAN | -7.64932 | -7.32828 | -8.03609 | -6.38404 | -7.92584 | -7.96956 | -8.01917 | -7.61123 |
| P49788\|TIG1_HUMAN | -6.58194 | -4.54477 | -5.47393 | -3.87906 | -5.34251 | -4.59307 | -4.95654 | -4.93929 |
| P06703\|S10A6_HUMAN | -2.1912 | -3.96216 | -2.39324 | -0.6731 | -1.03082 | 0.350935 | -2.27206 | -1.14158 |
| P16401\|H15_HUMAN | | | | | | | | |
| O60437\|PEPL_HUMAN | -4.71527 | -4.5745 | -4.55767 | -3.70047 | -4.62388 | -4.10128 | -4.52138 | -3.57806 |
| P47929\|LEG7_HUMAN | -9.08128 | -7.98398 | -6.06705 | -8.36113 | -8.20389 | -9.02615 | | -8.03894 |
| Q14508\|WFDC2_HUMAN | -5.66264 | -3.19849 | -4.48072 | -3.19129 | -4.68571 | -2.1067 | -2.83318 | -4.71558 |
| Q13835\|PKP1_HUMAN | | | | | | | | |
| P02750\|A2GL_HUMAN | -7.00383 | -5.56221 | -5.78697 | -4.81226 | -6.14577 | -4.91463 | -5.4201 | -5.76258 |
| P21926\|CD9_HUMAN | -5.85422 | -5.32619 | -4.91784 | -4.26445 | -5.1392 | -4.28118 | -5.63673 | -4.51685 |
| P19961\|AMY2B_HUMAN | | | | | | | | |
| P0DTE7\|AMY1B_HUMAN | | | | | | | | |
| P04746\|AMYP_HUMAN | | | | | | | | |
| P04745\|AMY1A_HUMAN | | | | | | | | |
| P0DTE8\|AMY1C_HUMAN | | | | | | | | |
| Q9UBC9\|SPRR3_HUMAN | | | | | | | | |
| Q96QA5\|GSDMA_HUMAN | -7.51023 | -8.14354 | -7.40532 | -6.87876 | -7.80045 | -6.82645 | -7.21608 | -7.31294 |
| Q13296\|SG2A2_HUMAN | | | | | | | | |

| | | | | | | | | |
|---|---|---|---|---|---|---|---|---|
| Q14002\|CEAM7_HUMAN | | | | | | | | |
| P62753\|RS6_HUMAN | | | | | | | | |
| P09429\|HMGB1_HUMAN | -4.30619 | -1.71702 | -3.34483 | -1.70355 | -2.77246 | -2.4302 | -1.90966 | -2.94153 |
| P83731\|RL24_HUMAN | -4.24888 | -1.49214 | -3.60613 | -2.15367 | -3.05259 | -2.33765 | -1.51951 | -4.78521 |
| Q96S96\|PEBP4_HUMAN | -8.43422 | -7.01777 | -7.70114 | -6.73085 | -7.9309 | -6.92821 | -6.97107 | -6.29901 |
| Q5SNV9\|CA167_HUMAN | | | | | | | | |
| Q6ZR08\|DYH12_HUMAN | | | | | | | | |
| P07910\|HNRPC_HUMAN | | | | | | | | |
| Q9UKZ1\|CNO11_HUMAN | | | | | | | | |
| P20933\|ASPG_HUMAN | -8.91962 | -7.05798 | -7.78844 | -7.08251 | -7.47987 | -6.89319 | -6.87673 | -6.68542 |
| Q8IZL8\|PELP1_HUMAN | | | | | | | | |
| P00505\|AATM_HUMAN | | | | | | | | |
| P48634\|PRC2A_HUMAN | | | | | | | | |
| Q96G74\|OTUD5_HUMAN | | | | | | | | |
| Q8IVF2\|AHNK2_HUMAN | | | | | | | | |
| Q8TF72\|SHRM3_HUMAN | | | | | | | | |
| Q14966\|ZN638_HUMAN | | | | | | | | |
| Q7LBE3\|S26A9_HUMAN | | | | | | | | |
| A6NKD9\|CC85C_HUMAN | | | | | | | | |
| A0A0B4J1U7\|HV601_HUMAN | | | | | | | | |
| Q6PID8\|KLD10_HUMAN | | | | | | | | |
| Q99574\|NEUS_HUMAN | -8.56902 | -7.88389 | -8.4361 | -7.08585 | -8.1909 | -7.05565 | -8.65176 | -7.47865 |
| Q14526\|HIC1_HUMAN | | | | | | | | |
| Q5VSY0\|GKAP1_HUMAN | | | | | | | | |
| Q8TCU6\|PREX1_HUMAN | | | | | | | | |
| Q7Z2W7\|TRPM8_HUMAN | | | | | | | | |
| Q14106\|TOB2_HUMAN | | | | | | | | |
| Q9GZQ3\|COMD5_HUMAN | | | | | | | | |
| Q8NHQ8\|RASF8_HUMAN | | | | | | | | |
| P01703\|LV140_HUMAN | | | | | | | | |

| | | | | | | | | | |
|---|---|---|---|---|---|---|---|---|---|
| O75882\|ATRN_HUMAN | | | | | | | | | |
| O60902\|SHOX2_HUMAN | | | | | | | | | |
| Q5VSP4\|LC1L1_HUMAN | | | | | | | | | |
| P01861\|IGHG4_HUMAN | -4.91207 | -5.04155 | -3.38423 | -2.90468 | | -3.58795 | -2.78477 | -4.26827 | -3.79868 |
| O95678\|K2C75_HUMAN | -5.61133 | -5.37615 | -4.58762 | -4.96475 | | -5.40254 | -4.91469 | -5.35821 | -2.53656 |
| P0DOX6\|IGM_HUMAN | | | | | | | | | |
| Q86YZ3\|HORN_HUMAN | -7.27215 | -7.26899 | -7.81488 | -9.86051 | | -8.71528 | -6.79401 | -7.69767 | -9.18194 |
| Q14525\|KT33B_HUMAN | | | | | | | | | |
| Q562R1\|ACTBL_HUMAN | | | | | | | | | |
| P31946\|1433B_HUMAN | -6.79135 | -8.43515 | -5.15249 | -4.54754 | | -5.20978 | -4.55441 | -5.53669 | -5.63332 |
| Q7Z3Y8\|K1C27_HUMAN | | | | | | | | | |
| P01780\|HV307_HUMAN | -6.80378 | -3.86966 | -5.98537 | -4.77197 | | -6.01753 | -4.6396 | -4.69831 | -5.51699 |
| A0A0J9YX35\|HV64D_HUMAN | | | | | | | | | |
| A0A0J9YXX1\|HV5X1_HUMAN | | | | | | | | | |
| A0A0C4DH38\|HV551_HUMAN | -6.58894 | -3.31501 | -5.75118 | -4.40615 | | -5.78328 | -4.21953 | -4.58842 | -5.30509 |
| P28799\|GRN_HUMAN | -9.16222 | -9.06958 | -9.43948 | -7.1543 | | -8.46922 | -7.68361 | -9.18824 | -8.06698 |
| P01762\|HV311_HUMAN | | | | | | | | | |
| Q7Z3Y9\|K1C26_HUMAN | | | | | | | | | |
| Q15149\|PLEC_HUMAN | -5.4555 | -8.37843 | -5.93255 | -6.31615 | | -6.66568 | -6.0304 | -7.30211 | -4.4488 |
| Q7Z406\|MYH14_HUMAN | -6.51801 | -6.82209 | -6.24599 | -5.0996 | | -6.51595 | -5.07186 | -6.17948 | -5.05347 |
| P06731\|CEAM5_HUMAN | -5.08703 | -3.39296 | -4.17574 | -2.93272 | | -3.8197 | -3.14534 | -3.92199 | -3.07586 |
| P04632\|CPNS1_HUMAN | -5.6949 | -5.92547 | -5.78485 | -4.16763 | | -5.22338 | -4.25169 | -5.84309 | -4.19521 |
| Q07666\|KHDR1_HUMAN | | | | | | | | | |
| O60716\|CTND1_HUMAN | | | | | | | | | |
| P01611\|KVD12_HUMAN | | | | | | | | | |

| | | | | | | | | | |
|---|---|---|---|---|---|---|---|---|---|
| A0A0C4DH72\|KV106_HUMAN | -7.39738 | -4.87933 | -7.25759 | -6.51427 | | -7.13025 | -5.03281 | -5.95253 | -6.06684 |
| P08246\|ELNE_HUMAN | -8.19162 | -6.10132 | -6.53036 | -6.10126 | | -7.29232 | -5.89295 | -6.30654 | -4.37846 |
| A0A075B6R9\|KVD24_HUMAN | | | | | | | | | |
| A0A0C4DH68\|KV224_HUMAN | | | | | | | | | |
| P40121\|CAPG_HUMAN | | | | | | | | | |
| O76021\|RL1D1_HUMAN | | | | | | | | | |
| A0A075B6H7\|KV37_HUMAN | | | | | | | | | |
| P42357\|HUTH_HUMAN | -9.89324 | -8.95714 | -10.0935 | -9.79523 | | -9.79649 | -9.92233 | | -8.981 |
| P13797\|PLST_HUMAN | -7.9307 | -7.21131 | -7.25345 | -5.46312 | | -6.1685 | -5.76487 | -6.2574 | -6.02897 |
| P04211\|LV743_HUMAN | -6.149 | -2.80097 | -5.41374 | -3.55872 | | -5.31662 | -3.864 | -4.19606 | -4.21531 |
| A0A075B6I9\|LV746_HUMAN | -6.149 | -2.80097 | -5.41374 | -3.55872 | | -5.31662 | -3.864 | -4.19606 | -4.21531 |
| P55064\|AQP5_HUMAN | -6.93952 | -6.45519 | -7.62426 | -5.2467 | | -6.59373 | -4.6039 | -6.58588 | -5.74359 |
| P07951\|TPM2_HUMAN | | | | | | | | | |
| P09493\|TPM1_HUMAN | | | | | | | | | |
| P22735\|TGM1_HUMAN | -5.83481 | -5.57766 | -5.2194 | -5.51134 | | -5.7776 | -5.77444 | -5.61703 | -5.11588 |
| P01701\|LV151_HUMAN | -6.80291 | -3.87512 | -6.04436 | -4.9774 | | -6.36271 | -4.48782 | -4.78764 | -5.38533 |
| P49840\|GSK3A_HUMAN | | | | | | | | | |
| P0DOX3\|IGD_HUMAN | | | | | | | | | |
| P10809\|CH60_HUMAN | | | | | | | | | |
| P02679\|FIBG_HUMAN | | | | | | | | | |
| Q9UPP2\|IQEC3_HUMAN | | | | | | | | | |
| A0A075B6I0\|LV861_HUMAN | -7.08247 | -4.13818 | -6.63797 | -5.09069 | | -6.08893 | -5.02981 | -5.20283 | -5.74729 |
| Q86V81\|THOC4_HUMAN | | | | | | | | | |
| P22681\|CBL_HUMAN | | | | | | | | | |
| Q9BVC4\|LST8_HUMAN | | | | | | | | | |
| Q9BWS9\|CHID1_HUMAN | | | | | | | | | |
| Q7RTR2\|NLRC3_HUMAN | | | | | | | | | |

| | | | | | | | | | |
|---|---|---|---|---|---|---|---|---|---|
| Q8N6G6\|ATL1_HUMAN | | | | | | | | | |
| P67809\|YBOX1_HUMAN | | | | | | | | | |
| Q8IZ21\|PHAR4_HUMAN | | | | | | | | | |
| Q8WXS5\|CCG8_HUMAN | | | | | | | | | |
| Q9ULD2\|MTUS1_HUMAN | | | | | | | | | |
| Q86SM8\|MRGRE_HUMAN | | | | | | | | | |
| Q13283\|G3BP1_HUMAN | | | | | | | | | |
| Q9NSA2\|KCND1_HUMAN | | | | | | | | | |
| Q15323\|K1H1_HUMAN | -4.17708 | -4.93801 | -4.24206 | -4.18249 | | -4.79533 | -4.44926 | -5.99873 | -1.95164 |
| O43790\|KRT86_HUMAN | -4.48144 | -3.39891 | -2.53705 | -3.00303 | | -3.65534 | -2.74812 | -3.55749 | -0.76342 |
| Q14533\|KRT81_HUMAN | -4.48144 | -3.39891 | -2.53705 | -3.00303 | | -3.65534 | -2.74812 | -3.55749 | -0.76342 |
| P62258\|1433E_HUMAN | -4.11569 | -4.34477 | -3.05917 | -2.12323 | | -2.93685 | -2.06779 | -3.45932 | -2.40907 |
| Q13228\|SBP1_HUMAN | -4.40748 | -3.65098 | -3.17256 | -2.40094 | | -3.10015 | -2.11609 | -1.91753 | -2.3252 |
| P78386\|KRT85_HUMAN | -7.25786 | -6.9796 | -7.22482 | -7.16058 | | -8.31982 | -6.17409 | -7.0963 | -4.57443 |
| P13929\|ENOB_HUMAN | -6.50825 | -6.34273 | -5.22486 | -4.88355 | | -5.40959 | -4.45062 | -4.87252 | -4.86023 |
| A0A0C4DH42\|HV366_HUMAN | -6.89198 | -4.96209 | -6.39115 | -5.35685 | | -6.73003 | -5.28739 | -6.10449 | -5.88278 |
| P02790\|HEMO_HUMAN | -3.43745 | -2.97528 | -1.74519 | -1.35793 | | -2.29697 | -1.20155 | -2.19095 | -2.31953 |
| P22392\|NDKB_HUMAN | -3.73656 | -3.13496 | -2.56887 | -1.82875 | | -2.28584 | -1.37721 | -2.08843 | -1.75179 |
| P37802\|TAGL2_HUMAN | -5.7052 | -5.85157 | -5.2137 | -3.95425 | | -4.85253 | -3.33181 | -4.94956 | -3.92075 |
| P29508\|SPB3_HUMAN | -4.51857 | -3.70505 | -1.96383 | -4.00041 | | -3.92863 | -3.98598 | -3.65727 | -1.31898 |
| P18510\|IL1RA_HUMAN | -4.70884 | -3.29074 | -3.17032 | -2.57111 | | -3.57835 | -1.33317 | -2.00326 | -3.02583 |
| P30041\|PRDX6_HUMAN | -3.47651 | -2.61504 | -1.5214 | -1.17344 | | -2.2684 | -1.15377 | -2.18669 | -1.59477 |
| P15259\|PGAM2_HUMAN | -5.11342 | -4.55631 | -4.2178 | -3.389 | | -3.55468 | -2.62181 | -3.12219 | -3.30111 |

| | | | | | | | | |
|---|---|---|---|---|---|---|---|---|
| P18669\|PGAM1_HUMAN | -5.11342 | -4.55631 | -4.2178 | -3.389 | -3.55468 | -2.62181 | -3.12219 | -3.30111 |
| P48594\|SPB4_HUMAN | -9.82264 | -8.46948 | -7.79307 | -8.15757 | -9.14716 | -8.57415 | -8.56066 | -7.77155 |
| P0DP24\|CALM2_HUMAN | -5.65717 | -4.59385 | -4.44521 | -3.86491 | -4.49524 | -3.23203 | -4.40989 | -4.70983 |
| P0DP23\|CALM1_HUMAN | -5.65717 | -4.59385 | -4.44521 | -3.86491 | -4.49524 | -3.23203 | -4.40989 | -4.70983 |
| P0DP25\|CALM3_HUMAN | -5.65717 | -4.59385 | -4.44521 | -3.86491 | -4.49524 | -3.23203 | -4.40989 | -4.70983 |
| P05387\|RLA2_HUMAN | -7.291 | -8.16967 | -6.49571 | -4.72647 | -5.72775 | -5.69407 | -6.63956 | -6.38512 |
| P04080\|CYTB_HUMAN | -5.41787 | -4.87593 | -3.72668 | -3.33346 | -3.78392 | -2.04119 | -3.35767 | -3.79937 |
| P22626\|ROA2_HUMAN | -6.1705 | -5.83755 | -5.04964 | -4.08416 | -5.12345 | -3.89426 | -6.14347 | -4.76078 |
| O75223\|GGCT_HUMAN | -3.99396 | -3.92174 | -2.63482 | -2.52724 | -3.02415 | -2.98629 | -3.27432 | -1.91831 |
| P37837\|TALDO_HUMAN | -4.91886 | -5.07225 | -3.92318 | -2.76881 | -3.37951 | -2.60424 | -4.00236 | -3.35693 |
| P27482\|CALL3_HUMAN | -7.63055 | -9.08008 | -6.16193 | -5.73248 | -6.37738 | -5.79767 | -7.69015 | -6.0121 |
| O00299\|CLIC1_HUMAN | -7.33671 | -6.09223 | -6.46397 | -5.44488 | -6.48856 | -5.02357 | -5.58803 | -5.72695 |
| P08582\|TRFM_HUMAN | -7.65534 | -5.85169 | -6.45439 | -4.7333 | -5.87237 | -6.70026 | -6.0295 | -5.01724 |
| Q9HC84\|MUC5B_HUMAN | | | -9.98007 | -10.081 | -11.1434 | | | -9.91286 |
| P0DME0\|SETLP_HUMAN | -6.56103 | -6.44724 | -5.79162 | -4.69304 | -5.67568 | -4.67097 | -6.32707 | -5.17235 |
| Q01105\|SET_HUMAN | -6.56103 | -6.44724 | -5.79162 | -4.69304 | -5.67568 | -4.67097 | -6.32707 | -5.17235 |
| P02511\|CRYAB_HUMAN | -3.2947 | -3.9123 | -3.21266 | -2.2971 | -2.5448 | -2.25792 | -3.11721 | -2.44593 |
| P01008\|ANT3_HUMAN | -4.78747 | -5.75364 | -3.37387 | -2.54329 | -3.43272 | -2.62144 | -3.98387 | -3.28741 |
| P08571\|CD14_HUMAN | -7.81051 | -6.62411 | -7.49835 | -6.26856 | -7.42417 | -5.92799 | -7.02679 | -6.0694 |

| | | | | | | | | |
|---|---|---|---|---|---|---|---|---|
| A0A075B6S9\|KV137_HUMAN | -7.61498 | -4.85708 | -7.04683 | -5.70781 | -7.20755 | -5.45858 | -6.31846 | -5.97282 |
| P0DSN7\|KVD37_HUMAN | -7.61498 | -4.85708 | -7.04683 | -5.70781 | -7.20755 | -5.45858 | -6.31846 | -5.97282 |
| P06454\|PTMA_HUMAN | -5.72791 | -6.89739 | -4.62996 | -3.42207 | -4.446 | -3.32724 | -4.58215 | -4.70171 |
| P08294\|SODE_HUMAN | -7.38256 | -6.15283 | -5.93242 | -5.17165 | -6.04001 | -4.59804 | -5.4324 | -6.11265 |
| Q9HAV0\|GBB4_HUMAN | -6.62464 | -7.21588 | -7.12097 | -5.60596 | -6.47456 | -5.6108 | -7.35845 | -5.1293 |
| P62873\|GBB1_HUMAN | -6.62464 | -7.21588 | -7.12097 | -5.60596 | -6.47456 | -5.6108 | -7.35845 | -5.1293 |
| P62879\|GBB2_HUMAN | -6.62464 | -7.21588 | -7.12097 | -5.60596 | -6.47456 | -5.6108 | -7.35845 | -5.1293 |
| P16520\|GBB3_HUMAN | -6.62464 | -7.21588 | -7.12097 | -5.60596 | -6.47456 | -5.6108 | -7.35845 | -5.1293 |
| P25705\|ATPA_HUMAN | -4.37077 | -5.40433 | -4.12356 | -4.33505 | -4.74399 | -4.40481 | -5.16939 | -3.71175 |
| P08670\|VIME_HUMAN | -8.08042 | -7.69675 | -6.6943 | -6.33172 | -7.48986 | -6.26106 | -7.44567 | -6.13101 |
| P61158\|ARP3_HUMAN | -9.05756 | -8.72536 | -8.1035 | -6.49922 | -7.51775 | -7.10121 | -8.26006 | -6.62985 |
| Q9H0U4\|RAB1B_HUMAN | -7.1847 | -7.97987 | -7.11777 | -5.28224 | -6.60249 | -5.26575 | -6.7804 | -5.1925 |
| Q92928\|RAB1C_HUMAN | -7.1847 | -7.97987 | -7.11777 | -5.28224 | -6.60249 | -5.26575 | -6.7804 | -5.1925 |
| P62820\|RAB1A_HUMAN | -7.1847 | -7.97987 | -7.11777 | -5.28224 | -6.60249 | -5.26575 | -6.7804 | -5.1925 |
| Q92930\|RAB8B_HUMAN | -7.1847 | -7.97987 | -7.11777 | -5.28224 | -6.60249 | -5.26575 | -6.7804 | -5.1925 |
| P61026\|RAB10_HUMAN | -7.1847 | -7.97987 | -7.11777 | -5.28224 | -6.60249 | -5.26575 | -6.7804 | -5.1925 |
| P59190\|RAB15_HUMAN | -7.1847 | -7.97987 | -7.11777 | -5.28224 | -6.60249 | -5.26575 | -6.7804 | -5.1925 |
| Q15286\|RAB35_HUMAN | -7.1847 | -7.97987 | -7.11777 | -5.28224 | -6.60249 | -5.26575 | -6.7804 | -5.1925 |
| P51153\|RAB13_HUMAN | -7.1847 | -7.97987 | -7.11777 | -5.28224 | -6.60249 | -5.26575 | -6.7804 | -5.1925 |

| | | | | | | | | |
|---|---|---|---|---|---|---|---|---|
| P61006\|RAB8A_HUMAN | -7.1847 | -7.97987 | -7.11777 | -5.28224 | -6.60249 | -5.26575 | -6.7804 | -5.1925 |
| P61160\|ARP2_HUMAN | -7.6351 | -7.59788 | -6.70616 | -5.68873 | -6.46928 | -5.25101 | -6.39611 | -5.68223 |
| P06576\|ATPB_HUMAN | -7.50531 | -9.81627 | -7.12065 | -6.84683 | -6.90698 | -6.7732 | | -6.121 |
| P60660\|MYL6_HUMAN | -5.19404 | -5.07053 | -4.33036 | -3.56059 | -4.54523 | -3.74432 | -4.668 | -3.84131 |
| P14649\|MYL6B_HUMAN | -5.19404 | -5.07053 | -4.33036 | -3.56059 | -4.54523 | -3.74432 | -4.668 | -3.84131 |
| P05386\|RLA1_HUMAN | -8.29723 | -8.42305 | -7.15994 | -6.25613 | -6.95541 | -6.44121 | -8.41282 | -7.25191 |
| Q14764\|MVP_HUMAN | -7.30854 | -9.04394 | -7.98702 | -6.57124 | -7.60539 | -7.50629 | -7.98846 | -6.11973 |
| Q14118\|DAG1_HUMAN | -7.98463 | -7.08307 | -7.70306 | -6.19199 | -7.27903 | -6.73243 | -7.22692 | -6.25563 |
| P04217\|A1BG_HUMAN | -7.44709 | -6.4902 | -6.09173 | -5.89844 | -7.25831 | -5.56164 | -6.11256 | -6.30114 |
| P36952\|SPB5_HUMAN | -5.68609 | -5.61546 | -4.90729 | -3.8146 | -4.29227 | -3.72917 | -4.70432 | -3.72984 |
| Q02809\|PLOD1_HUMAN | -8.34341 | -7.87959 | -8.18377 | -6.92336 | -8.269 | -6.99541 | -8.61344 | -7.18841 |
| Q6ZVX7\|FBX50_HUMAN | -5.96179 | -5.70438 | -4.95305 | -4.52522 | -5.34794 | -4.54055 | -5.2544 | -5.01756 |
| P14174\|MIF_HUMAN | -7.06236 | -7.01663 | -5.85675 | -6.03936 | -5.99429 | -4.40638 | -5.8085 | -5.6304 |
| Q58FF3\|ENPLL_HUMAN | -6.34338 | -6.68887 | -5.97699 | -5.46155 | -6.55396 | -5.41358 | -6.37033 | -5.46448 |
| P14625\|ENPL_HUMAN | -6.34338 | -6.68887 | -5.97699 | -5.46155 | -6.55396 | -5.41358 | -6.37033 | -5.46448 |
| P25786\|PSA1_HUMAN | -7.37038 | -6.5536 | -5.71457 | -5.32016 | -6.194 | -5.05472 | -5.91449 | -6.03237 |
| O95436\|NPT2B_HUMAN | -8.45748 | -6.62261 | -8.98196 | -6.26085 | -7.62421 | -6.19595 | -7.32892 | -7.48391 |
| P13489\|RINI_HUMAN | -6.63335 | -6.68831 | -5.90595 | -5.0036 | -5.7202 | -4.94995 | -5.8604 | -5.18513 |
| Q9UL46\|PSME2_HUMAN | -6.8014 | -6.89024 | -5.94566 | -4.10112 | -4.95846 | -4.5136 | -5.27725 | -4.61211 |
| P36955\|PEDF_HUMAN | -9.2427 | -8.98854 | -8.42665 | -7.39017 | -8.76255 | -7.65599 | -9.28224 | -7.70928 |

| | | | | | | | | |
|---|---|---|---|---|---|---|---|---|
| P60981\|DEST_HUMAN | -7.20333 | -6.98786 | -7.21604 | -4.93522 | -5.62581 | -4.92284 | -5.62827 | -5.28683 |
| P62913\|RL11_HUMAN | -7.98183 | -8.51996 | -7.10188 | -6.48777 | -7.60015 | -5.95558 | -8.0836 | -6.33405 |
| O75888\|TNF13_HUMAN | -6.91224 | -6.54336 | -6.43524 | -6.00339 | -7.37252 | -6.36723 | -6.72843 | -4.96454 |
| P57723\|PCBP4_HUMAN | -5.45258 | -5.48868 | -5.55691 | -4.40351 | -4.96111 | -4.30208 | -5.31892 | -3.68026 |
| Q15365\|PCBP1_HUMAN | -5.45258 | -5.48868 | -5.55691 | -4.40351 | -4.96111 | -4.30208 | -5.31892 | -3.68026 |
| P57721\|PCBP3_HUMAN | -5.45258 | -5.48868 | -5.55691 | -4.40351 | -4.96111 | -4.30208 | -5.31892 | -3.68026 |
| Q15366\|PCBP2_HUMAN | -5.45258 | -5.48868 | -5.55691 | -4.40351 | -4.96111 | -4.30208 | -5.31892 | -3.68026 |
| P15309\|PPAP_HUMAN | -9.47002 | | -8.99957 | -8.0643 | -9.06233 | -8.5929 | -8.71589 | -8.10219 |
| P62491\|RB11A_HUMAN | -8.16482 | -8.92975 | -7.3095 | -6.43707 | -7.22432 | -6.99127 | -8.89639 | -6.3383 |
| Q15907\|RB11B_HUMAN | -8.16482 | -8.92975 | -7.3095 | -6.43707 | -7.22432 | -6.99127 | -8.89639 | -6.3383 |
| Q13867\|BLMH_HUMAN | -9.52159 | -8.93319 | -8.24747 | -8.29861 | -9.00812 | -6.57861 | -8.10729 | -7.20246 |
| P33241\|LSP1_HUMAN | -6.44934 | -7.45803 | -6.38395 | -5.2852 | -5.94663 | -5.62009 | -6.12383 | -5.08728 |
| P27635\|RL10_HUMAN | -5.61205 | -6.35664 | -5.65799 | -4.98563 | -5.48902 | -4.83595 | -6.04915 | -4.52613 |
| O75083\|WDR1_HUMAN | -6.74913 | -6.08706 | -6.01812 | -5.00557 | -5.42793 | -4.62304 | -5.13631 | -5.01018 |
| P09467\|F16P1_HUMAN | -7.14372 | -6.31041 | -6.6134 | -5.01476 | -5.61244 | -4.64299 | -4.87252 | -5.45379 |
| P02452\|CO1A1_HUMAN | -11.6563 | | | | | | | |
| P18124\|RL7_HUMAN | -7.90379 | -8.98854 | -7.22401 | -6.54374 | -7.38268 | -6.01226 | -9.10881 | -6.22857 |
| P62899\|RL31_HUMAN | -6.44007 | -6.58924 | -6.10237 | -5.31583 | -6.18432 | -5.47916 | -6.24514 | -4.92432 |
| P25789\|PSA4_HUMAN | -6.95975 | -6.82376 | -5.56072 | -5.26725 | -6.28714 | -4.73726 | -5.92453 | -5.70261 |
| Q9UN76\|S6A14_HUMAN | -6.09481 | -5.00008 | -6.09953 | -4.31554 | -5.7134 | -4.03271 | -5.33993 | -5.19296 |

| | | | | | | | | |
|---|---|---|---|---|---|---|---|---|
| P06748\|NPM_HUMAN | -7.38392 | -4.91659 | -6.10708 | -6.43763 | -7.05253 | -5.3366 | -5.44122 | -5.9857 |
| O43516\|WIPF1_HUMAN | -9.54408 | -8.9209 | -8.28815 | -7.86338 | -8.79144 | -7.73466 | -8.10192 | -6.44138 |
| Q99497\|PARK7_HUMAN | -7.18607 | -6.85945 | -5.90975 | -5.49373 | -6.04711 | -5.46676 | -5.27544 | -5.18146 |
| Q86X10\|RLGPB_HUMAN | -10.54 | -9.62665 | -9.42808 | | -10.9103 | | | -9.71882 |
| P40939\|ECHA_HUMAN | -8.08297 | -6.28079 | -6.66802 | -6.03457 | -7.69186 | -6.27167 | -6.88302 | -6.46017 |
| P09668\|CATH_HUMAN | -6.33526 | -4.20765 | -5.23408 | -4.70853 | -5.62692 | -4.64903 | -4.80538 | -4.02772 |
| O14497\|ARI1A_HUMAN | -8.76794 | -7.79038 | -7.32139 | -7.06114 | -8.69659 | -6.59941 | -8.38313 | -7.63731 |
| Q13200\|PSMD2_HUMAN | -8.12463 | -8.37509 | -6.5779 | -6.58847 | -8.08088 | -5.70516 | -8.11986 | -6.68484 |
| Q96SC8\|DMTA2_HUMAN | -5.98054 | -4.32207 | -5.21147 | -4.88192 | -5.80146 | -4.69955 | -5.13216 | -5.16739 |
| Q14574\|DSC3_HUMAN | -5.44596 | -6.00447 | -5.37764 | -5.3269 | -5.49627 | -5.78739 | -5.90199 | -4.69185 |
| Q9UPN9\|TRI33_HUMAN | -9.4489 | -6.79283 | -8.63331 | -8.03309 | -8.27635 | -7.68937 | -7.1009 | -8.54784 |
| P08174\|DAF_HUMAN | -7.25235 | -4.97988 | -6.55677 | -4.7169 | -5.98898 | -5.12385 | -5.66228 | -5.92664 |
| P06727\|APOA4_HUMAN | -10.1223 | | -9.21932 | -7.96827 | -8.99801 | -8.02797 | -8.70908 | -8.48499 |
| Q5T1R4\|ZEP3_HUMAN | | | -9.8324 | | | | | -10.538 |
| Q96JM2\|ZN462_HUMAN | -8.3478 | -7.60107 | -7.94204 | -7.05639 | -7.96857 | -6.74039 | -6.12652 | -8.34219 |
| Q96RY5\|CRML_HUMAN | -7.30258 | -6.54718 | -6.50717 | -6.07265 | -7.43144 | -6.06845 | -6.68703 | -6.5525 |
| P04179\|SODM_HUMAN | -6.11379 | -5.38731 | -4.75793 | -4.09417 | -5.26823 | -3.8032 | -4.81572 | -4.52694 |

**Supplementary Table 4**

*Comparison of Protein Abundance to Protein Characteristics*

| Protein Characteristic | Comfilcon A | Delefilcon A | Etafilcon A | Lotrafilcon B | Nelfilcon A | Nesofilcon A | Senofilcon A | Verofilcon A |
|---|---|---|---|---|---|---|---|---|
| **sequence length** | -0.14439 | -0.12755 | -0.16494 | -0.1132 | -0.14865 | -0.16288 | -0.14403 | -0.13945 |
| **molecular weight** | -0.14251 | -0.12572 | -0.16248 | -0.11225 | -0.14652 | -0.16088 | -0.14206 | -0.13701 |
| **aromaticity** | 0.096878 | 0.057953 | 0.07167 | 0.067598 | 0.070676 | 0.072862 | 0.08059 | 0.062632 |
| **instability index** | -0.08959 | -0.10268 | -0.11722 | -0.04137 | -0.12734 | -0.1202 | -0.10312 | -0.11621 |
| **gravy** | 0.023184 | -0.02195 | -0.00599 | 0.000653 | 0.003694 | -0.00038 | 0.008747 | 0.009649 |
| **isoelectric point** | 0.02804 | 0.001145 | 0.007234 | 0.04542 | -0.01361 | 0.00736 | 0.015488 | 0.004496 |
| **charge at pH** | 0.046491 | 0.017927 | 0.027934 | 0.063075 | 0.008142 | 0.029575 | 0.038448 | 0.025835 |
| **molar extinction coefficient reduced** | -0.06366 | -0.09049 | -0.11308 | -0.04232 | -0.09822 | -0.11545 | -0.08071 | -0.09736 |
| **molar extinction coefficient** | -0.06406 | -0.09082 | -0.11355 | -0.04355 | -0.09867 | -0.11585 | -0.08099 | -0.09735 |
| **percent content A** | -0.07239 | -0.0598 | -0.04434 | -0.04989 | -0.04452 | -0.04957 | -0.05005 | -0.05431 |
| **percent content C** | 0.029373 | -0.02959 | -0.01652 | 0.012303 | -0.01736 | -0.01156 | 0.014679 | -0.00533 |
| **percent content D** | -0.0719 | -0.01824 | -0.01287 | -0.08685 | -0.00825 | -0.02211 | -0.05012 | -0.01734 |
| **percent content E** | -0.08682 | -0.01658 | -0.03489 | -0.11329 | -0.01695 | -0.03604 | -0.05408 | -0.03172 |
| **percent content F** | -0.02627 | -0.00347 | 0.000614 | -0.05535 | -0.00157 | -0.00019 | -0.01538 | 0.001233 |
| **percent content G** | -0.04153 | -0.0372 | -0.06056 | -0.03834 | -0.05576 | -0.05308 | -0.04984 | -0.04381 |
| **percent content H** | -0.02771 | -0.00584 | -0.02285 | -0.0277 | -0.01847 | -0.02777 | -0.02542 | -0.00731 |
| **percent content I** | -0.07075 | -0.03404 | -0.01569 | -0.11994 | -0.00574 | -0.01438 | -0.05667 | -0.00758 |
| **percent content K** | 0.111955 | 0.16656 | 0.181 | 0.086096 | 0.173561 | 0.175918 | 0.147013 | 0.161273 |
| **percent content L** | 0.051032 | 0.014933 | 0.012913 | 0.051306 | 0.012291 | 0.023228 | 0.036254 | 0.019592 |
| **percent content M** | -0.05882 | -0.03984 | -0.0391 | -0.0856 | -0.03376 | -0.02845 | -0.04289 | -0.0317 |
| **percent content N** | -0.04401 | -0.00499 | -0.02127 | -0.07644 | -0.00463 | -0.01155 | -0.031 | -0.00896 |
| **percent content P** | -0.08747 | -0.1254 | -0.13469 | -0.06734 | -0.14462 | -0.14335 | -0.10408 | -0.14854 |
| **percent content Q** | 0.061884 | 0.023445 | 0.00781 | 0.102999 | 0.006609 | 0.014846 | 0.043121 | 0.003242 |
| **percent content R** | -0.14089 | -0.12564 | -0.14123 | -0.09912 | -0.15082 | -0.14627 | -0.15125 | -0.12787 |
| **percent content S** | 0.061874 | -0.00802 | -0.01411 | 0.102021 | -0.00987 | 0.000818 | 0.026634 | -0.00246 |
| **percent content T** | 0.10674 | 0.078471 | 0.105347 | 0.08966 | 0.098689 | 0.091461 | 0.092344 | 0.114975 |
| **percent content V** | 0.089411 | 0.091066 | 0.087327 | 0.061858 | 0.103477 | 0.085292 | 0.085141 | 0.094052 |
| **percent content W** | -0.00435 | -0.07243 | -0.05463 | -0.01267 | -0.05955 | -0.0609 | -0.03716 | -0.06216 |
| **percent content Y** | 0.174929 | 0.135721 | 0.137389 | 0.169797 | 0.140365 | 0.143777 | 0.156456 | 0.127259 |

Molecular weight, aromaticity, instability index, gravy, isoelectric point, charge at pH 7.0, molar extinction coefficient, and percent amino acid content for each SCL material

**Supplementary Table 5**

*Comparison of Proteins Between Etafilcon A and Verofilcon A*

| Uniprot | Protein | Fold Change (log2) | [1] p-value |
|---|---|---|---|
| A0A075B6K4 | LV310 | 0.03 | 0.25 |
| A0A075B6P5 | KV228 | 0.19 | 0.25 |
| A0A0A0MS15 | HV349 | 0.03 | 0.25 |
| A0A0B4J1X5 | HV374 | 0.03 | 0.25 |
| A0A0C4DH31 | HV118 | 0.03 | 0.25 |
| A0A0C4DH41 | HV461 | 0.04 | 0.25 |
| A0M8Q6 | IGLC7 | 0.02 | 0.25 |
| B9A064 | IGLL5 | 0.03 | 0.25 |
| P00450 | CERU | 0.03 | 0.25 |
| P00738 | HPT | 0.02 | 0.25 |
| P01011 | AACT | 0.04 | 0.25 |
| P01024 | CO3 | 0.02 | 0.25 |
| P01036 | CYTS | 0.02 | 0.25 |
| P01037 | CYTN | 0.03 | 0.25 |
| P01591 | IGJ | 0.03 | 0.25 |
| P01593 | KVD33 | 0.08 | 0.25 |
| P01594 | KV133 | 0.08 | 0.25 |
| P01615 | KVD28 | 0.19 | 0.25 |
| P01619 | KV320 | 0.02 | 0.25 |
| P01624 | KV315 | 0.02 | 0.25 |
| P01717 | LV325 | 0.03 | 0.25 |
| P01824 | HV439 | 0.04 | 0.25 |
| P01825 | HV459 | 0.04 | 0.25 |
| P01833 | PIGR | 0.02 | 0.25 |
| P01859 | IGHG2 | 0.03 | 0.25 |
| P01871 | IGHM | 0.04 | 0.25 |
| P01876 | IGHA1 | 0.02 | 0.25 |
| P02533 | K1C14 | 0.03 | 0.25 |
| P02647 | APOA1 | 0.03 | 0.25 |

| | | | |
|---|---|---|---|
| P02787 | TRFE | 0.02 | 0.25 |
| P02788 | TRFL | 0.13 | 0.25 |
| P03973 | SLPI | -0.07 | 0.25 |
| P04264 | K2C1 | 0.04 | 0.25 |
| P04406 | G3P | 0.07 | 0.25 |
| P05089 | ARGI1 | 0.06 | 0.25 |
| P05783 | K1C18 | 0.05 | 0.25 |
| P06312 | KV401 | 0.03 | 0.25 |
| P06331 | HV434 | 0.04 | 0.25 |
| P06744 | G6PI | 0.05 | 0.25 |
| P07384 | CAN1 | 0.08 | 0.25 |
| P07737 | PROF1 | 0.08 | 0.25 |
| P09228 | CYTT | 0.02 | 0.25 |
| P0DOX2 | IGA2 | 0.03 | 0.25 |
| P0DOX5 | IGG1 | 0.02 | 0.25 |
| P0DOX7 | IGK | 0.03 | 0.25 |
| P0DOX8 | IGL1 | 0.03 | 0.25 |
| P0DOY2 | IGLC2 | 0.03 | 0.25 |
| P0DP06 | HVD34 | 0.04 | 0.25 |
| P0DP07 | HV431 | 0.04 | 0.25 |
| P0DP08 | HVD82 | 0.04 | 0.25 |
| P10599 | THIO | 0.04 | 0.25 |
| P10909 | CLUS | 0.01 | 0.25 |
| P11142 | HSP7C | 0.04 | 0.25 |
| P12035 | K2C3 | 0.05 | 0.25 |
| P12273 | PIP | 0.02 | 0.25 |
| P13645 | K1C10 | 0.05 | 0.25 |
| P14923 | PLAK | 0.03 | 0.25 |
| P15814 | IGLL1 | 0.02 | 0.25 |
| P15924 | DESP | 0.03 | 0.25 |
| P20061 | TCO1 | 0.02 | 0.25 |
| P20930 | FILA | 0.05 | 0.25 |
| P22079 | PERL | 0.04 | 0.25 |

| | | | |
|---|---|---|---|
| P23083 | HV102 | 0.03 | 0.25 |
| P23284 | PPIB | 0.05 | 0.25 |
| P25311 | ZA2G | 0.03 | 0.25 |
| P30044 | PRDX5 | 0.08 | 0.25 |
| P30086 | PEBP1 | 0.06 | 0.25 |
| P30838 | AL3A1 | 0.04 | 0.25 |
| P31941 | ABC3A | 0.03 | 0.25 |
| P31944 | CASPE | 0.07 | 0.25 |
| P32119 | PRDX2 | 0.04 | 0.25 |
| P35527 | K1C9 | 0.04 | 0.25 |
| P35908 | K22E | 0.05 | 0.25 |
| P52209 | 6PGD | 0.05 | 0.25 |
| P55058 | PLTP | 0.02 | 0.25 |
| P61769 | B2MG | 0.03 | 0.25 |
| P80748 | LV321 | 0.03 | 0.25 |
| P83731 | RL24 | -0.07 | 0.25 |
| Q02413 | DSG1 | 0.04 | 0.25 |
| Q04695 | K1C17 | 0.06 | 0.25 |
| Q08188 | TGM3 | 0.06 | 0.25 |
| Q08380 | LG3BP | 0.01 | 0.25 |
| Q08554 | DSC1 | 0.05 | 0.25 |
| Q13421 | MSLN | 0.02 | 0.25 |
| Q14515 | SPRL1 | 0.07 | 0.25 |
| Q15517 | CDSN | 0.05 | 0.25 |
| Q5D862 | FILA2 | 0.07 | 0.25 |
| Q5T749 | KPRP | 0.04 | 0.25 |
| Q6UWP8 | SBSN | 0.07 | 0.25 |
| Q7Z5P9 | MUC19 | 0.08 | 0.25 |
| Q7Z794 | K2C1B | 0.06 | 0.25 |
| Q8N474 | SFRP1 | 0.10 | 0.25 |
| Q96DA0 | ZG16B | 0.05 | 0.25 |
| Q96P63 | SPB12 | 0.04 | 0.25 |
| Q96QA5 | GSDMA | 0.04 | 0.25 |

| | | | |
|---|---|---|---|
| Q99574 | NEUS | 0.04 | 0.25 |
| Q9UGM3 | DMBT1 | 0.04 | 0.25 |
| A0A075B6I9 | LV746 | 0.01 | 0.5 |
| A0A075B6S5 | KV127 | 0.01 | 0.5 |
| A0A0A0MRZ8 | KVD11 | 0.01 | 0.5 |
| A0A0B4J1V0 | HV315 | 0.03 | 0.5 |
| A0A0B4J1V6 | HV373 | 0.01 | 0.5 |
| A0A0C4DH34 | HV428 | 0.01 | 0.5 |
| A0A0C4DH38 | HV551 | 0.01 | 0.5 |
| O00584 | RNT2 | 0.01 | 0.5 |
| O75874 | IDHC | 0.03 | 0.5 |
| O95678 | K2C75 | 0.08 | 0.5 |
| P00338 | LDHA | 0.02 | 0.5 |
| P00352 | AL1A1 | 0.03 | 0.5 |
| P00558 | PGK1 | 0.05 | 0.5 |
| P00751 | CFAB | 0.01 | 0.5 |
| P01009 | A1AT | 0.03 | 0.5 |
| P01040 | CYTA | 0.05 | 0.5 |
| P01700 | LV147 | 0.02 | 0.5 |
| P01701 | LV151 | 0.02 | 0.5 |
| P01714 | LV319 | 0.01 | 0.5 |
| P01742 | HV169 | 0.01 | 0.5 |
| P01780 | HV307 | 0.02 | 0.5 |
| P01861 | IGHG4 | 0.01 | 0.5 |
| P02538 | K2C6A | 0.03 | 0.5 |
| P02750 | A2GL | 0.04 | 0.5 |
| P02763 | A1AG1 | 0.03 | 0.5 |
| P02766 | TTHY | 0.03 | 0.5 |
| P02768 | ALBU | 0.02 | 0.5 |
| P02774 | VTDB | 0.17 | 0.5 |
| P04075 | ALDOA | 0.03 | 0.5 |
| P04211 | LV743 | 0.01 | 0.5 |
| P04259 | K2C6B | 0.02 | 0.5 |

| | | | |
|---|---|---|---|
| P04433 | KV311 | 0.01 | 0.5 |
| P04632 | CPNS1 | 0.03 | 0.5 |
| P04792 | HSPB1 | 0.02 | 0.5 |
| P05090 | APOD | 0.07 | 0.5 |
| P05109 | S10A8 | 0.03 | 0.5 |
| P06396 | GELS | 0.02 | 0.5 |
| P06702 | S10A9 | 0.04 | 0.5 |
| P06733 | ENOA | 0.04 | 0.5 |
| P07355 | ANXA2 | 0.04 | 0.5 |
| P07858 | CATB | 0.02 | 0.5 |
| P07900 | HS90A | 0.04 | 0.5 |
| P08758 | ANXA5 | 0.04 | 0.5 |
| P09211 | GSTP1 | 0.04 | 0.5 |
| P09429 | HMGB1 | -0.04 | 0.5 |
| P0DMV8 | HS71A | 0.02 | 0.5 |
| P0DMV9 | HS71B | 0.02 | 0.5 |
| P11021 | BIP | 0.03 | 0.5 |
| P12814 | ACTN1 | 0.04 | 0.5 |
| P12830 | CADH1 | 0.04 | 0.5 |
| P14550 | AK1A1 | 0.05 | 0.5 |
| P16403 | H12 | 0.05 | 0.5 |
| P17931 | LEG3 | 0.03 | 0.5 |
| P19012 | K1C15 | 0.24 | 0.5 |
| P21926 | CD9 | 0.02 | 0.5 |
| P21980 | TGM2 | 0.04 | 0.5 |
| P22735 | TGM1 | 0.04 | 0.5 |
| P27348 | 1433T | 0.03 | 0.5 |
| P28799 | GRN | 0.01 | 0.5 |
| P29401 | TKT | 0.02 | 0.5 |
| P30085 | KCY | 0.04 | 0.5 |
| P30740 | ILEU | 0.03 | 0.5 |
| P31151 | S10A7 | 0.04 | 0.5 |
| P31947 | 1433S | 0.02 | 0.5 |

| | | | |
|---|---|---|---|
| P31949 | S10AB | 0.06 | 0.5 |
| P34096 | RNAS4 | -0.01 | 0.5 |
| P40394 | ADH7 | 0.04 | 0.5 |
| P40925 | MDHC | 0.04 | 0.5 |
| P40926 | MDHM | 0.03 | 0.5 |
| P42357 | HUTH | 0.20 | 0.5 |
| P47929 | LEG7 | 0.03 | 0.5 |
| P49788 | TIG1 | 0.02 | 0.5 |
| P60174 | TPIS | 0.03 | 0.5 |
| P60709 | ACTB | 0.02 | 0.5 |
| P60953 | CDC42 | 0.04 | 0.5 |
| P61626 | LYSC | 0.02 | 0.5 |
| P62753 | RS6 | -0.02 | 0.5 |
| P62937 | PPIA | 0.04 | 0.5 |
| P63261 | ACTG | 0.02 | 0.5 |
| P68032 | ACTC | 0.04 | 0.5 |
| P68104 | EF1A1 | 0.04 | 0.5 |
| P68133 | ACTS | 0.04 | 0.5 |
| P68371 | TBB4B | 0.04 | 0.5 |
| P80188 | NGAL | 0.02 | 0.5 |
| P98088 | MUC5A | 0.09 | 0.5 |
| P98160 | PGBM | 0.02 | 0.5 |
| Q01469 | FABP5 | 0.04 | 0.5 |
| Q02818 | NUCB1 | 0.01 | 0.5 |
| Q06830 | PRDX1 | 0.02 | 0.5 |
| Q14508 | WFDC2 | 0.03 | 0.5 |
| Q16651 | PRSS8 | 0.02 | 0.5 |
| Q5VTE0 | EF1A3 | 0.04 | 0.5 |
| Q6KB66 | K2C80 | 0.03 | 0.5 |
| Q86YZ3 | HORN | 0.06 | 0.5 |
| Q8N1N4 | K2C78 | 0.03 | 0.5 |
| Q96S96 | PEBP4 | 0.04 | 0.5 |
| Q99456 | K1C12 | 0.05 | 0.5 |

| | | | |
|---|---|---|---|
| Q99935 | PROL1 | 0.02 | 0.5 |
| Q9GZZ8 | LACRT | -0.02 | 0.5 |
| Q9UBT3 | DKK4 | 0.06 | 0.5 |
| O75556 | SG2A1 | -0.02 | 0.75 |
| O95968 | SG1D1 | 0.00 | 0.75 |
| P01034 | CYTC | 0.00 | 0.75 |
| P04083 | ANXA1 | 0.03 | 0.75 |
| P07339 | CATD | 0.02 | 0.75 |
| P07602 | SAP | 0.01 | 0.75 |
| P08727 | K1C19 | 0.02 | 0.75 |
| P08729 | K2C7 | 0.04 | 0.75 |
| P08779 | K1C16 | 0.02 | 0.75 |
| P0CG47 | UBB | 0.02 | 0.75 |
| P0CG48 | UBC | 0.02 | 0.75 |
| P13647 | K2C5 | 0.04 | 0.75 |
| P13987 | CD59 | 0.01 | 0.75 |
| P14555 | PA2GA | -0.04 | 0.75 |
| P14618 | KPYM | 0.01 | 0.75 |
| P23528 | COF1 | 0.01 | 0.75 |
| P31025 | LCN1 | 0.00 | 0.75 |
| P62979 | RS27A | 0.02 | 0.75 |
| P62987 | RL40 | 0.02 | 0.75 |
| P63104 | 1433Z | 0.02 | 0.75 |
| P68363 | TBA1B | 0.01 | 0.75 |
| P80303 | NUCB2 | 0.00 | 0.75 |
| P81605 | DCD | 0.02 | 0.75 |
| Q16378 | PROL4 | 0.01 | 0.75 |
| Q6UXB2 | CXL17 | -0.02 | 0.75 |
| A0A075B6I0 | LV861 | 0.01 | 1 |
| A0A0C4DH72 | KV106 | 0.03 | 1 |
| O43653 | PSCA | -0.01 | 1 |
| O43852 | CALU | 0.00 | 1 |
| O60437 | PEPL | 0.00 | 1 |

| | | | |
|---|---|---|---|
| O60814 | H2B1K | 0.00 | 1 |
| P02545 | LMNA | 0.01 | 1 |
| P02765 | FETUA | 0.01 | 1 |
| P05787 | K2C8 | 0.00 | 1 |
| P06703 | S10A6 | 0.00 | 1 |
| P06731 | CEAM5 | 0.01 | 1 |
| P07237 | PDIA1 | 0.01 | 1 |
| P08246 | ELNE | 0.04 | 1 |
| P10412 | H14 | 0.00 | 1 |
| P13646 | K1C13 | 0.02 | 1 |
| P13797 | PLST | -0.01 | 1 |
| P15311 | EZRI | 0.01 | 1 |
| P16401 | H15 | -0.04 | 1 |
| P19013 | K2C4 | 0.02 | 1 |
| P20933 | ASPG | 0.00 | 1 |
| P26447 | S10A4 | 0.00 | 1 |
| P27797 | CALR | -0.01 | 1 |
| P30101 | PDIA3 | 0.00 | 1 |
| P31946 | 1433B | -0.01 | 1 |
| P55064 | AQP5 | -0.01 | 1 |
| P57053 | H2BFS | 0.00 | 1 |
| P58876 | H2B1D | 0.00 | 1 |
| P62805 | H4 | 0.01 | 1 |
| P62807 | H2B1C | 0.00 | 1 |
| P84243 | H33 | 0.00 | 1 |
| Q09666 | AHNK | 0.00 | 1 |
| Q14697 | GANAB | -0.14 | 1 |
| Q15149 | PLEC | 0.03 | 1 |
| Q5QNW6 | H2B2F | 0.00 | 1 |
| Q7Z406 | MYH14 | -0.16 | 1 |
| Q8NBJ4 | GOLM1 | 0.00 | 1 |
| Q93079 | H2B1H | 0.00 | 1 |
| Q99877 | H2B1N | 0.00 | 1 |

| | | | |
|---|---|---|---|
| Q99879 | H2B1M | 0.00 | 1 |
| Q99880 | H2B1L | 0.00 | 1 |
| Q9UBC9 | SPRR3 | 0.02 | 1 |

[1] p-values calculated using the Wilcoxon Signed-Rank test
